# Socioeconomic status determines COVID-19 incidence and related mortality in Santiago, Chile

**DOI:** 10.1101/2021.01.12.21249682

**Authors:** Gonzalo Mena, Pamela P. Martinez, Ayesha S. Mahmud, Pablo A. Marquet, Caroline O. Buckee, Mauricio Santillana

## Abstract

The current coronavirus disease 2019 (COVID-19) pandemic has impacted dense urban populations particularly hard. Here, we provide an in-depth characterization of disease incidence and mortality patterns, and their dependence on demographic and socioeconomic strata in Santiago, a highly segregated city and the capital of Chile. We find that among all age groups, there is a strong association between socioeconomic status and both mortality –measured either by direct COVID-19 attributed deaths or excess deaths– and public health capacity. Specifically, we show that behavioral factors like human mobility, as well as health system factors such as testing volumes, testing delays, and test positivity rates are associated with disease outcomes. These robust patterns suggest multiple possibly interacting pathways that can explain the observed disease burden and mortality differentials: (i) in lower socioeconomic status municipalities, human mobility was not reduced as much as in more affluent municipalities; (ii) testing volumes in these locations were insufficient early in the pandemic and public health interventions were applied too late to be effective; (iii) test positivity and testing delays were much higher in less affluent municipalities, indicating an impaired capacity of the health-care system to contain the spread of the epidemic; and (iv) infection fatality rates appear much higher in the lower end of the socioeconomic spectrum. Together, these findings highlight the exacerbated consequences of health-care inequalities in a large city of the developing world, and provide practical methodological approaches useful for characterizing COVID-19 burden and mortality in other segregated urban centers.

## Introduction

The coronavirus disease 2019 (COVID-19) pandemic is an ongoing public health crisis. While many studies have described the transmission of SARS-CoV-2 –the virus that causes COVID-19– in North America, Europe, and parts of Asia [1–5], the characterization of the pandemic in South America has received less attention, despite the severe impact in many countries during the Southern Hemisphere winter season. While confirmed COVID-19 cases are an important public health measure to estimate the level of spread of infections caused by SARS-CoV-2, they may not be a complete indicator of COVID-19 incidence since they may be biased due to population-level health-seeking behavior, surveillance capacities, and the presence of asymptomatic individuals across regions [6]. Analyses of COVID-19 deaths and excess deaths provide an alternative and potentially less biased picture of the epidemic intensity [7, 8]. This is in part because ascertainment biases may be less pronounced for deaths than for confirmed cases as people dying from COVID-19 are more likely to have experienced severe symptoms and thus, more likely to have been documented as COVID-19 positive cases by health surveillance systems. Age specific death data may help explain the heterogeneity in COVID-19 burden and COVID-19 attributable death numbers observed among different countries [9]. However, the role of other factors, such as socioeconomic status – which is correlated with health care access– on the fatality and disease burden, remains an open question [10], especially in highly heterogeneous urban settings.

Here, we analyzed incidence and mortality attributed to SARS-CoV-2 infection and its association with demographic and socioeconomic status across the urban metropolitan area of the capital of Chile, known as ‘Greater Santiago’. To understand spatial variations in disease burden, we first estimated excess deaths and infection fatality rates across this urban area. To understand disparities in the health care system we analyzed testing capacity and delays across municipalities. We then demonstrate strong associations of these health indicators with demographic and socioeconomic factors. Together, our results show that socioeconomic disparities explain a large part of the variation in COVID-19 deaths and under-reporting, and that those inequalities disproportionately affected younger people.

### Socioeconomic differences in disease incidence, COVID-19 attributed deaths, and human movement

The Greater Santiago area is composed of 34 municipalities –defined as having more than 95% of its area urbanized– and is home to almost 7 million people. While this urban center accounts for 36% of the country’s population, it has reported 55% of the confirmed COVID-19 cases and 65% of the COVID-19 attributed deaths prior to epidemiological week 36 (end of August 2020). Socioeconomic status (SES) in the municipalities varies widely, with Vitacura having the highest value (SES = 93.7) and La Pintana the lowest one (SES = 17.0; Fig. 1A), and this difference is reflected in the impact of the pandemic during the Southern Hemisphere winter of 2020. The maximum incidence in Vitacura was 22.6 weekly cases per 10,000 individuals during the middle of May, while La Pintana reported a maximum of 76.4 weekly cases per 10,000 individuals during the first week of June (Fig. 1B). As shown in figure 1C, the attributed COVID-19 deaths follow a similar pattern, with the highest rate of 4.4 weekly deaths per 10,000 individuals reported during epidemiological week 24 (second week of June) in San Ramon, a municipality with a SES of 19.7, and less than 1 weekly deaths per 10,000 in Vitacura. These social inequalities impact the overall COVID-19 mortality rates as shown in figure 1D.

**Figure 1:**
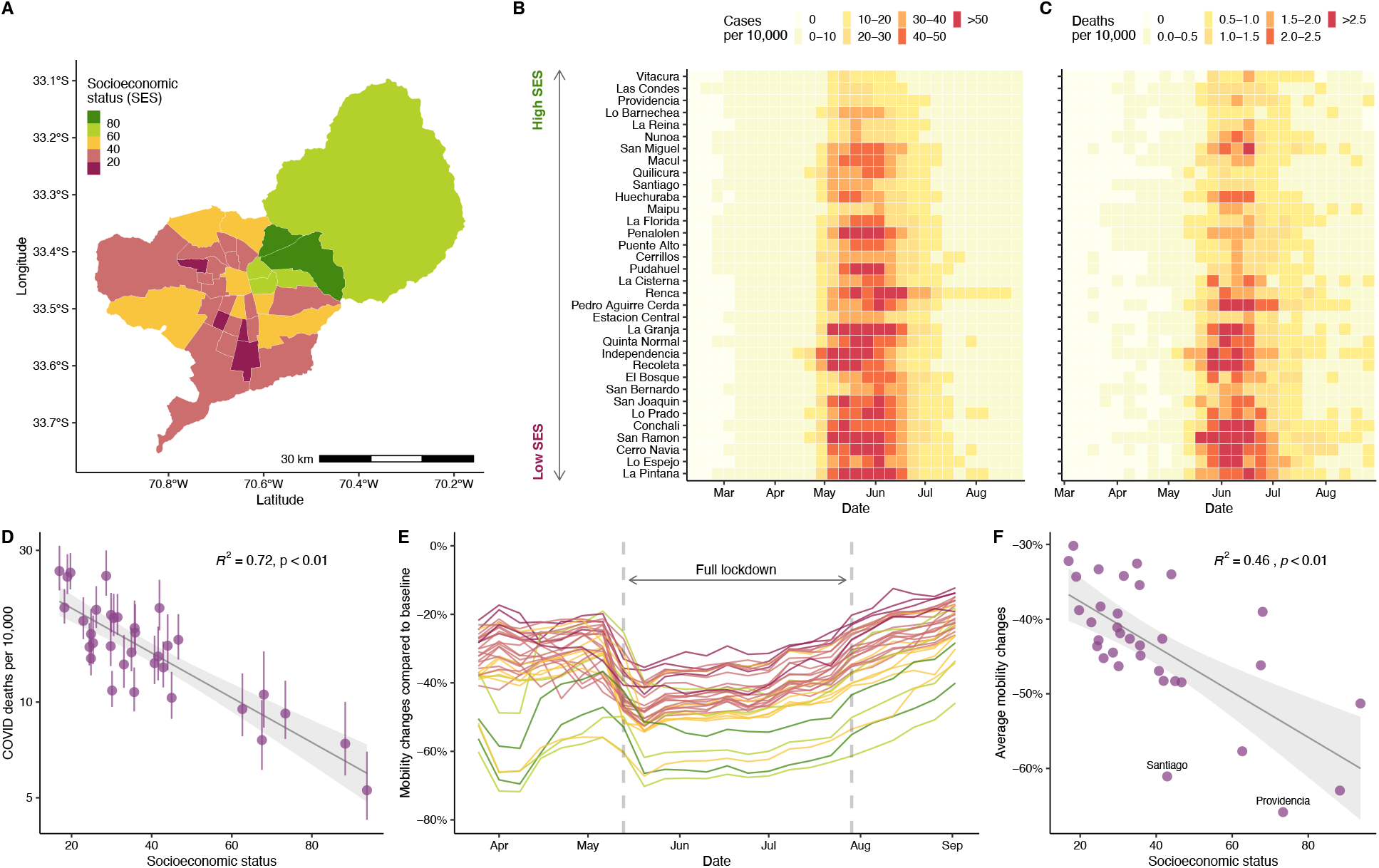
Socioeconomic status, COVID-19 cases, COVID-19 deaths, and mobility data in Greater Santiago. **A**. Municipalities that are part of the Greater Santiago are colored according to their socioeconomic status (SES). For the purpose of this analysis, we considered the Social Priority Index as a measure of socioeconomic disparity, an index that takes into account income, education, and health access. **B**. COVID-19 cases normalized by population size per municipality. Municipalities are sorted by SES starting with the one that has the highest SES at the top. **C**. COVID-19 attributed deaths normalized by population size per municipality. **D**. Demography adjusted death rates and its association with SES. The dots and the whiskers represent the median and the 95% confidence intervals respectively. The y axis is shown in logarithmic scale. **E**. Daily reduction in mobility by municipality. Each location is colored according to its SES value. Baseline values are based on 45 days previous to March 25, date that Facebook Data for Good started to generate the data for this region. **F**. Average reduction in mobility during the full lockdown period and its association with SES. The urban and the business centers, Santiago and Providencia respectively, experienced a greater reduction in mobility than expected based just on their socioeconomic profile.

Changes in human mobility –a proxy for physical distancing– during lockdown periods follow similar patterns. Using human mobility indicators, inferred from anonymized mobile phone data obtained from the Facebook Data for Good Initiative, we show that the two municipalities with highest socioeconomic status exhibited a reduction in mobility by up to 61% during the full lockdown (dark green, Fig. 1E), compared to the ones with lowest SES, which, on average, reduced their mobility to 40% during the this period (dark pink, Fig. 1E). This relationship between reductions in mobility and SES was present during all time-periods considered for this study (Fig. 1F) and is consistent with the hypothesis that people in poorer regions cannot afford to stay at home during lockdowns. Our result is consistent with analyses of New York City neighborhoods [11] and with findings from other studies conducted in Santiago that used different socioeconomic and mobility metrics [12–14].

### Excess deaths match the dynamics and demographic distribution of COVID-19 deaths

Excess deaths –defined as the difference between observed and expected deaths– can provide a measure of the actual impact of the pandemic in mortality by quantifying direct and indirect deaths related to COVID-19 [7, 8, 15]. We estimated the expected deaths for 2020 by fitting a Gaussian process model [16] to historical mortality data from the past twenty years, and used them to identify the increased mortality during to the pandemic, controlling by population growth and seasonality. As shown in Fig. 2A, the number of deaths observed between May and July 2020 is more than 1.7 times the expected value, with a peak surpassing 2110 death counts in epidemiological week 24 (first week of June) compared to an expected value of 802 deaths and an average number of deaths of 798 between 2015 and 2019.

**Figure 2:**
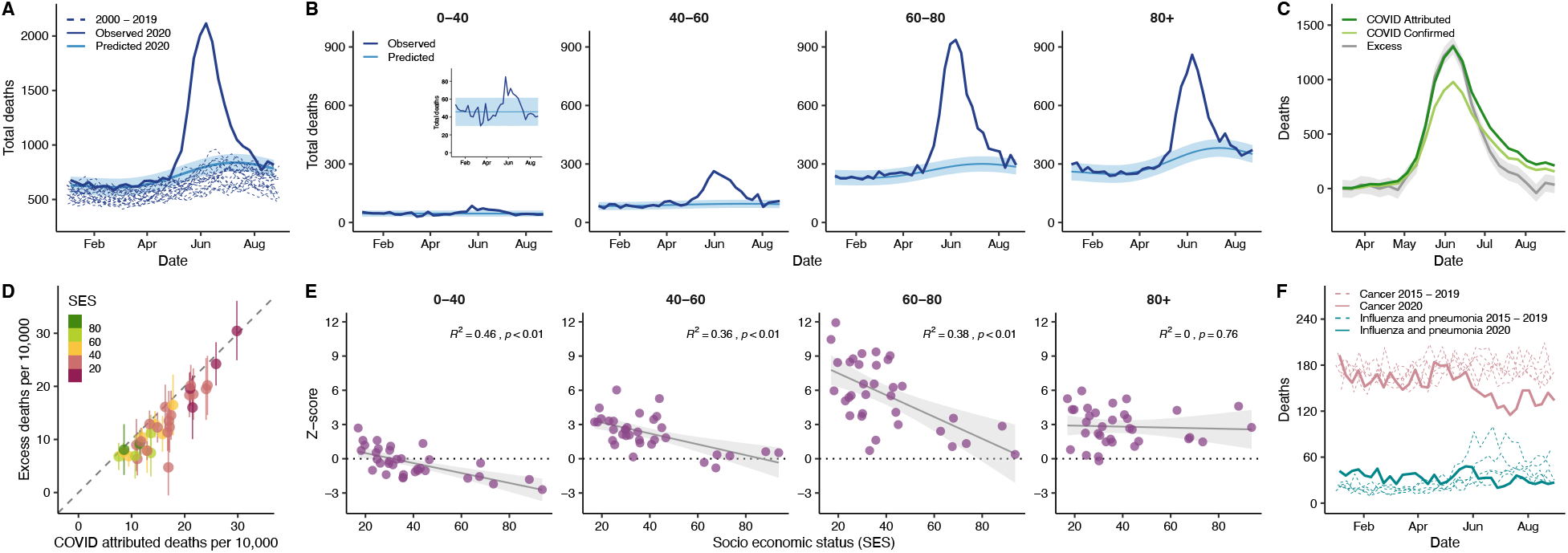
Excess deaths and its association with demographic and socioeconomic factors. **A**. Observed deaths (solid dark blue line) in Greater Santiago compared to predicted deaths for 2020 (solid light blue line and its confident intervals shaded in lighter color), using a Gaussian process regression model built with historical mortality data from 2000 to 2019 (dashed blue lines). The values contain all the possible causes of deaths. **B**. Age-specific trends of the observed deaths compared to the predicted deaths for 2020. **C**. COVID-19 deaths versus excess deaths. COVID-19 confirm deaths are shown in light green, while COVID-19 attributed deaths are shown in dark green. Excess deaths correspond to difference between observed and predicted deaths. **D**. Comparison of excess deaths and COVID-19 attributed deaths per municipality colored by SES, and normalized by population size. **E**. Monthly average of Z-scores of observed deaths between April and July by age group. The Z-scores correspond to the standard deviations over expected values. **F**. Historical deaths due to influenza and pneumonia (teal dashed lines), and cancer (pink dashed lines) compared to the observed deaths during 2020 (solid lines).

When comparing the number of deaths by age in the year 2020 with our model’s predictions we observe striking patterns. Although people younger than 40 years old have an overall lower mortality rate than those from older age groups as expected, they still exhibit a nearly two-fold increase in the total deaths with a peak in the observed deaths occurring 2 weeks earlier than for those older than 60 years old (Fig. 2B). For the age groups 40-60, 60-80, and older than 80, the observed deaths are 2.8, 3.2, and 2.4 times higher than expected, respectively. Even though the age group 80+ exhibits the highest expected mortality values for 2020, the group that contains people between 60 and 80 years old displays the highest weekly count (936 during epidemiological week 24), the biggest deviation from the predicted values, and the highest values of excess deaths (645 more deaths than expected, Fig. 2B).

COVID-19 attributed deaths for the entire Greater Santiago area fall withing the credible intervals of excess deaths until late June, when the attributed deaths increase to rates that are even higher than the excess deaths, suggesting that under-reporting in COVID-19 attributed deaths is unlikely (Fig. 2C). COVID-19 confirmed deaths –those with a PCR-confirmed SARS-CoV-2 test– follow a similar temporal pattern, and the difference between confirmed and COVID-19 attributed deaths gets smaller toward the end of August, indicative of an improved testing capacity. This pattern is consistent when compared to normalized deaths by population size for each municipality (Fig. 2D), which also shows COVID-19 attributed deaths higher than the excess deaths in most of the cases. Anomalies of 2020 across different age groups also display a significant negative association with socioeconomic status, except for the 80+ group (Fig. 2E), suggesting a higher death burden in lower SES municipalities, independent of their age composition. Furthermore, the two municipalities with SES higher than 80 (Las Condes and Vitacura) had z-scores of much smaller magnitude (with the exception of the oldest age group) indicating that there patterns of mortality did not deviate much from what would have been expected on a normal year in people younger than 80 years old.

Although the observation that COVID-19 attributed deaths are greater than the estimated excess deaths might be counterintuitive (Fig. 2D), it may indicate the presence of changes in overall mortality patterns due to other causes. Lower numbers of deaths were reported for respiratory infectious diseases such as influenza and pneumonia, and cancer during July and August of 2020 compared to the period 2015-2019 (Fig. 2F). Changes in mortality from respiratory diseases can be explained by a mild influenza season in the Southern Hemisphere during the winter of 2020 [17], which is consistent with our observation that much fewer cases of respiratory viruses have been detected in Chile during the current season (supplementary materials). A decrease in the number of cancer attributed deaths can be explained by a competing causes of death phenomenon [18], but additional analyses need to be conducted to establish this hypothesis. More detailed analyses are presented in the supplementary materials.

### Epidemic reconstruction indicates an early under-reporting in low socioeconomic areas

In order to reconstruct SARS-CoV-2 infections over time, we implemented an approach named as RmMAP that back calculates the most likely infection numbers given the temporal sequence of deaths, the onset-to-death distribution, and the demography-adjusted infection fatality rate (IFR). Figure 3A shows the outcomes of this inference process, where the reconstructions from our approach and other methods are able to capture the main peak observed in May and June, with an estimate of the number of infected individuals that is 5 to 10 times larger than the reported values.

**Figure 3:**
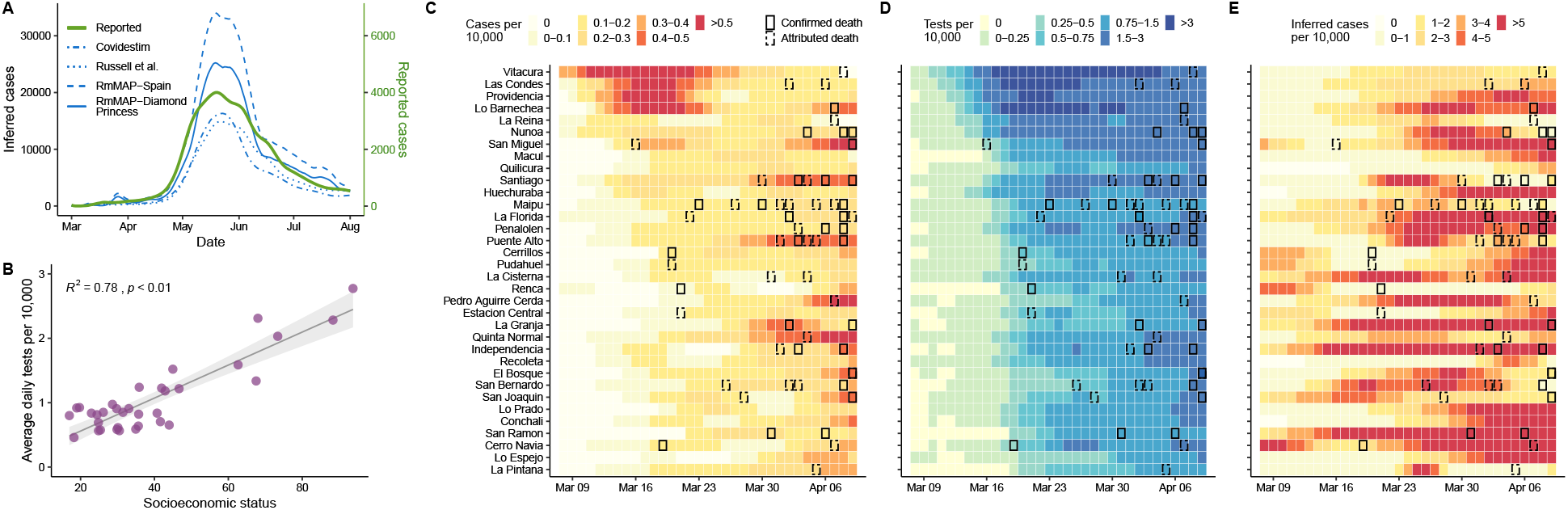
Inferred cases and tests conducted for the Santiago metro area. **A** Inferred cases by different methods (blue lines) compared to the reported cases (y axis on the right, green line). For these analyses, we considered the log-normal onset-to-death distribution described in [20] and two demography-stratified IFR estimates, one from Diamond Princess cruise ship [21] and from a seroprevalence study in Spain [22]. For comparison, we also present reconstructions based on the Covidestim method [23], and by the re-scaling of case counts by the under-reporting estimates obtained with the method of [24]. **B** Association between Average daily test and SES during the early peak. The early peak is defined as those cases reported between 03/08 and 04/09. **C** Reported cases per 10,000 by municipality during the early peak. **D** Test per 10,000 by municipality during the early peak. **E** Inferred cases obtained from the RmMAP-Spain model per 10,000 by municipality during the early peak. For panels C-F, COVID-19 confirmed and attributed cases are highlighted with solid and dashed boxes respectively.

The reconstructions also reveal important differences in the inferred number of infections during March of 2020, the month in which the virus was introduced in Chile by travellers from affluent municipalities. We analyzed the number of tests performed between March 8th and April 9th, and find a significantly higher number of tests performed in municipalities with high SES (Fig. 3B), especially during first two weeks of March (Fig. 3D). In addition, an early peak of reported cases was only observed in high SES municipalities during middle March, despite the fact that several COVID-19 deaths, which are lagged with respect to infection by up to several weeks, were reported in low SES municipalities (Fig.3C) during the same period. These findings suggest that an early first wave of infections occurred during March and quickly spread through the rest of the city without being captured by the official counts. Our RmMAP estimates at the municipality level support this claim, as they capture a high volume of early infections in most municipalities (Fig.3E), an scenario that largely deviates from the official tallies (Fig.3C). To further validate the hypothesis of an early under-reporting in low SES municipalities, and to rule that these early activity estimates are an artifact of our method, we performed experiments on a synthetic elementary model of two peaks of different sizes separated in time (supplementary materials). These experiments confirm that RmMAP is capable of recovering this bi-modal phenomena, while other methods fail to do so; they over-smooth the true signal and the earlier peak is typically not recovered. This early under-reporting signal suggests that the patterns of mortality and testing observed across the Greater Santiago are partially explained by an early failure of health-care systems in informing the population with sufficient situational awareness about the real magnitude of the threat [19].

### Wealthy areas have conducted more testing with lower waiting times

To further understand the consequences of insufficient early testing, we conducted a deeper analysis of different testing metrics at the municipality level. We first looked at testing capacity measured as weekly positivity rates, the fraction of tests that are positive for SARS-CoV-2. Our results show that the positivity signal tracked the course of the epidemic, peaking at times of highest incidence between May and July, and suggesting a highly saturated health-care system during this period across the entire city (Fig. 4A). A strong negative association between positivity and SES (Fig. 4B) further denotes difficulties in access to health care that is even more pronounced in lower SES municipalities. Despite changes in positivity rates over time, this testing metric also significantly correlated with number of cases (Fig. 4C) and number deaths (Fig. 4D).

**Figure 4:**
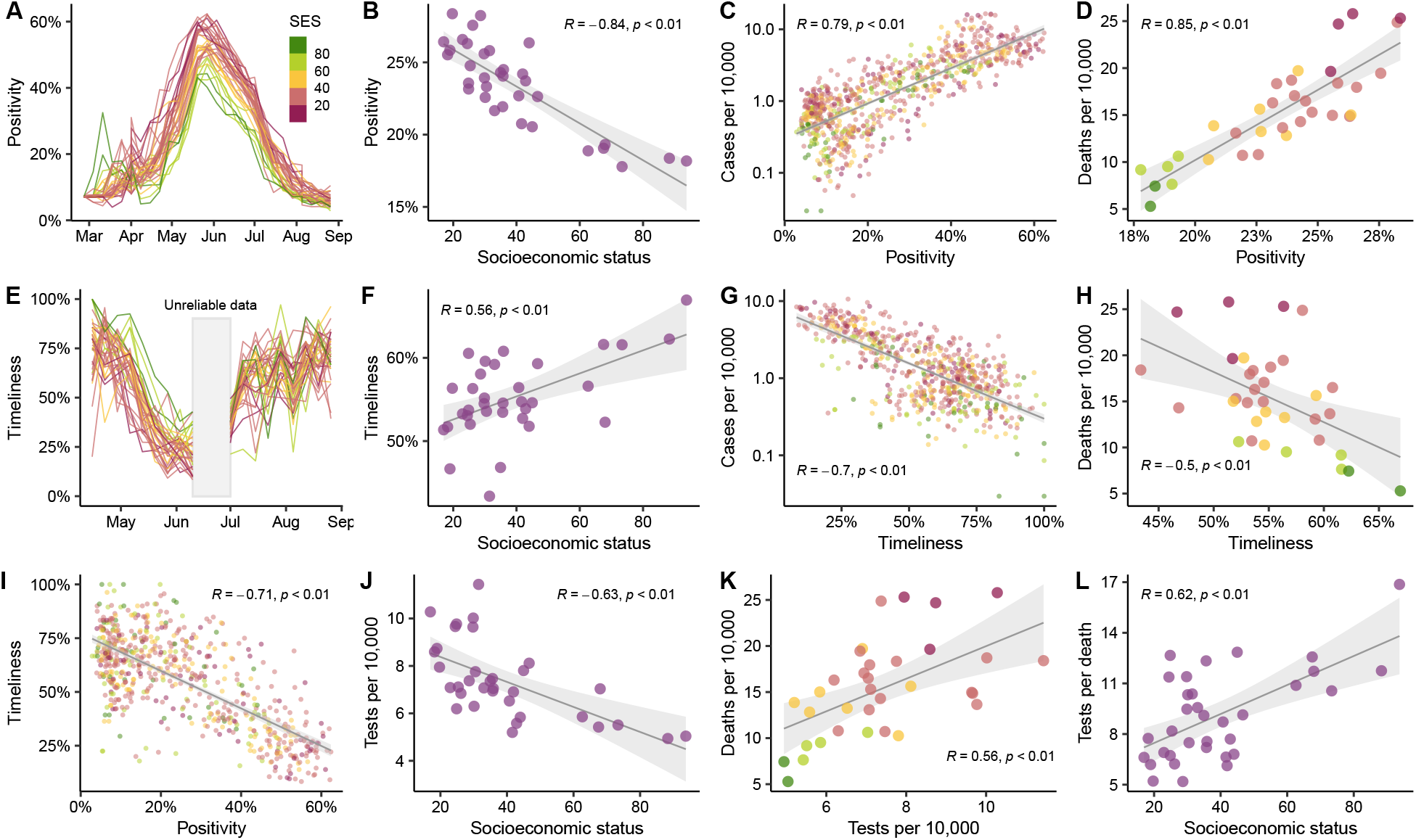
Testing capacity and timeliness. **A**. Positivity over time. Positivity is defined as the proportion of PCR tests that are positive on a given week. The municipalities are colored according to their SES value. **B**. Association between positivity and SES. Average positivity values are shown for each municipality. **C**. Association between weekly number of cases per municipality and positivity. The y-axis is in logarithmic scale. **D**. Association between the overall number of deaths per municipality and the positivity. **E**. Timeliness over time. Timeliness is defined as the proportion of PCR tests that appear in the public records within one week from the onset of symptoms. **F**. Association between timeliness and SES. Two weeks in June were excluded from the analysis due to inconsistencies in data, leading to unreliable delay estimates **G**. Association between weekly number of cases per municipality and timeliness. **H**. Association between the overall number of deaths per municipality and the timeliness. **I**. Association between timeliness and positivity. Dots are representative of weekly data per municipality. **J**. Association between tests per 10,000 and SES. **K**. Association between tests per 10,000 and deaths per 10,000. **L**. Association between tests per death individual and SES. Figures with different dot colors illustrate the SES value according to the reference presented in panel A.

We also analyzed testing capacity by estimating the delays in obtaining test results. We inferred the distribution of the delay between onset of symptoms and report of the results, from which we obtained the proportion of cases that are publicly reported within one week since the onset of symptoms or *timeliness* [25]. As shown in Fig. 4E, timeliness follows a similar temporal course as test positivity during June. This metric is also associated with SES, suggesting that municipalities with low SES, on average, get their test results later than the ones with high SES (Fig. 4F). Timeliness also correlates with number of cases (Fig. 4G), and with the number of deaths (Fig. 4H). Moreover, the negative strong correlation between timeliness and positivity (*r* =− 0.71, *p <* 0.01, Fig. 4I), suggests that they can be used to measure differences in access to health care and are strong predictors of mortality.

Our findings on the number of tests conducted show a rather paradoxical association with SES and mortality. Many months into the epidemic, the early positive association between tests per capita and SES (Fig. 3B) reversed (Fig. 4J), and tests per capita started to positively correlate with mortality (Fig. 4K). This result suggests an improvement in testing capacity over time, so that more tests were performed in more affected areas. However, when looking at tests per death, a metric that can be used as a faithful proxy of testing capacity [26], we observe a positive correlation with socioeconomic status (Fig. 4L), indicating that testing disparities persisted during the epidemic, with low SES areas being affected more. In the supplementary materials we further discuss the associations between our metrics and case counts.

### Infection fatality rate for young people is significantly higher in less affluent locations

In the absence of serological surveys, a direct inference of an infection fatality rate (IFR) is challenging. The degree of ascertainment depends on many factors, including testing capabilities and the likelihood of having symptomatic infections. The lack of age information in the reported cases at the municipality level makes this inference more challenging. To address these hurdles and to have estimates of the IFR, we implemented a hierarchical Bayesian model that considers the relationship between deaths, observed cases, and true infections across location, time, and age group.

We first estimated the case fatality rate (CFR) by assigning total cases into age groups in a simple way that projects the overall age-distribution of cases to particular municipality demographics (Fig. 5A, see supplementary materials for details). With the exception of the oldest age group, case fatality rate shows a negative association with socioeconomic status. Similarly, our resulting IFR estimates once corrected for under-ascertainment display a similar pattern (Fig. 5B) but on an order of magnitude lower than the CFRs. We then grouped the municipalities into four categories of similar sizes an label them as low, mid-low, mid-high, and high socioeconomic category. The IFR estimates from this alternative model have a similar trend (Fig. 5C), with a monotonic decrease of the IFR as a function of the socioeconomic categories, regardless of the inherent uncertainty of our estimates. When comparing the IFR ratio between the low and the high SES categories, the results show significantly higher infection fatality rate in the low SES group in people younger than 80 years old (Fig. 5D). The age groups 60-80 and 40-60 exhibit an IFR that are 1.4 and 1.6 times higher respectively in low SES municipalities, compared to the high SES ones. The difference is even more pronounced in the younger age group (0-40 years old), which shows values of IFR that is almost 3 times higher for the municipalities with the lowest socioeconomic status. Altogether, these results are in line with the analyses of excess deaths presented in Fig. 2E. A comprehensive explanation of our hierarchical Bayesian methodology, including a discussion of its assumptions and several robustness checks appear in the supplementary materials.

**Figure 5:**
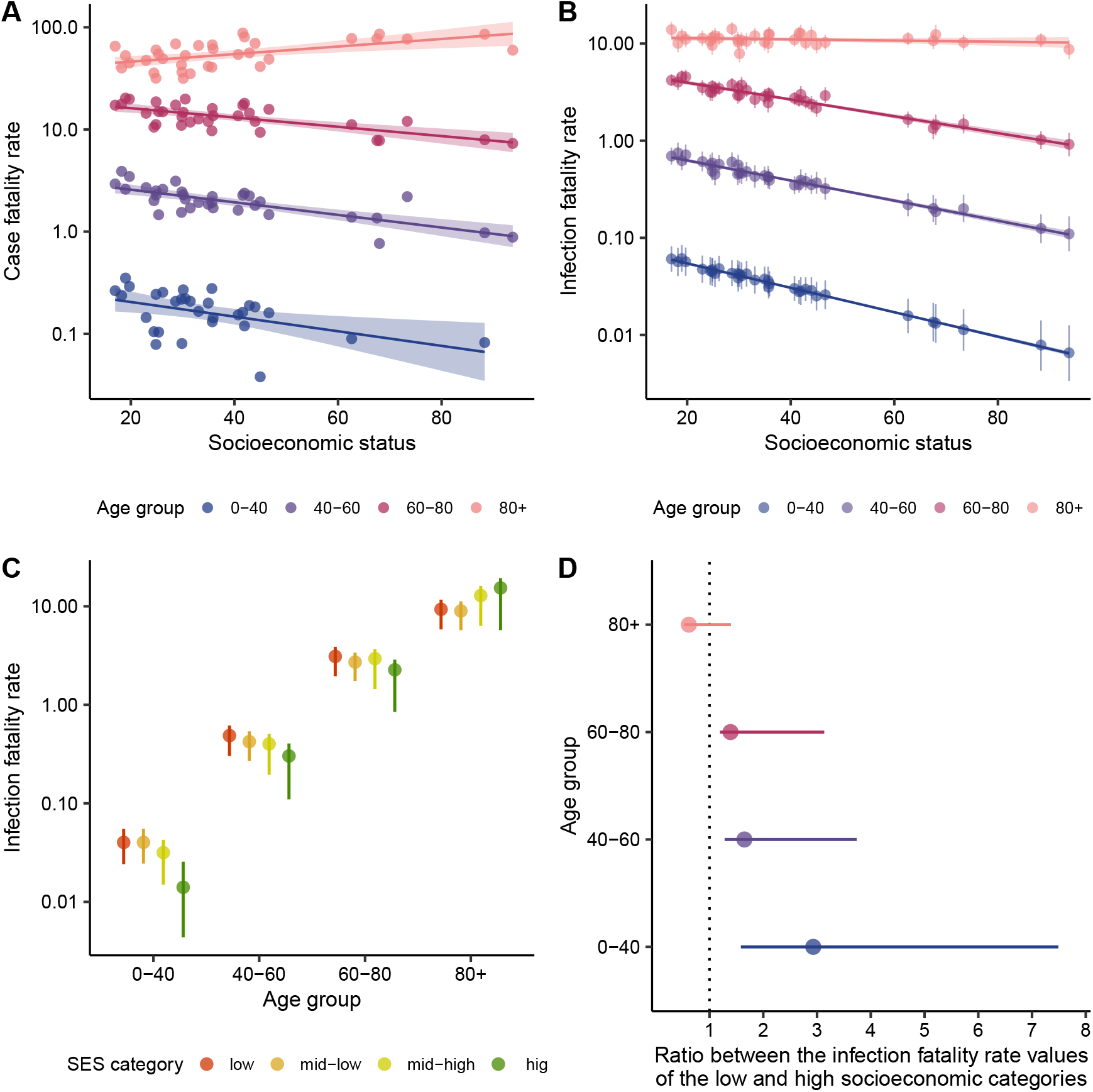
Inference of infection fatality rate by age and socioeconomic status. **A**. Estimates of case fatality rate by age and SES based on a simple assignment of cases to age groups. **B**. Inferred infection fatality rate (IFR) by age and SES using our hierarchical Bayesian model. **C**. IFR as a function of age, for four socioeconomic categories defined based on quantiles. **D**. Ratio of the low and high socioeconomic category IFR values by age group.

## Discussion

In order to disentangle the real burden of COVID-19, it is critical to consider demographic and socioeconomic factors and their consequences for the public health response. Here, we analyzed data from the capital of Chile, a highly segregated city. Our results align with the recent literature uneven health risks globally, making socially and economically deprived populations more vulnerable to the burden of epidemics [27, 28]. Mounting evidence suggests that such differences have also manifested in the context of the COVID-19 pandemic [29, 30], and since the pathways modulating these differential outcomes are yet not well-understood, comprehensive accounts are urgently needed [31], so that more resilient and socially-aware public-health strategies can be planned in advance of forthcoming pandemics. Current economic losses is forcing people into poorer conditions and likely increasing the inequality even further. In this context economic decisions can fuel new outbreaks, unless we solve the health problem first improving access and coverage to people in unequal parts of the country. In Chile, recent studies have suggested a link between SES and effectiveness of non-pharmaceutical interventions such as stay-at-home orders [12, 13, 32], and our work further explores this topic by providing an holistic perspective about how the interplay between behavioral, social, economic, and public-health aspects modulates the observed heterogeneity in infection incidence, and ultimately, in mortality.

Taken together, our results show a strong link between socioeconomic and demographic factors with incidence, mortality, and testing capacity of COVID-19 in urban settings, an association that is manifested as a reinforcing feedback loop supported by several findings. First, our analysis of human mobility indicates that municipalities with lower socioeconomic status were less compliant with stay-at-home orders, possibly because people from lower income areas are unable to work from home, leaving them at a higher disease risk. Second, anomalies in the overall excess deaths are higher in low SES areas, particularly in people younger than 80 years old, suggesting that poorer municipalities were hit the hardest. Third, our analyses revealed an early under-reporting of infections in low income areas. Since public-health measures were taken in response to nominal case counts, these places were under prepared, with a poor health care response that resulted in higher death counts. Fourth, the analyses of test positivity, timeliness, and tests per deaths indicate an insufficient deployment of resources for epidemiological surveillance. Higher positivity rates in health care centers suggest the need for greater testing and detection. At the same time, slower turnaround in test results can lead to greater potential for transmission, since even small delays between onset of symptom, testing and final isolation significantly hinder the capability of public health systems to contain the epidemic [33]. Finally, infection fatality rates were higher in lower SES municipalities, especially in young people. This can be explained either by the lack of access to quality health-care or by a higher prevalence of the comorbidities that associate with more severe presentations of the COVID-19 disease in socially deprived populations.

Along with the main findings of our work, we also introduced several methodological innovations. To our understanding, the Gaussian process modelling framework for excess deaths is new, and improves upon existing methodology [7, 8, 34, 35] by allowing a seamless and parsimonious modeling of long-term and seasonal variation in mortality. Instead of representing seasonal components as individual variables as usual, our model features a continuous representation of time where individual time variances and their covariance is represented by a suited Kernel. Additionally, the implementation of RmMAP is also new, and represents a balanced solution between the completely unregularized mMAP method [6, 36] and the expectation-maximization smoothing method (EMS) [37]. Here we have showed the usefulness of our RmMAP, since reconstructions are smooth enough to be sensible, unlike many instances of mMAP, but not too smooth so relevant underlying early activity could be missed, as with EMS (see supplementary materials for details). Our computations of timeliness and delays from publicly available data are original and extend the early analysis of [13]. Although this analysis is dependent on the idiosyncrasies of the official Chilean epidemiological surveillance system, we believe our rationale extends to other setups. Finally, our Bayesian method for joint inference of infection fatality rates and under-reporting signifies an independent contribution, and shows that it may not be necessary to have complete epidemiological datasets (here, age) to draw valid inferences, as long as the solution space is constrained enough by meaningful priors and demographic structure. Altogether, we believe that our methodological contributions are valuable for better understanding the impact of past, current, and future infectious disease dynamics.

## Data Availability

Code is available upon request. All analysis in this study are based on publicly available data.

## Acknowledgments

We would like to thank Marc Lipsitch, Oliver Stoner, Jorge Perez and Alonso Silva for insightful comments.

## Author contributions

G.E.M. and P.P.M. designed the study, conducted the analyses, and wrote the manuscript. A.S.M. and P.A.M assisted with the analyses and conceptualization. C.O.B. and M.S. designed and oversaw the study. All authors edited the manuscript.

## S1 Materials and Methods

### S1.1 Regularized Richardson-Lucy for epidemic curve estimation: the regularized mortality MAP method (RmMAP)

The Richardson-Lucy (RL) Algorithm is an expectation maximization (EM)-type scheme that since decades has been successfully applied to address the deconvolution problem in fields as medical imaging (Positron Emition Tomography, PET), and Astronomy. More recently, in has been proposed for epidemic curve estimation: for example, for the reconstruction of HIV infection curves [38] based on case incidence, and of the 1918 Influenza pandemic [36] based on mortality time series. The COVID-19 pandemic has sparked a renewed interest in this algorithm; indeed, [6] use RL for a state-wise estimation of the early pandemic course in the US, under the name of *mortality MAP* (mMAP). The idea is simple: the date of death is the sum between time of onset of symptoms and the time between onset of symptoms and death. This relation expresses as a convolution in the space of measures,

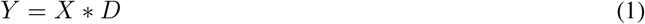

where *Y* and *X* are the distributions of deaths and infections, and *D* is the onset-to-death distribution (assumed known). Then, given a mortality series *Y*_*t*_ that ends at time *T* , we aim to reconstruct the time distribution of cases *X*_*s*_ based on our knowledge of *D* by deconvolving *X* from (1). The final series of cases *c*_*T*_ are obtained by re-scaling the obtained *X*_*t*_ by the inverse of the infection fatality rate.

In our work we propose a fix for one of the main shortcomings of RL, and in particular, of mMAP: the deconvolution problem is ill-posed, and the log-likelihood (see (3)) may possess many local optima, which often don’t correspond to a sensible solutions. Then, it is common that RL iterates progressively deteriorate and become wiggly. This phenomenon is also called noise-amplification and has been extensively discussed in the signal processing literature [39, 40].

The remainder of this section is organized as follow: in S1.1.1 we describe in detail the RL algorithm. Then, in S1.1.2 we describe a simple type of regularized RL method, the basis of our *RmMAP*. We conclude by discussing in S1.1.3 on the role of regularization and the relation between mMAP, RmMAP, and an alternative S1.1.3.

#### S1.1.1 Richardson-Lucy Algorithm

Suppose counts *Y*_*t*_ of observations on a *projection space t* (*t* typically represents bins over time, space, etc) have a Poisson distribution where the parameter depends linearly on a latent process *X*_*s*_, through the linear operator *A* = (*a*_*t,s*_). Thus, we write the conditional distribution of the observed counts given the latent *X* and *A* as

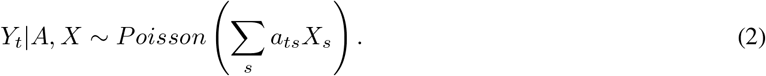

The associated log-likelihood writes as

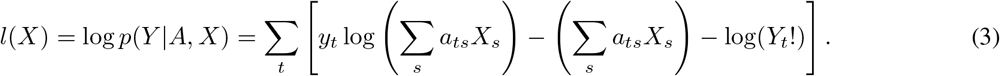

Here, we restrict our treatment to the case where *A* is of the form *a*_*t,s*_ = *P* (*t*|*s*) for some kernel of probabilities *P* . In this case we can interpret the generative model 2 as the one where particles are emitted on the *s* space with intensities *X*_*s*_. Each of them has a probability *a*_*t,s*_ to appear on *t* space. Therefore, the observer receives *Y*_*t*_ that has the intensity *n*_*t*_ = Σ _*s*_ *a*_*ts*_*X*_*s*_; it is the superposition of the processes with intensities *a*_*ts*_*X*_*s*_ [41]. The deconvolution problem is the one of recovering *X* given observed *Y* and known *P* .

Additionally, notice that conditioning on the total number observations we can also understand (2) as a *denoising* problem: consider a latent density *q*_*s*_ = Σ*n*_*s*_*/ n*_*s*_ and an observed histogram *r*_*t*_ = Σ*n*_*t*_*/ n*_*t*_. This histogram *r*_*t*_ is obtained through a simple sampling mechanism: first, observe *Q ∼ q*, and then, corrupt this variable with a noise distributed according to *P* (*·*|*Q*) to obtain *R ∼ r*. Then, solving (3) amounts to finding a *clean* version of the histogram *r*.

The Richardson-Lucy algorithm can be applied to find such estimate *X* in (2). It is simply an EM algorithm [41, 42] that iterates between computing the conditional expectations of intensities *a*_*ts*_*X*_*s*_ (E step) and maximizing the resulting expected log likelihood of (2) with respect to these (M step). These two may be written as [43]

- E-step. Compute the conditional expectations *Z*_*ts*_ of *a*_*ts*_*X*_*s*_ given current parameter estimates and Data

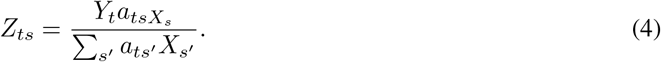
- M-step. Maximize the expected (with respect to *Z*_*ts*_) log-likelihood (3)

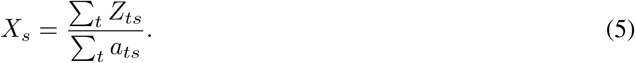

Notice the denominator in (5) accounts for the fact that *a*_*ts*_ may not add up to one. This will be critical in our applications (see below). When combined, the E and M steps (4)-(5) give rise to the Richardson-Lucy algorithm, which simply states as

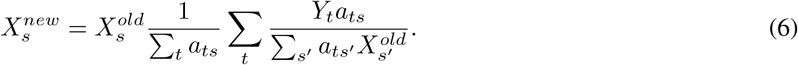

RL iterations are seamlessly implemented through elementary linear algebra operations.

As mentioned before, this method can be readily applied for reconstruction of epidemic curves based on mortality data. Interestingly, in this case the denominator in (5) is simply the probability that someone will die before *T* , having got sick at time *s*. Therefore, RL automatically accounts for right truncation (although in practice performance typically degrades at the end of the recording period).

#### S1.1.2 Regularized mortality map (RmMAP)

Here propose to use a simple and scalable modification of (6), originally presented in [39] in the context of PET imaging. The idea is to add a quadratic penalty to *l*(*X*), so we now maximize 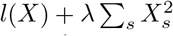 This leads to a small change of the M-step, in turn, leading to the following modified Richardson-Lucy scheme

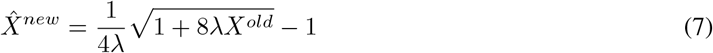

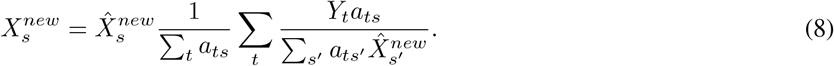

One important question is how to choose the regularization parameter *λ*. Although this is largely an open question, we have observed (see S1.1.3) that one sensible approach is to consider a grid of possible values (here, *λ ∈* [10^−10^, 10^2^]) from which we select the best. For choosing the best, we consider two criteria that have to be simultaneously satisfied: first, curves should be smooth enough, and as a measure of smoothness we take Σ|*X*_*s*+1_− *X*_*s*_|. Also, curves should provide a good fit to data, and we use the *χ*^2^ goodness of fit statistic described in [36]. We have observed that curves that don’t satisfy this criterion are typically the ones that appear too smooth. In summary, we choose *λ* such that the associated smoothness measure is the smallest among the ones for which the goodness of fit criterion is satisfied. Alternatively, we may choose *α* by cross validation, as it has been suggested recently [44].

We call *regularized mortality MAP, RmMAP* the method that consists on reconstructing epidemiological curves based on mortality data by appealing the procedure just described.

#### S1.1.3 Experiments and discussion

##### Effect of initialization and amount of regularization

we start by showing, in Fig. S1 the effect of regularization, and of random initialization. We observe that random initialization leads to more irregular curves. Nonetheless, we also observe that sensible results can also be obtained by averaging the resulting RmMAP solutions from many seeds (not shown). In general, the obtained solution will be a compromise between not being excessively wiggly (black curves, no regularization) or too smooth (pink curves).

##### Robustness to changes in the onset-to-death distribution

in Fig S2 we show reconstructions using two log-normal onset-to-death distributions from [20]. One of them is the one used in the analysis of the main text, with mean 14.5 and median 13.2 days. The second accounts for right truncation and has mean 20.2 and median 17.1 days. We observe that reconstructions don’t change substantially from one reconstruction to another, and moreover, that the difference reconstructions is different than a shift of 5 − 7 days.

##### Comparison with other methods

in Fig S3 we show results for an alternative method, based on expectation maximization smoothing (EMS). EMS was first described in [37] and applied in HIV epidemiology in [38]. EMS iterations are very similar to RL’s, but the M step (5) is further complemented by a smoothing (S) step where the current curve *X*^*new*^ is filtered by convolution with a bounded support filter that smooths out possible spurious noise (here, we consider a triangular kernel). We observe solutions given by EMS are similar to the ones by RmMAP, but they are smoother for all values of *k*.

##### Supplemental reconstructions

we first show in Figs S4 and S5 a comparison of the reconstructions given by RmMAP and EMS in the same setup of the main text. Whether RmMAP or EMS provide the most sensible and whether earlier peaks suggested by RmMAP are *bona fide* or rather reconstruction artifacts is an open question whose full answer ideally would make use of additional sources of information.

##### Experiments with synthetic data

Nonetheless, we believe it might be the case that EMS tends to oversmooth, and RmMAP estimates are more realistic. Our claim is supported by Figs. S6 and S7, where we consider the simplest possible hypothetical scenario of two peaks: a total of *n* = 10, 000 cases occur at exactly one of two possible dates; the first of these events occurs around March 10th, 2020 (*t*_0_), and occupies a proportion of *w <* 0.5 of the total number of cases. The second event follows *h* days after. We observe that in most of the situations EMS tends to smooth-out the earliest peak (mostly where *w* or *h* are small), but RmMAP tends to recover a most sensible (although not perfect) solution that at least represents this bi-modal behavior. In Fig. S8 we show additional reconstructions with the Covidestim method [23]; they also tend to oversmooth the first peak.

### S1.2 Gaussian Processes for excess mortality estimation

We performed Gaussian Processes (GP) regression [16] to predict mortality in 2020 and report confidence intervals. GPs can be understood as an infinite dimensional Bayesian regression: indeed, in usual finite Bayesian regression settings one fits *y*_*i*_ = Σ_*i*_ *w*_*i*_*x*_*i*_ + _*i*_ where _*i*_ are symmetric Gaussian i.i.d errors, *x*_*i*_ are given covariates and *w*_*i*_ the coefficients, sampled from a prior distribution *p*(*w*). In contrast, when using GPs one fits *y*_*i*_ = *f* (*x*_*i*_) + _*i*_ where *f* is a function sampled from a prior over functions *p*(*f*). GPs are appealing because two reasons: first, as a non-parametric Bayesian method, the level of complexity is automatically adjusted by the complexity of data. Second, they are computationally tractable,regardless of their infinite dimensional structure.

Priors over *f* are specified through a *kernel K*, which is supposed to encode the correlational structure of data. Indeed, *K*(*x, x*′) is simply the *prior* covariance between *f* (*x*) and *f* (*x*′). This kernel may depend on a finite number of unknowns *θ* (i.e. *K* = *K*_*θ*_) and a main step to fitting a GP regression model is performing inference over such unknowns.

In our setup, we used a GP to account for both long-term trends in mortality as well as seasonality. To do so, we follow the same setup as in [16, Chapter 5]: specifically, we consider a kernels of the form

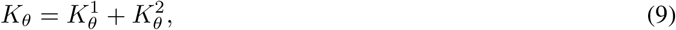

where 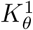 is an exponential kernel representing the long-term variation, and is given by

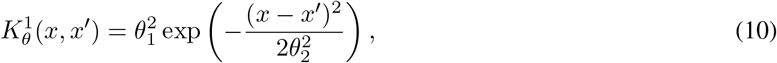

and 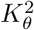 is a periodic times exponential kernel representing seasonal variation

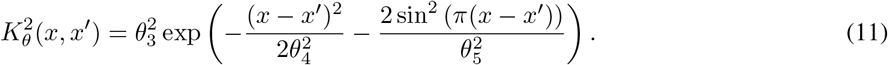

Finally, we considered a secondary source of unstructured randomness through the term ϵ_*i∼*_ 𝒩 (0, *σ*^2^), whose parameter is also fitted. We notice that although it would be more realistic to consider a Poisson or Negative Binomial link function instead of this Gaussian error, in practice our model provides a good fit to the data, so we keep this more elementary Gaussian treatment.

We performed Bayesian inference (MCMC) over the parameters (*θ, σ*^2^) which are assumed to come from usual prior distributions with parameters compatible with the scales of the process. Specifically, the length-scale parameters *θ*_2_, *θ*_4_, *θ*_5_ were drawn from

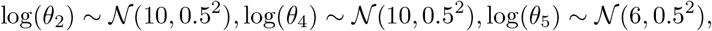

the variance (strength) parameters *θ*_1_, *θ*_3_ were drawn from

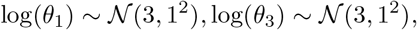

and finally, the unstructured noise term *σ* was also drawn from

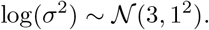

In all the above specifications time is represented in days. We implemented our inferences through the R package *Greta*.

#### S1.2.1 Details

We fitted Gaussian processes for daily all-cause mortality data available since 2000 from the *Department of Vital Statistics DEIS*). These weekly report contain anonymized individual death data including date of death. We only considered data up to late August so correcting for reporting delays was not necessary.

We performed separate analysis for each combination of age group (0-40, 40-60, 60-80 and 80+) and each urban municipality, and we also performed analysis for the entire population (all ages) and the entire Greater Santiago (the set of urban municipalities). To ensure parameters could be identified, we considered monthly series for all municipality-specific analysis and weekly series for the analysis of the entire Greater Santiago. To enhance tractability we used the inducing point method [45] with *n* = 70 inducing point for weekly series and *n* = 50 for monthly series.

We used data from 2000-2019 to fit the model and learn the posterior distribution of (*θ, γ*) sample estimates for 2020 using the usual GP prediction formulae [16] and the inferred posterior. We considered 5 chains and 10,000 samples per chain, with 10,000 burn-in samples as well.

We visually inspected resulting datasets in order to ensure convergence. On a couple of cases, implausible estimates were observed in chains with unusually low variance. We discarded such chains and re-run the algorithm. In table S1 we report an elementary posterior predictive check [46] that suggest inferences are correct. This check consist on observing the proportion of cases (between 2000 and 2019) that data falls below or above the predicted lower or upper limits of the 95% credible interval. This number is typically very close to 95%.

### S1.3 Delay estimation

Here we describe our methodology for delay and timeliness estimation. Our analysis is based on reports by the epidemiological surveillance system *[EPIVIGILA](https://www.minsal.cl/nuevo-coronavirus-2019-ncov/informe-epidemiologico-covid-19/)*, from the Ministry of Health. These were scrapped, and made digitally available by the [MinistryofScience](https://github.com/MinCiencia/Datos-COVID19). These reports have the structure described in Fig S9: they provide the number of cases whose onset of symptom started at a given epidemiological week, for each municipality. Given they are published twice a week (typically Monday and Friday) we were able to profit from the history of such reports to create a reporting triangle [25] from which we estimate delays and timeliness.

More specifically, given a certain report *R* we can estimate the (empirical) probability *P* (*D*_*r*_ ≤ *t*), that the *retrospective delay* is smaller than *t* days. Here, *t* = 0, 7, 14, … , *T* or *t* = 3, 10, 17, … *T* , depending on the day of the week of the report (Monday or Friday, respectively). by just looking at how cases have been updated. In this way, our *timeliness* estimate is simply *P* (*D*_*r*_ ≤ 7), which is obtained from Monday’s reports. Likewise, for a given epidemiological week *W* we can estimate the empirical distribution of the *prospective delay P* (*D*_*p*_ ≤ *t*), for *t* = 0, 3, 7, 10, 14, … , *T* by looking how cases for that week are eventually updated in subsequent reports. We notice a similar analysis has been previously described in [47]. At the time of developing this method we were unaware of this work.

We chose the maximum delay at *T* = 50 since we observed delays are typically much smaller. We only report delays on the period encompassing April 17th, 2020 (the first date for which this analysis is possible) and ending June 15th, 2020. We chose this date because after that reporting frequency changed arbitrarily, and reporting became unreliable for the purposes of delay estimation.

In order to make our analysis more robust we grouped our empirical probabilities *P* (*D*_*r*_ ≤ *t*) and *P* (*D*_*p*_ ≤ *t*) in windows encompassing two weeks. In that way we are able to represent randomness in our estimates: we choose *n* = 5, 000 ‘bootstrap samples’, and since windows may contain several *P* (*D*_*r*_ ≤ *t*) or *P* (*D*_*p*_ ≤ *t*) for the same *t*, at each time we randomly sample *P* (*D*_*r*_ ≤ *t*) and *P* (*D*_*p*_ ≤ *t*) from their possible values. The outcome of this procedure is sequence of *n* bootstrap cdfs, where to avoid the problem that these ‘pasted’ pseudo-cdfs may not be increasing, we hace forced them to be increasing by casting a suitable isotonic regression problem [48].

Additionally, to obtain cdfs defined on the entire temporal daily scale, we further decomposed *P* (*D*_*r*_ ≤ *t*) and *P* (*D*_*p*_ ≤ *t*) on sequences *P* (*D*_*r*_ = ≤ *t*), *P* (*D*_*p*_ = ≤ *t*) by simply sampling uniformly the remaining probabilities from the probability simplex (that is, from a Dirichlet distribution with uniform parameter *α* = (1, … , 1)). This procedure can be understood as simply randomly imputing the day of report or onset of symptom, given the inherent lack of knowledge since reports are bi-weekly, and cases are available only at the resolution of weeks.

As a result, for each period time interval we obtain a the sequences 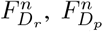 of bootstrap cdfs from which we can compute expectations, variances, etc, using the usual moment formulae. These moment estimates are naturally endowed by variance estimates coming from randomness in the *n* estimates.

### S1.4 Infection fatality rate estimation

For the inference of IFR we adhered to the hierarchical Bayesian framework, and deployed a joint model for reporting rates (and hence, IFR) based on reported cases and tests over time (*t*, month) and municipality (*m*), and deaths per age group (*a* taking values 0 − 40,40−60,60−80 and 80+) and municipality (collapsing over the temporal dimension). We used the age-stratified demographics to cap the possible number of cases and deaths through a cascade of binomial models. The main appeal of this framework, is that although most of the components are not identifiable (e.g. if reporting rates and true cases are both unknown, the same observed case counts can be achieved by multiplying both by the same factor) [49], we can borrow from better known quantities (e.g. rough estimates of prevalence, reporting, etc) to enhance identification while propagating the appropriate levels of uncertainty over the several parameters.

Specifically, the reporting rate *r*_*m,t*_ links to the observed positivity rates *pos*_*m,t*_ (in log-scale) through a logistic-linear relation (with parameters *β*), and we have also included random effects _*m,t*_ to represent unobserved causes of reporting

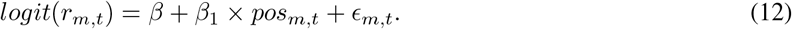

These are ultimately multiplied by age specific factors *ρ*_*a*_ *∈* [0, 1] so that the final reporting rate by age, municipality and time is *r*_*m,t,a*_ = *ρ*_*a*_ *× r*_*m,t*_.

Total infections by municipality and age 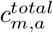 are a fraction *p*_*m*_ of the total population *P*_*m,a*_, i.e.

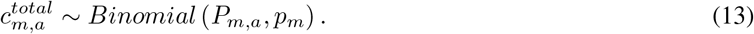

Notice that a implicit assumption is that attack rates by are proportional to the demographics of each municipality. Moreover, we assumed the following relation for *p*_*m*_

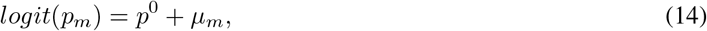

Where *p*^0^ represents a baseline (on the logit scale) of the proportion infected and *µ*_*m*_ is a municipality-specific random effect, sampled from a certain distribution.

These total cases spread across time through certain parameters *γ*_*m,t*_ *∈* [0, 1], *Σ*_*t*_ *γ*_*m,t*_ = 1 so that 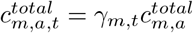.

By marginalizing over age (and thus re-weighting the age-specific reporting rates by their relative importance) we compute an effective reporting rate *r*^*effective*^ and use it to relate total case counts 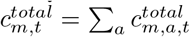 with the reported cases over time and municipality 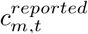.

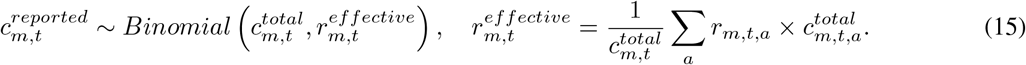

Additionally, infection fatality rates *IFR*_*m,a*_ relate to total cases 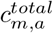 and total deaths *d*_*m,a*_ through another binomial model

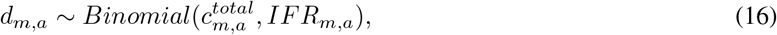

where the *IFR*s follow a stratified logistic-linear relation with socioeconomic status (SES) and age mediated by parameters *α, η, δ*:

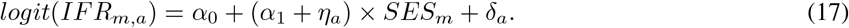

We finalize the definition of our model by specifying priors for the remaining parameters. In all cases we consider Gaussian distributions and apply suitable logistic transformation to enforce them to belong to certain ranges as appropriate. Specifically, by denoting *T* (*a, b*) the truncation operator over the interval [*a, b*] we defined

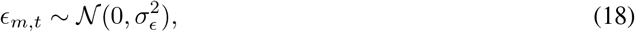

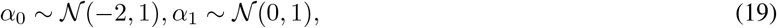

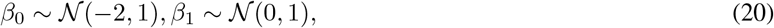

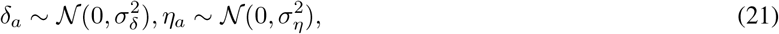

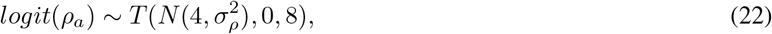

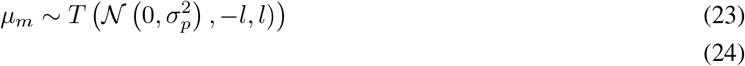

We chose the prior means for *α*_0_, *β*_0_ so that they would produce reasonable ascertainment and IFR rates, and the variances of *α*_*i*_, *β*_*i*_ arbitrarily equal to one (we did not observe sensitivity to such choice). For the remaining variances of the ‘random-effect’-like quantities we used truncated Gaussian priors, that is, all of 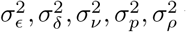 were sampled from a *T* (*N* (0, 1), 0, + ∞) distribution. The (logit) prior for *ρ*_*a*_ was chosen in the range [0, 8] interval so that that age-specific detection rates lied on reasonable ranges, and so that bad-local-optima pathologies would be avoided.

We observed that specifications of the distribution of *p*_*m*_ had a large impact on inferences, and that in the absence of any control over *p*^0^ and *µ*_*m*_, the resulting posteriors over *p*_*m*_ could oscillate too wildly and be unrealistic (e.g. above 50% of infected in some municipalities). Because of this, we treated *p*^0^ and *l* (the maximum possible value for each *µ*_*n*_, as described in (23)) as fixed external parameters and studied the effect of this value over inference, by pooling (ensembling) the predictions of each model.

For the priors over *γ*, for each municipality we used a set of *T* log-Gaussian priors *κ*_*m,t*_ (one for each month) and converted them into an element of the simplex by taking a logistic transformation, followed by normalization. This is an alternative to the usual Dirichlet distribution, that we found to perform better in practice. Specifically,

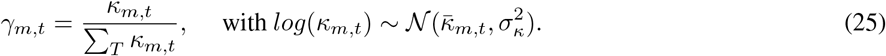

We chose the mean parameters 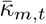 so that in the absence fo noise the resulting *γ*_*m,t*_ would match the temporal distribution of cases given by the Covidestim deconvolutions (see Fig. S33). This provides a strong signal that enables the identifications of the sub-reporting parameters through the discrepancy between inferred and reported cases at each municipality and time.

In summary, the definition of our model is made up by equations (12)-(25) and all descriptions above. In particular, our model is specified by

1. A logistic-linear relation for the IFR as a function of SES and age, equation (17).
2. A prior model for the profiles of infections as a function of time, equation (25)
3. Fixed values of *p*^0^, *l* that modulate the possible values of proportion infected

#### S1.4.1 Variations

To study robustness of our results to the choice of the model, we considered several variations over the above specifications 1-2. Specifically, we considered two variations over the functional relation of IFR: first, we considered two alternative models for the IFR as alternatives to equation (17). In the first of them, we added a municipality-specific random effect *υ*_*m*_ so we can contrast the inferences of perhaps too simple (17) with this new saturated model. In the second, we replaced the linear relation (17) by the categorical relation

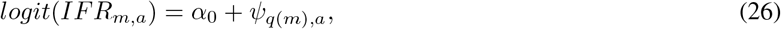

where *q*(*m*) *∈* {1, … ,4} indicates the *quartile* of municipality *m* according to its SES; low (1), middle-low (2), middle-high (3), or high (4). One appeal of this model is that, for each age group *a*, we can directly compute IFR ratios of two quartiles *q*_1_, *q*_2_ as

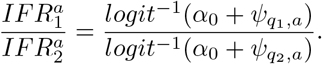

Indeed, this is the computation shown in Fig. 5D of the main text.

Additionally, we studied robustness to specification of the temporal profiles in infections, equation (25). Specifically, in addition to the official specification given by Covidestim, we also used as priors, for each municipality, the inferences given by our *RmMAP* (see Fig. S33).

#### S1.4.2 Sampling details

We considered the *T* = 6 months between March and August. For simplicity, we have ignored right-truncation effects in the computation of IFRs, but because of the apparent decreases of cases and deaths after July, we believe the error is negligible. For inference we used the R package *Nimble* [50], a wrapper of *JAGS* [51]. We considered all possible values *p*^0^ *∈* {−1.5, −1.25, −1.0, −0.75, −0.5, −0.25, 0} (7 possible values) and *l ∈* {0, 0.1, 0.2, 0.3, 0.4, 0.5, 0.6, 0.7, 0.8, 0.9, 1.0, 1.1, 1.2, 1.3, 1.4, 1.5} (16 possible values), and for each of them

two different random seeds and three arbitrary different initial conditions for the parameters to ensure the parameter space was well explored. We chose such values of *p*^0^ as they would be consistent to overall estimates of proportion infected in Chile, around 25% for a comparable period [9]. Considering the 6 = 2×3 variations described above, this gives a total of around *K* = (7 × 16 × 2 × 4) × (2× 3) ≈ 5300 possible configurations, and we refer to each of them as a separate model. For each of these models we obtained posterior draws using two chains, each with 100,000 burn-in samples and 20,000, samples which where thinned with an interval of 40.

#### S1.4.3 Model averaging and checking

We combined the resulting models by appealing to the usual Bayesian model averaging (BMA) framework [52]. Specifically, let *Y* denote the data, Δ be a quantity of interest (e.g. an IFR) and ℳ_*k*_ a model. We averaged the models posteriors according to the formula

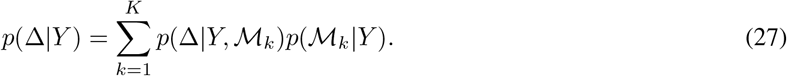

The posterior likelihood of the *k*-th model, *w*_*k*_ = *p*(ℳ_*k*_ |*Y*) can be hard to compute, as it necessitates the calculus of the marginal likelihood of the data given that model. Here we follow a pseudo-BMA strategy [53], and instead of explicitly computing the weights *w*_*k*_ we approximate them with a suited model evaluation quantity. We used the WAIC (widely applicable or Watanabe-Akaike information criterion) [54] because it is readily available in the *Nimble* framework. Specifically, we used the following approximation for the weights (under the convention that WAIC is defined as a deviance-like quantity, i.e. with a −2 factor and with lower values of WAIC indicating a better fit):

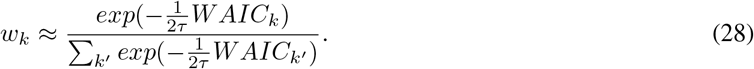

Notice that we have included a temperature parameter *τ* . The main goal of this simple heuristic is to include an appropriate number of models in the average, as we preliminary observed that choosing *τ* = 1 (the original prescription) leads to the average of only two models; i.e. *w*_*k*_ = 0 in the vast majority of cases. Although our official estimates always correspond to *τ* = 1, by considering a suitable value of *τ* we can explore how our results are sensible to the choice of better or worse models. Notice that in the limit of *τ* =*∞* we give an equal weight to each model, and in the limit of *τ* = 0 we only select the model with the lowest WAIC. Therefore, by letting *τ* move freely we are able to explore the continuum between ‘only the best model is selected’ and ‘all the models are given the same weight’, with the official pseuso-Bayesian averaging criterion being an intermediate point.

In terms of Bayesian modeling, our heuristic is justified by the necessity to accounting for the facts that Bayesian averaging methods i) are not valid in the ℳ −open setup (i.e. when the models are misspecified, as they are inevitably in our case), and ii) they ignore uncertainty over the data generating distribution, which is different from the observed. We defer the reader to [53] for somewhat more involved alternatives for better Bayesian model averaging.

#### S1.4.4 Experiments and discussion

**Model averaging with** *τ* Fig. S34A illustrates the heterogeneity in WAIC in our data, while Fig. S34A depicts the role of *τ* (equation 28) for model averaging; if *τ* is too small only one model is taken into account, and if *τ* is too large then the mixture is uniform over models.

#### Role of the prior over *γ* on ascertainment inference

In S38A we show that the averaged models infer a negative relation between ascertainment rates and positivity. Additionally, in S38B we observe that Covidestim predictions lead to negative coefficients for the under-reporting rate, while the ones based on *RmMAP* are positive. This implies that models with Covidestim as prior are may perform better and weight more in the mixture of models. If we assume the most natural interpretation that positivity and ascertainment rates should be negatively correlated, we conclude that Covidestim reconstructions are more faithful for this analysis. This might be a consequence of the fact that i) Covidestim takes positivity rates as an input so data might be implicitly being used twice, and ii)*RmMAP* reconstructions tend to spread the mass too uniformly over the months that encompassed the peak (around May-July, see Fig. S33). This observation suggests a direction for further improvements in RmMAP reconstructions, but we emphasize that the validity of our previous analysis (Fig. 3 in the main text) have to do with a very localized setup (the early peaks in March), and therefore it is not compromised by these findings.

#### Posterior predictive checks

As sanity checks we studied the extent to which our fitted models were able to produce data similar to the one observed. In Fig. S37 we show predicted and observed cases (as in [25]) and deaths for our mixture of models. We note that similar results are obtained for individual models as well (not shown), and for mixtures. but also include deaths and showed that that fitted model is compatible with the observations in that the credible intervals of key quantities cover the true values in the vast majority (if not all) of cases. Additionally, in Fig. S37C we show the median of the random effects _*m,t*_ and observed they are typically close to zero.

## S2 Supplementary text

### S2.1 Socio-economic status

In our sociodemographic analyses we considered as covariate socio-economic status (SES). We defined this number as *SES* = 100 − *SPI*, where SPI is the *social priority index* (indice de prioridad social, in Spanish) [55] a composite index (between 0 and 100) that has been developed by the Ministry of Social Development (Ministerio de Desarrollo Social, in Spanish) for the Region Metropolitana since 1995. The SPI is an actionable indicator; it equally weights the three dimensions of income, and access/quality of education and healthcare to produce a ranking in a way that a high SPI indicates that a municipality is of priority for the social programs of regional government because of its lack of resources. Therefore, our definition of SES is consistent with the fact that a relatively low SES indicates that a municipality is at an disadvantaged condition with respect to higher SES’ ones.

### S2.2 Viral surveillance

We scrapped the official records of viral surveillance from the institute for public health (ISP) (see for example, [56]) and tabulated total number of test performed and positives in Region Metropolitana, both in the entire population and over the hospitalized subpopulation. In Figs. S13 and S14 we show descriptive summaries of viral circulation over time. These made clear that 2020 has been unusual in terms of both cases and tests.

### S2.3 Mortality data reliability

For our excess mortality analysis we looked for additional confirmation to ensure our data was reliable. First, in Table S3 we show total numbers of deaths in the *Greater Santiago* up to August 30th, as reported in four different subsequent reports. There is little variation, suggesting that the reports have stabilised and our analysis is not corrupted by reporting delays.

Specifically, in Fig. S15 we show monthly reported deaths according to DEIS (our official data for analysis) and the Civil Registry. We observe these quantities are historically close to one another, and this agreement also holds for this year, suggesting that DEIS data is reliable if Civil Registry data is reliable. We further confirmed the reliability of the Civil Registry data by looking at historical birth series in Greater Santiago, also provided by the Civil Registry (see Fig. S16). We do not observe gross changes in birth recording, indicating reliability. Notice we did not use Civil Registry data for our analysis as deaths are tabulated by the municipality of death, and not of residence.

### S2.4 Causes of death

To better understand the seeming COVID-19 death over-reporting (Fig. 2C-D in the main text) we analyzed trends of individual causes of mortality. In Fig. S17 we show the total deaths (between March and August) from the 16 most significant causes of death. Additionally, in Fig. S18 we show the four more dominant causes; circulatory, respiratory, digestive and cancer in all age groups. These all exhibit a somewhat unusual pattern for the 2020 calendar year, with seemingly fewer deaths than expected in the oldest age groups. Further, in Figs. S19, S20 S21 and S22 we show trends for individual sub-causes for each of these grand-causes, and in Figs S23 and S24 we show deaths at the weekly resolution for some sub-causes of death of respiratory and circulatory causes, so seasonality can be looked at in more detail. Altogether, these Figures help explain the over-reporting phenomenon: first, fewer recorded deaths by cancer in 2020 could be the result COVID-19 appearing as the cause of death in some cancer patients. second, the lack of a peak for deaths by respiratory causes in 2020 (Fig. S23) suggests that either some seasonal respiratory deaths could have been misatributed as COVID-19 or by a truly lower number of respiratory deaths in 2020. Our data on viral surveillance supports the later hypothesis, as lower deaths by Influenza and Pneumonia coincide with lower numbers of viral circulation (Figs. S13 and S14). However, the hypothesis that surveillance systems might have missed other viruses in 2020 because on the focus on COVID-19 is an alternative plausible explanation that cannot be ruled out by our analysis and deserves further study.

### S2.5 Extended excess mortality analysis

To support and supplement the analysis of excess deaths in the main text, here, in Figs S25-S29 we display the monthly observed deaths in the 2000-2020 period with their corresponding GP estimates, for each age group and each urban municipality. Likewise, in Fig. S30 we show the corresponding historical data and estimates for the entire *Greater Santiago*. These estimates are the basis for the calculations shown in Fig. 2 of the main text. In table S1 we show the coverage probabilities (the proportion of times that deaths before 2020 fell within the predicted 95% credible intervals) for each of the municipality and age specific GP model. These tend to be close to the nominal 95% value, indicating that the method is well-calibrated. Additionally, in Fig S31 we extend the time-varying excess death calculations (Fig. 2C in the main text) to all age groups. Finally, in S32 we extend Fig. 2E of the main text: we show z-scores for individual months (and not only their total, as in the main text) and we also consider the alternative measure of relative deviation in mortality from what was expected.

### S2.6 Extended case reconstructions

In order to reconstruct SARS-CoV-2 infections over time, we implemented an approach that back calculates the most likely infection tallies given the temporal sequence of deaths, the onset-to-death distribution, and the demography-adjusted infection fatality rate (IFR). Our method –referred to as RmMAP in the subsequent paragraphs– is a regularized modification of the mortality MAP (mMAP) method introduced in [36] and further investigated in [6] for COVID-19, that circumvents a well-known and undesired noise-amplification phenomenon in its original version. Operationally, RmMAP corresponds to a penalized [39], most likely de-convolution infection estimate [42]. For our analyses, we considered the log-normal onset-to-death distribution described in [20] and two demography-stratified IFR estimates, one from Diamond Princess cruise ship [21] and from a seroprevalence study in Spain [22]. For comparison, we also present reconstructions based on the Covidestim method [23], and by the re-scaling of case counts by the under-reporting estimates obtained with the method of [24]. Details are shown in S1.1.

Fig. S33 we supplements Fig. 3A of the main text, by providing reconstructions of cases in all municipalities of *Greater Santiago*. Notice these reconstructions are also used in some models for our IFR estimates (see S1.4)

### S2.7 Delay estimates

In addition to the reported timeliness (Fig 4 in the main text), we estimated mean delays according to the method described in S1.3. Results are shown in Fig. S10, where we show series of weekly estimates for the *Greater Santiago*, Fig. S11, where we show these estimates at the municipality resolution, and Fig. S12, where we show the bootstrap of distribution of the mean delay at different two-week intervals using the procedure described in S1.3

### S2.8 Extended IFR estimation analysis

Our methodology (described in (S1.4)) is inspired by ideas from [25] and [57] in that first, we leverage a proxy for ascertainment (here, positivity rates) to map the recorded cases into true infections. Second, the use of simple linear relations for the quantities within the model (here, the relation between socioeconomic status and IFR) help us smooth out the observation noises. Third, by using suitable priors we are able to borrow information on quantities we are more certainty about, and study how changes in such quantities affect inferences (sensitivity analysis). Fourth, we profit from the heterogeneity in the demographic composition of municipalities to inform and further constrain model inferences. We refer the reader to the supplementary materials for details on our models, inference methods and sensitivity analyses.

The main limitation of our methodology is that IFRs cannot be fully resolved if there is much uncertainty on ascertainment rates, age distribution of cases and prevalence (i.e., the proportion that has been infected), and therefore, the uncertainty in our IFR estimates inherits from uncertainty in these quantities. To address this limitation we have considered an ensemble of different models with different values (strong priors) for the prevalence across municipalities, each leading to different IFR estimates (see S1.4). Our displayed results are a Bayesian average of such models [58], and therefore are supposed to be more robust [52] as they take into account the uncertainty in model choice, although at the price of wider credible intervals. By using a temperature parameter *τ* we show in Figs. S35 and S36 that although there is certainly variation from model to model, the main message (the relation between IFR and SES) persist regardless of the model. Indeed,we have performed our inferences under different structural relations in our model, and results have proven stable under such changes.

As a sanity check, we also compared our estimates of IFR with those available from other countries, and find them consistent [21, 22]. Nonetheless, our sensibility analysis is not immune to possible misspecification of the age distribution of cases across municipalities, and we deem this as a potential source of bias. This may be specially critical in the youngest age group; there, only a few deaths have been observed, and therefore, our IFR estimates may be particularly unstable to changes in our assumptions. Instead of conducting additional analysis, we recognize the need for further confirmation of our results in a future study where ages of each confirmed case are available.

### S2.9 Estimation of proportion infected

In S39 we show our estimates of proportion infected on each municipality until late August, 2020. We compare Covidestim with the two methods based on a fixed IFR (from Spain and Diamond Princess, used in RmMAP) with our method that jointly estimates proportion infected and IFR (S1.4). There is a clear and consistent (across methods) negative correlation between proportion infected and SES.

### S2.10 Association between case counts, timeliness and positivity

In Fig. S41 we extend the analysis of Figs. 4C and 4G in the main text. Specifically, in the main text we show a strong association between weekly case counts and weekly positivities and timelinesses, indicating that timeliness and positivity are relevant real-time proxies of the burden of the epidemic. In S41 we average all quantities over time (weeks), and show that correlations persist, although they are attenuated, as averaging destroys the time-dependent correlational structure between cases and our measures.

This new correlation analysis is comparable with the ones for mortality (Figs 4D and 4H in the main text), and the final message is that the pattern of correlations between positivity, timeliness and the outcome is consistent regardless of whether we use cases or death as our outcome measure.

### S2.11 Case fatality ratio estimation

For estimating case-fatality ratios, we divided total cases for each municipality into age-strata in the following way: first, we looked at the age-distribution of cases, which was only available at the country level. From this distribution we compute the marginal proportion of cases among all individuals in the population, *p*_*a*_, for each age group *a*

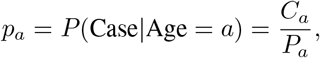

where *C*_*a*_ and *P*_*a*_ are the total of cases and individuals in the country. Then, for municipality *m* we computed the probability that a case came from group *a* using the Bayes rule,

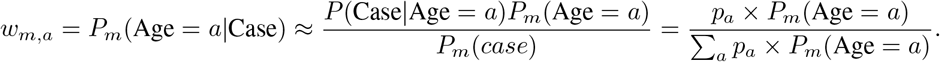

In words, we take the country-level likelihoods of cases for each age group as templates, and project them into municipality specific weights using the municipality-specific demographics represented by 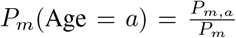 , the proportion of individuals in municipality *m* that belongs to age group *a*. To obtain age-specific municipality cases we then simply multiply *C*_*m,a*_ = *w*_*m,a*_*C*_*a*_.

We note that in our estimates we did not use any involved method for correcting for right-truncation. We justify our decision by the fact that by late August, 2020, i) the epidemic had a much smaller magnitude than at its peak, ii) enough cases and deaths were already registered so that the simple cases/death ratio was a quantity that was already stable. In Fig. S40 we show alternative estimates where we consider different recording periods *t* and offsets Δ between death and cases so that we can more generally estimate 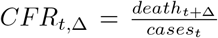. Altogether, these estimates give an idea about possible uncertainty of our estimates around their true value, and since we observe that the *CFR*_*t*,Δ_ don’t fluctuate much, this uncertainty is small and conclude that right-truncation biases don’t systematically and/or substantially affect our main conclusions.

**Figure S1:**
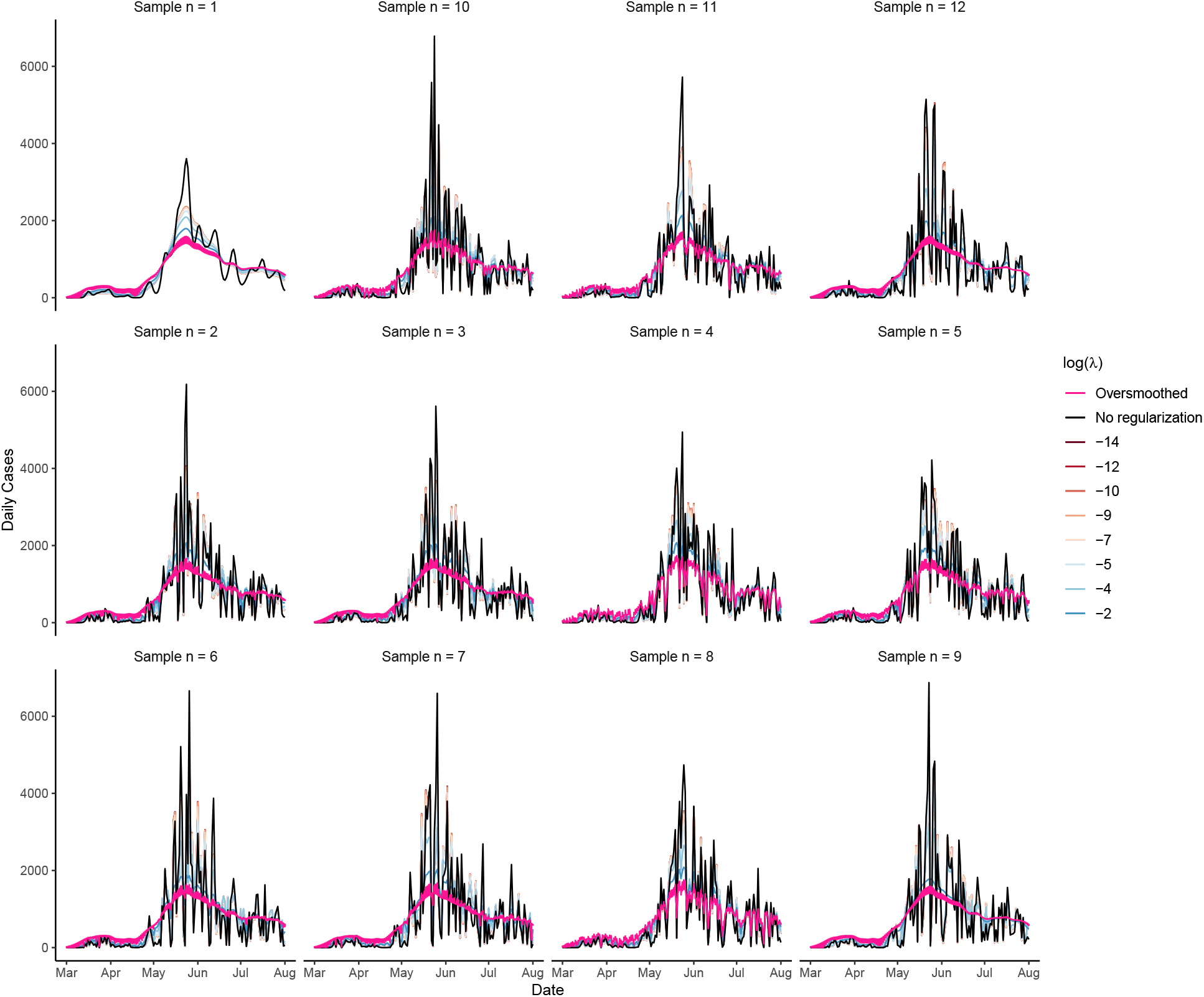
RmMAP estimates for different values of. *λ* (different colors), including *λ* = 0 (no regularization), based on mortality data from the Maipu municipality. Each of the twelve plots corresponds to a different initial random seed (first plot is the uniform initialization) Curves that do not satisfy the goodness of fit criterion are colored in pink (*oversmoothed*). Daily case counts are obtained by weighing the demography adjusted IFR (here, from Spain seroprevalence data).

**Figure S2:**
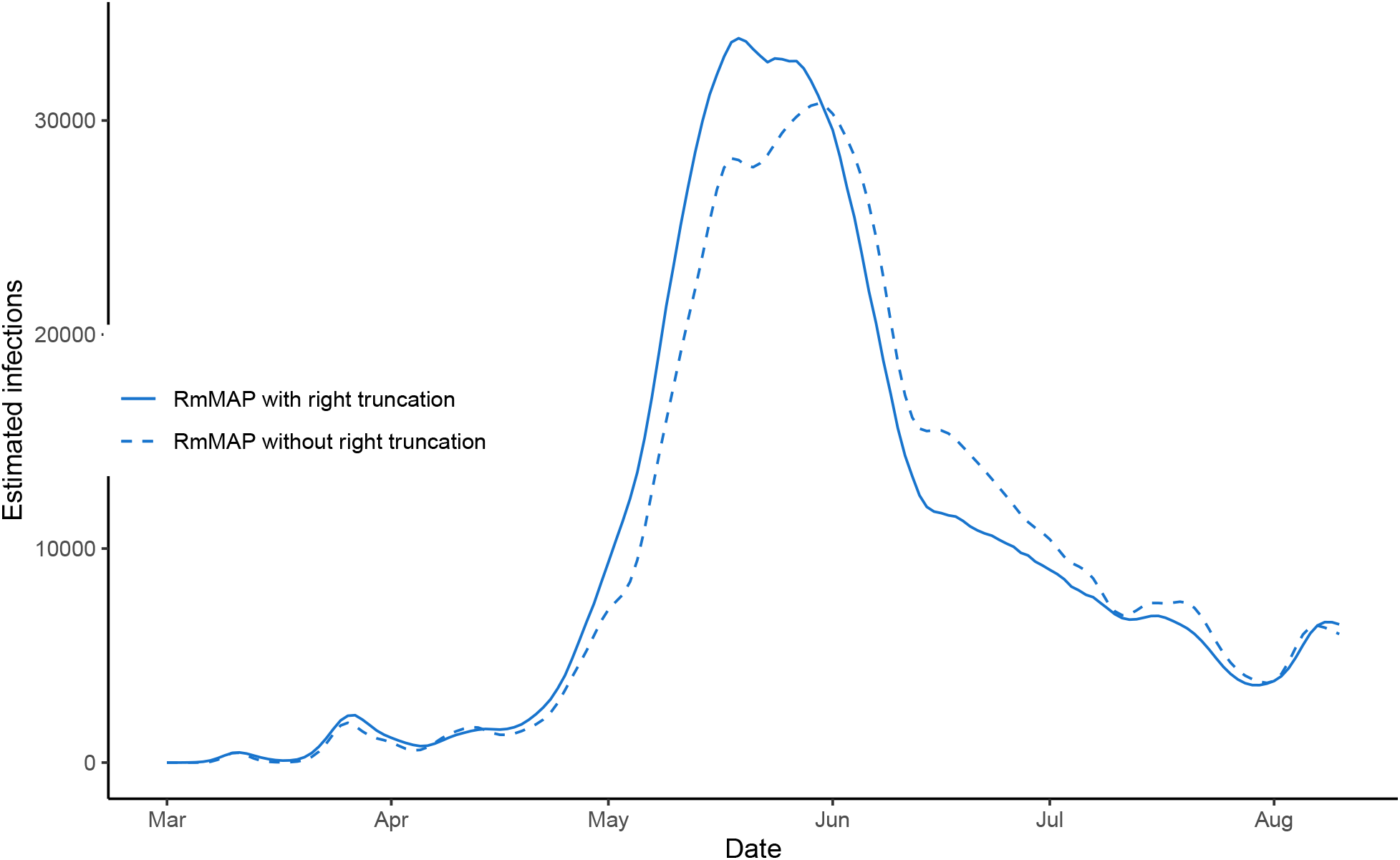
Robustness of reconstructions to changes in delay distribution. We compared reconstructions in the *Greater Santiago* using two log-normal onset-to-death delay distributions (with parameters estimated with and without right truncation), as described in [20].

**Figure S3:**
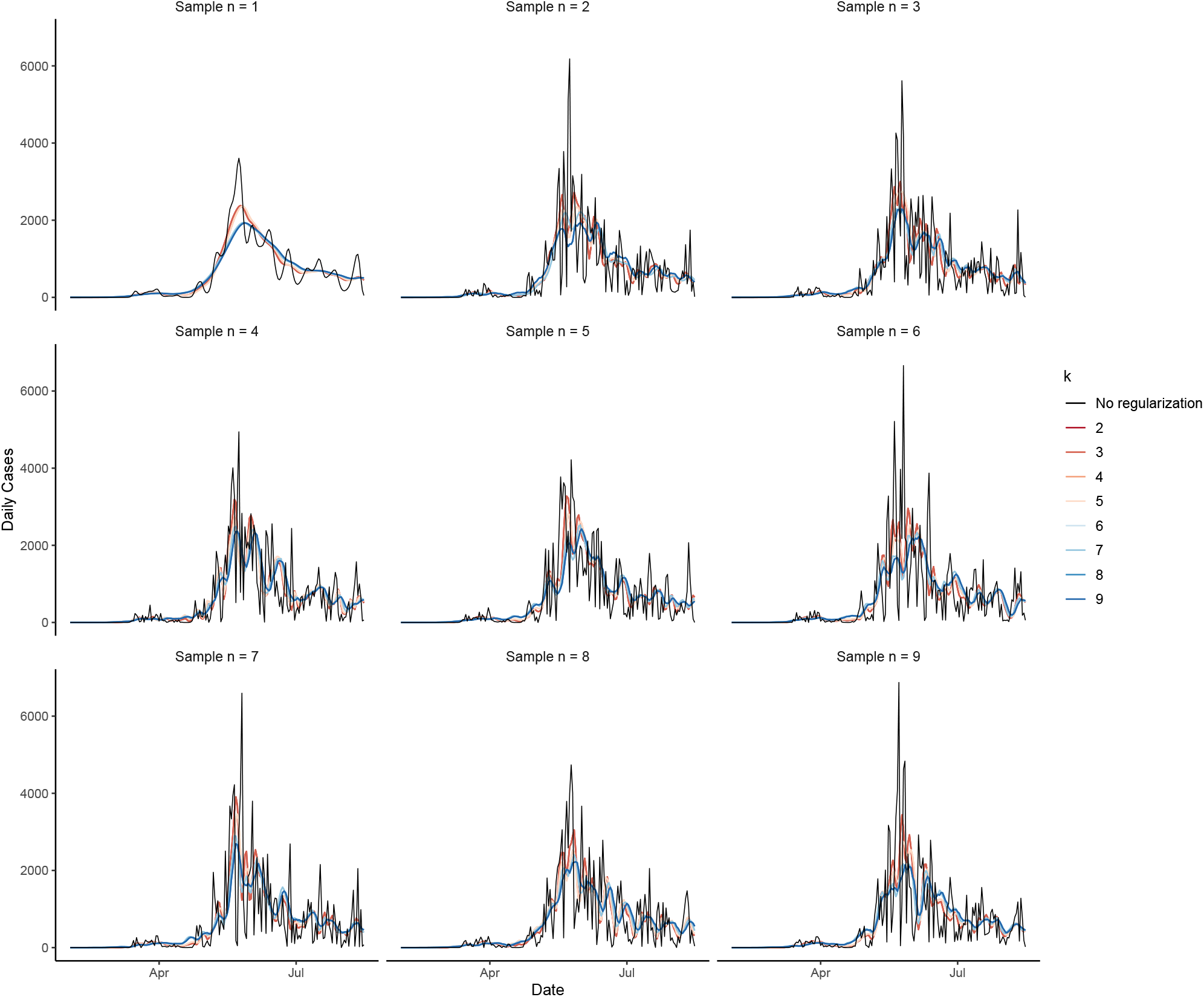
EMS estimates for different values of smoothing window. *k* The setup is analog to Fig S1.

**Figure S4:**
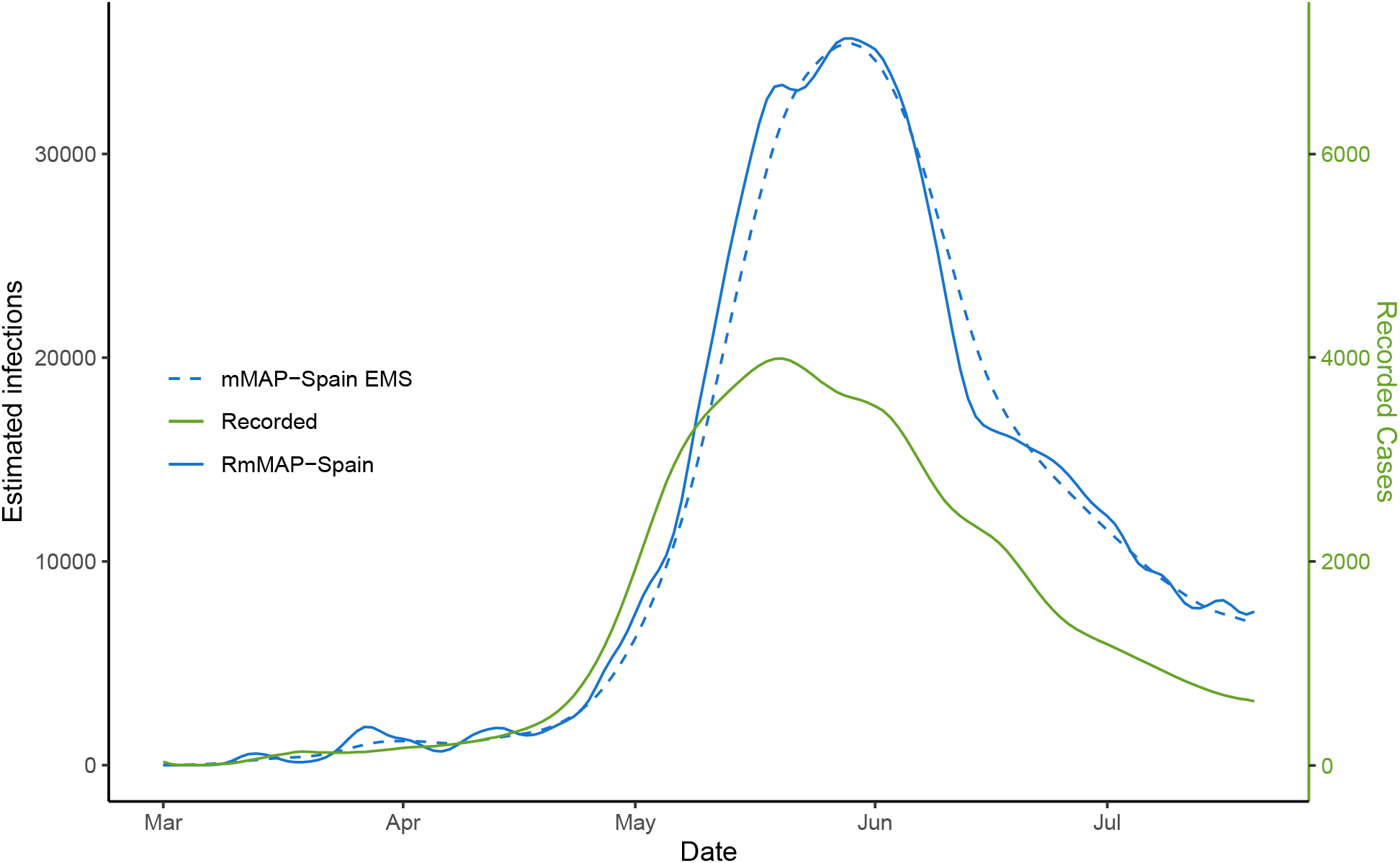
Comparison between RmMAP and EMS for the reconstruction of epidemic at the entire *Greater Santiago*. This figure supplements the one of main text (Fig. 3A) by providing EMS reconstructions as well.

**Figure S5:**
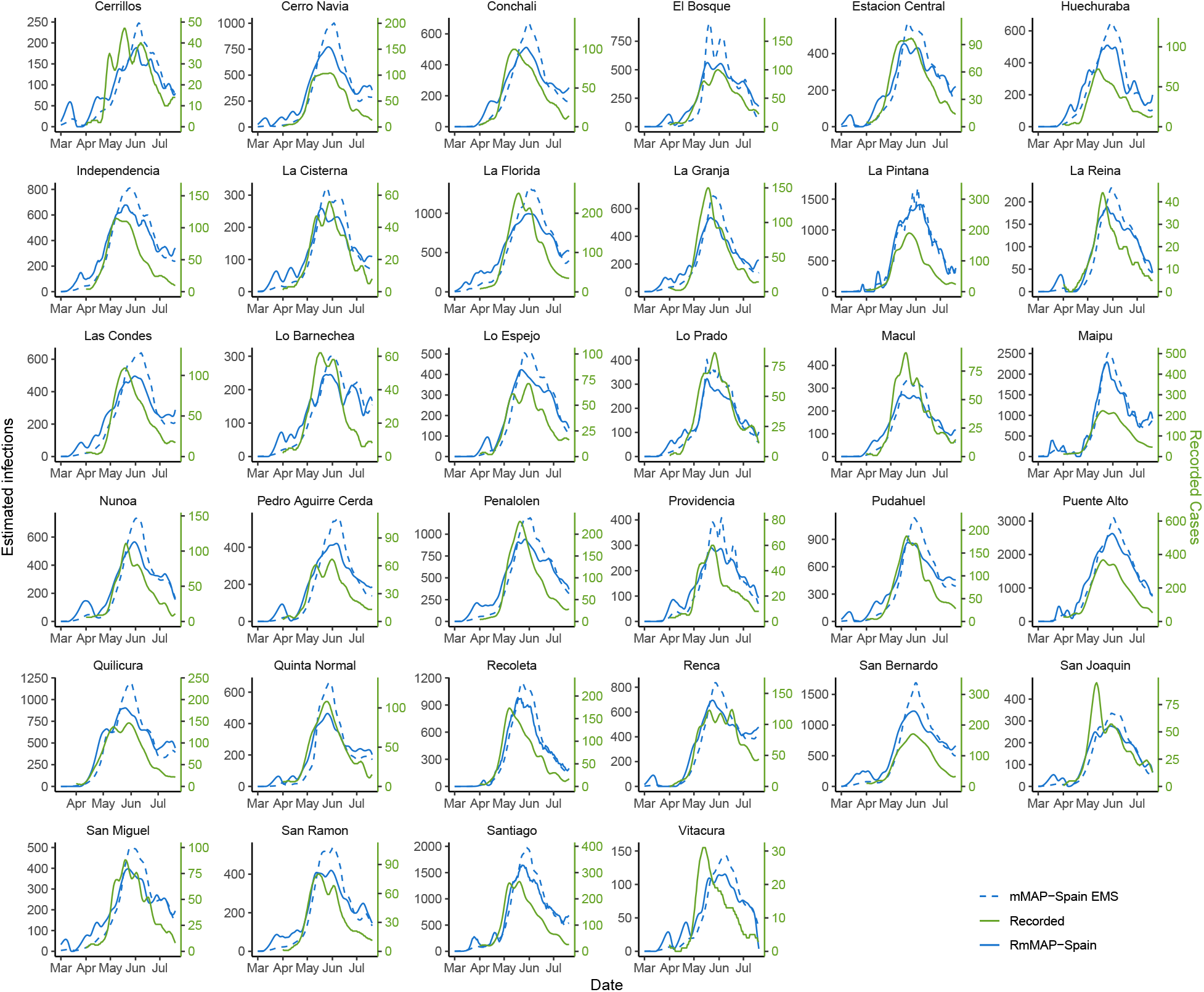
Comparison between RmMAP and EMS for the reconstruction of epidemic municipalities in the *Greater Santiago*. While RmMAP recovers an early peak in many municipalities, this is not the case with EMS, whose reconstructions tend to suggest a single peak.

**Figure S6:**
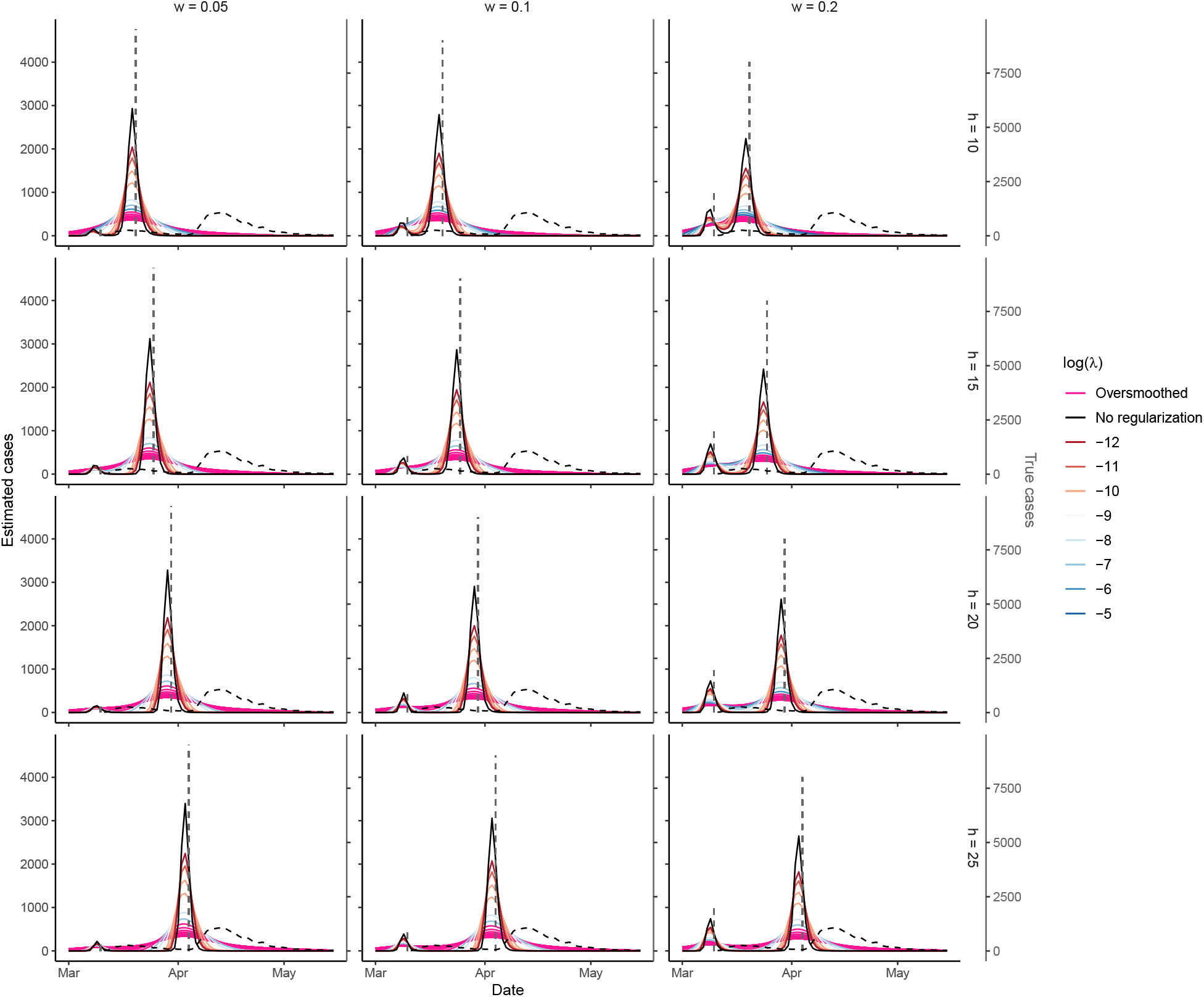
Results of a synthetic scenario for RmMAP. Each plot represents a different configuration of *h* (separation between peaks) and *w* (relative size of the earliest peak). Dashed vertical gray lines indicate magnitude of two point event (cases, on a different scale shown on the right y-axis) and dashed black line indicates the distribution of observed deaths, the convolution *D*\**S* between the onset-to-death distribution *D* and the sum *S* of the two point events, 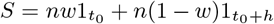. Coloring of solid lines is analog to Fig S1. We observe that in the majority of cases there is a range of valid regularization parameters *λ* leading to reconstructions that account for the two peaks, i.e. RmMAP does perform well. For simplicity we have assumed *IFR* = 1.

**Figure S7:**
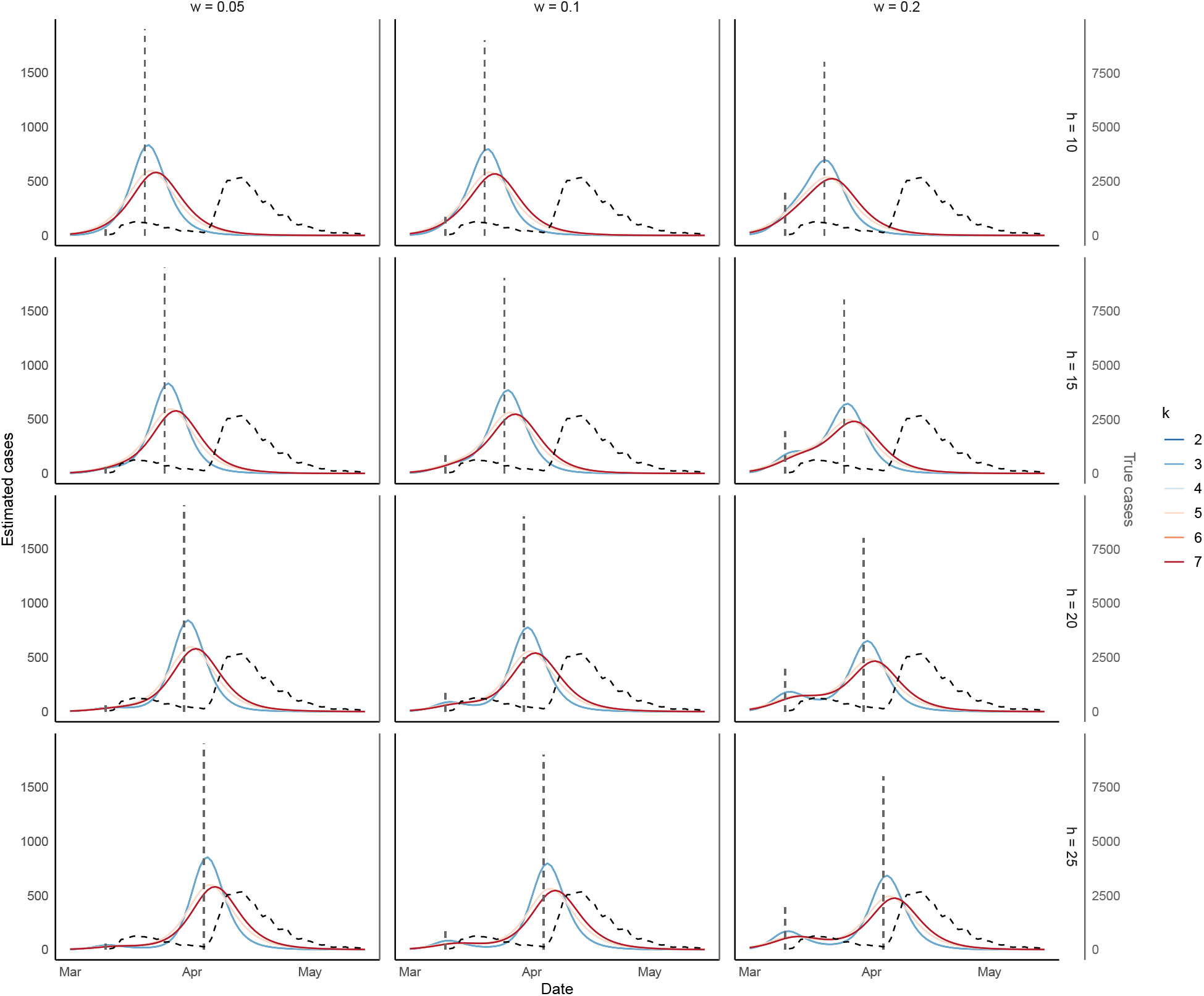
Results of a synthetic scenario for EMS. Setup is analog to Fig S7. We observe most of the cases EMS fails to recover the bi-modality of our hypothetical scenario, for all possible values of smoothing window *k*.

**Figure S8:**
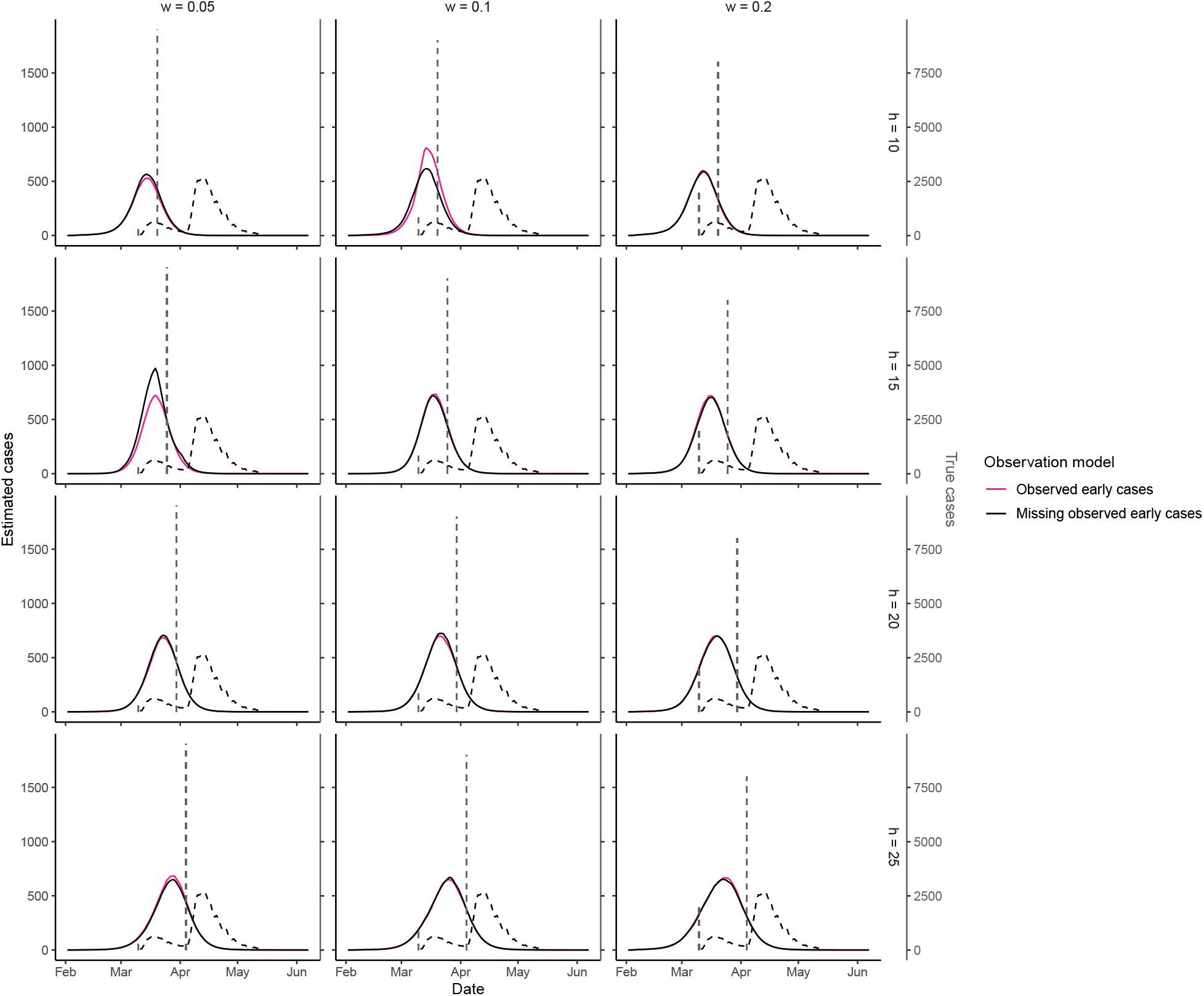
Results of a synthetic scenario for *covidestim*. Setup is analog to Fig S7. We assume that either all cases are observed (pink) or only the second peak is recorded (black). In both cases, Covidestim fails to recover a bimodal distribution of cases.

**Figure S9:**
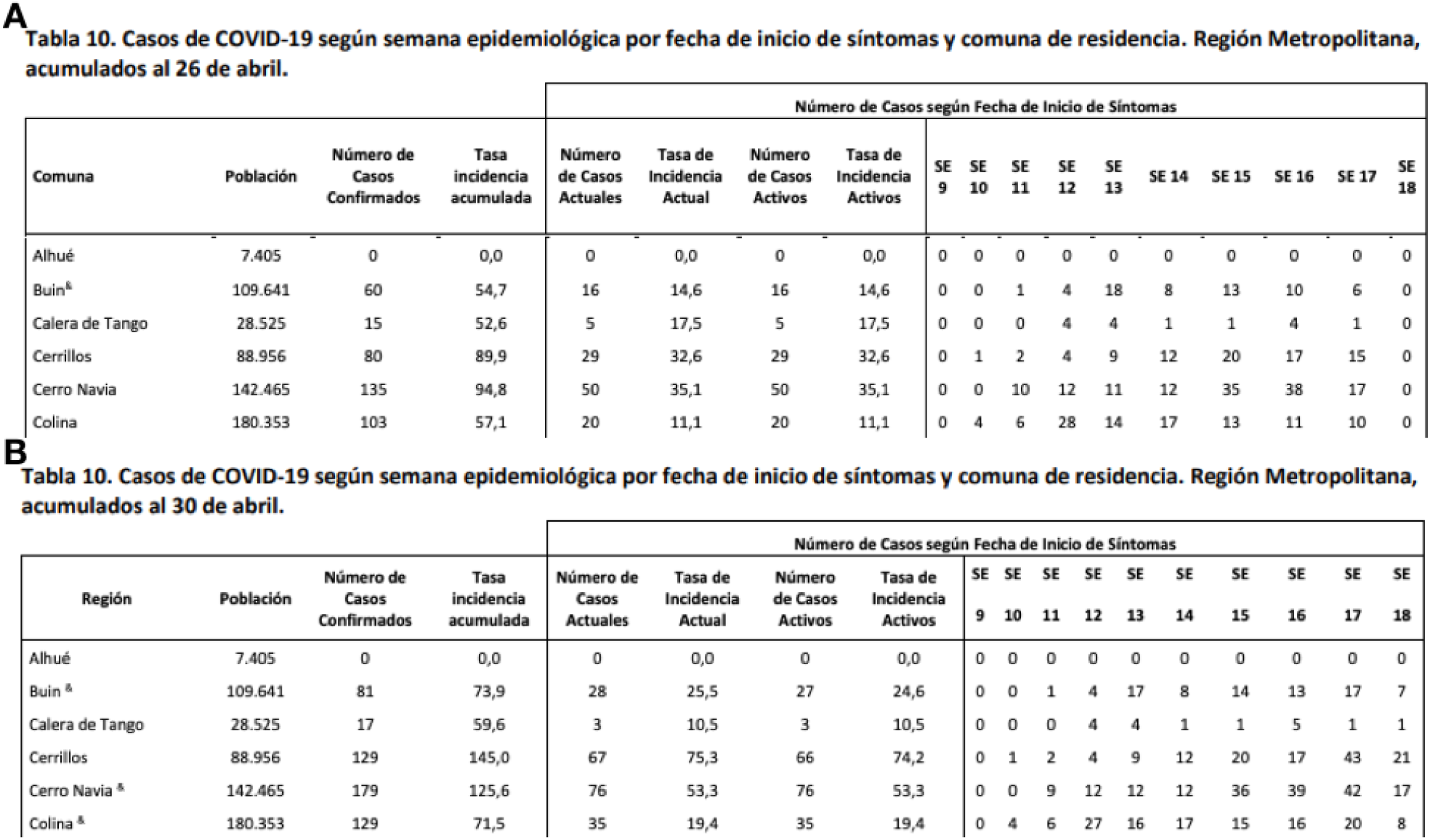
Examples of available data by onset of symptoms. **A** and **B** correspond to reports at two consecutive reports (April 26th and April 30th, 2020) in some municipalities at Region Metropolitana. The last columns show cases that has been reported with onset of symptoms at each epidemiological week (SE).

**Figure S10:**
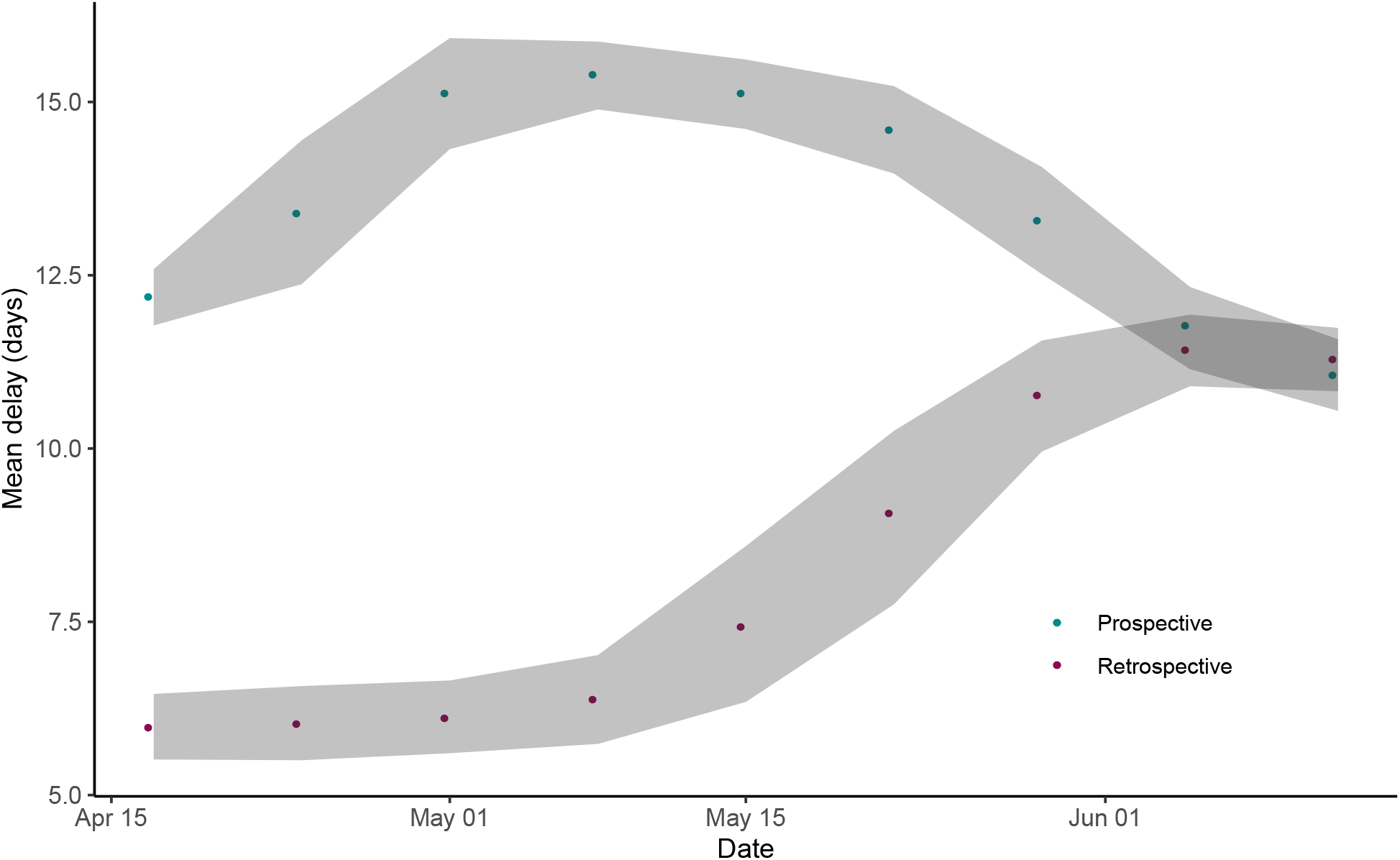
Mean delay (days) between onset of symptoms to public report in Region Metropolitana. Shading indicates 95% confidence interval. We consider retrospective (aligned to date of report) and prospective (aligned to onset of symptoms). Since reporting occurs only twice a week, prospective report is biased to be larger than the retrospecive one. Our analysis encompasses only until early June, 2020, since in mid June public data appeared heavily corrupted, making estimation more virtually impossible.

**Figure S11:**
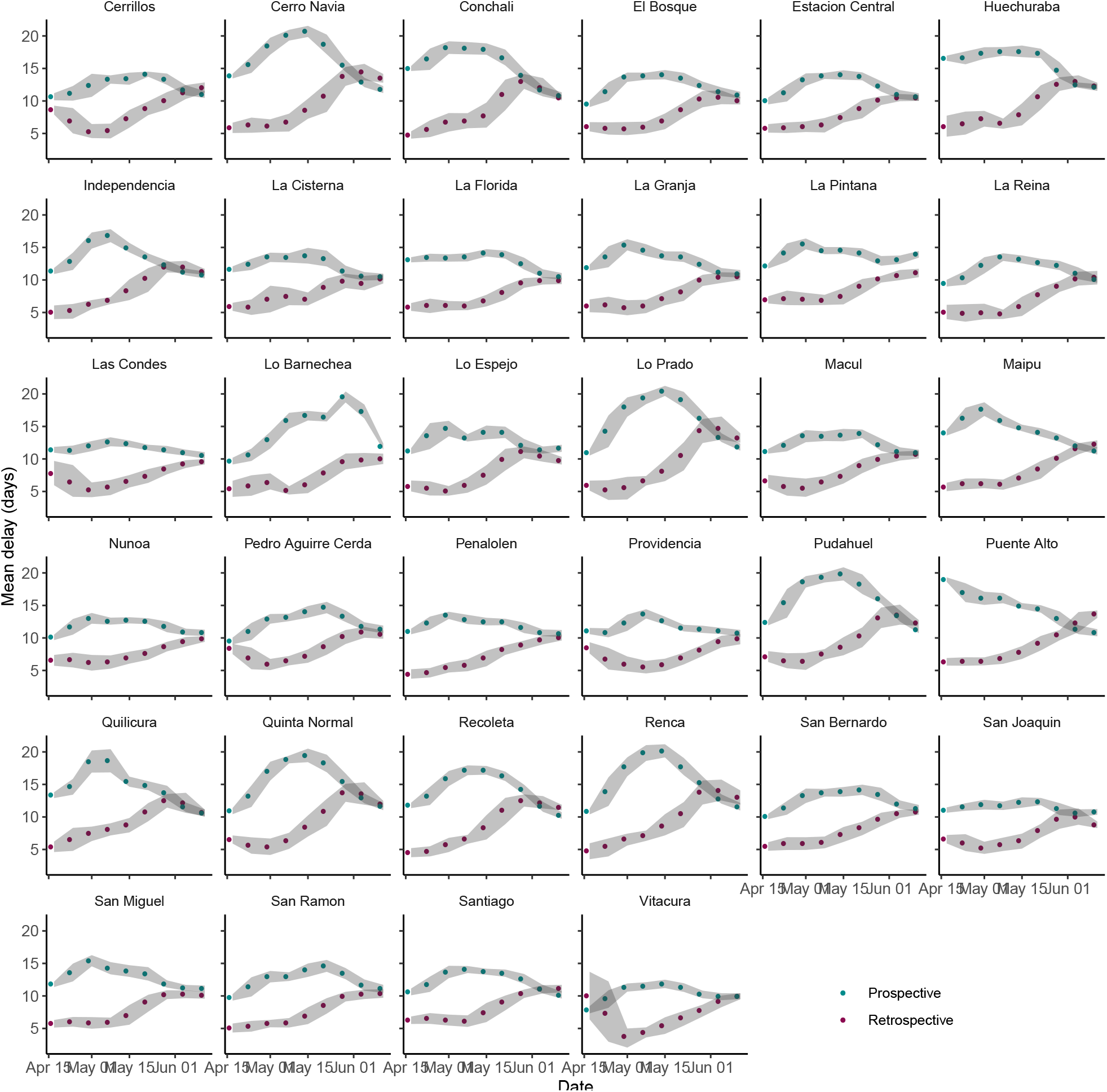
Mean delay (days) between onset of symptoms to public report in urban municipalities. Setup is the same as in Fig S10.

**Figure S12:**
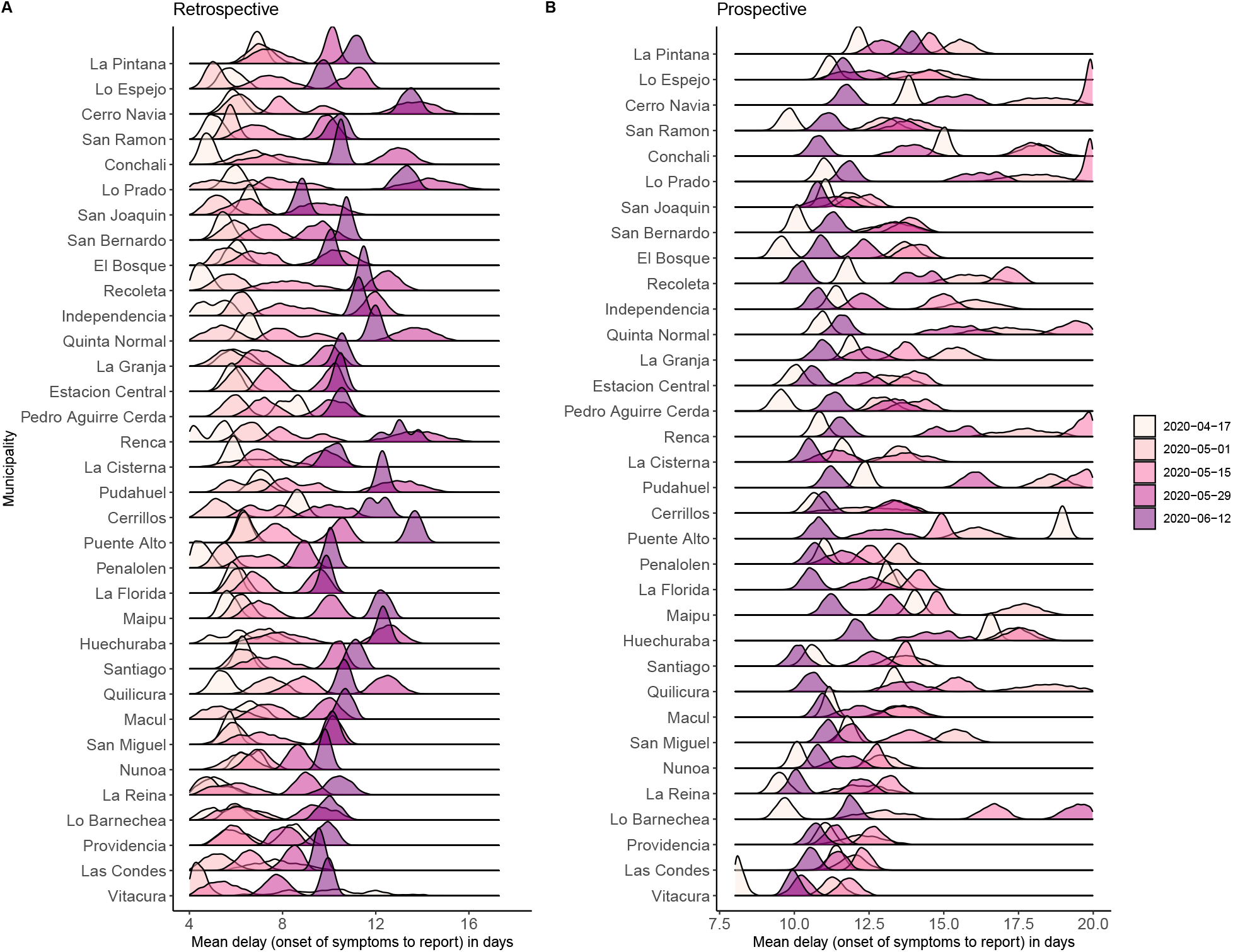
Distribution of delay at urban municipalities during selected times. Delays are highly variable and heterogeneous among municipalities. The information displayed closely relates to the one shown in Fig S11.

**Figure S13:**
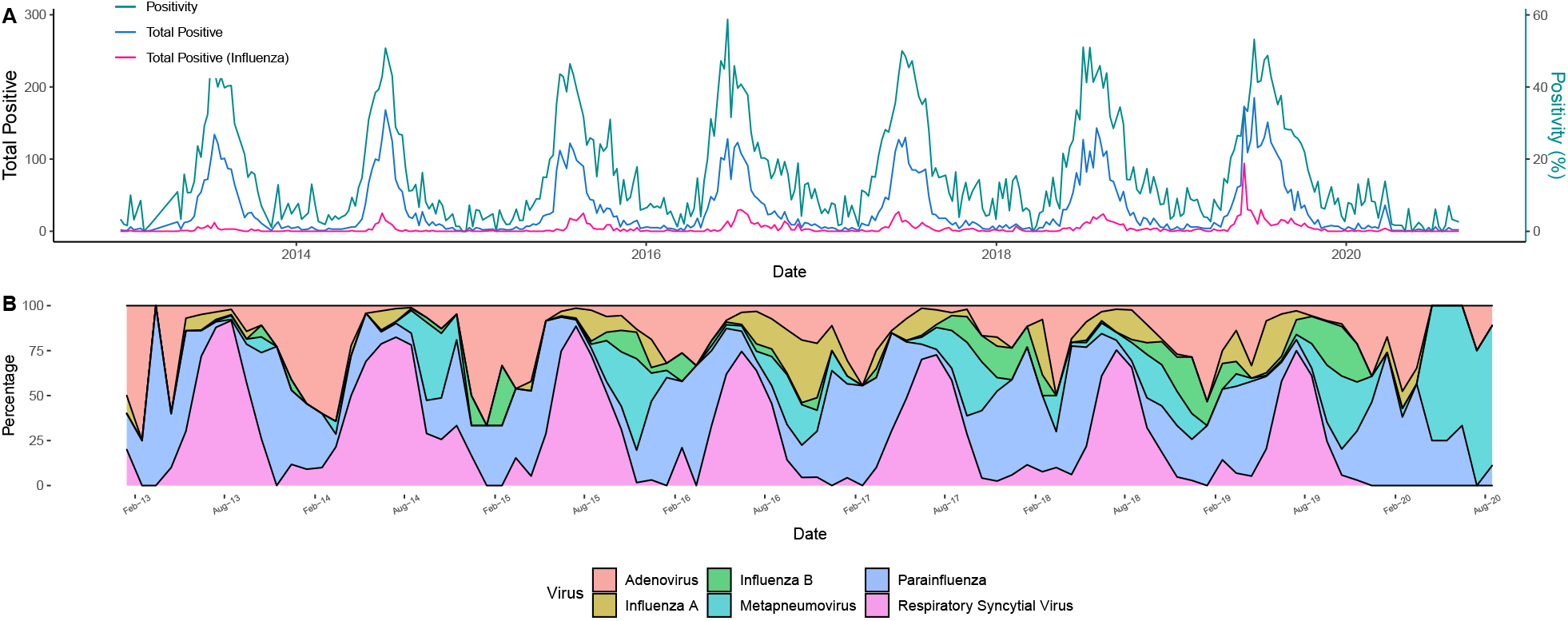
Historical reports of respiratory viral circulation over a hospitalized population suggest the 2020 season is unusual. **A**:Previous years have experienced seasonal (winter) peaks of viral activity, and influenza (A and B sub-types) is a major contributor. During winter seasons positivity also tends to rise. For the 2020 winter season the amount of tests, diagnosis and positivity have been unusually low. **B**: Data is available for 7 respiratory viruses, whose activity and share of positives fluctuates across years and seasons.

**Figure S14:**
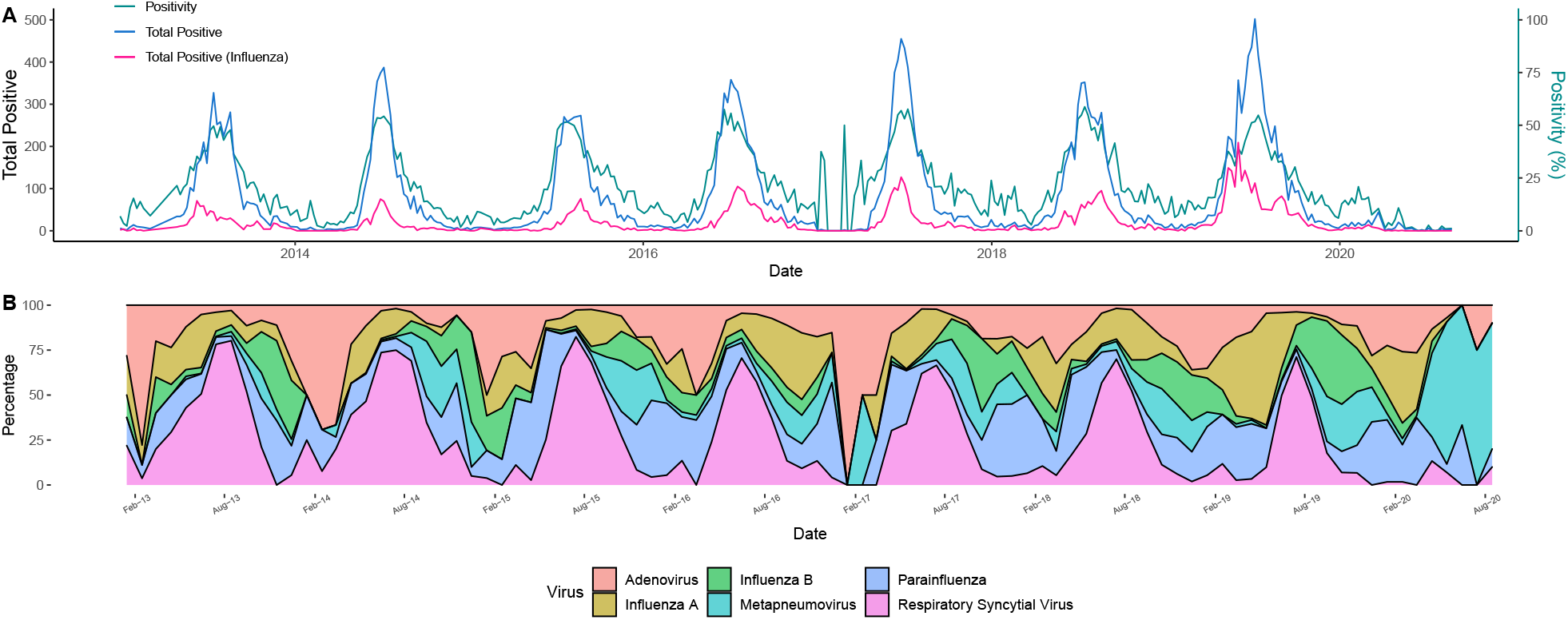
Historical reports of respiratory viral circulation suggest the 2020 season is unusual. Details are the same as in Fig S13, but over the general population, not necessarily hospitalized.

**Figure S15:**
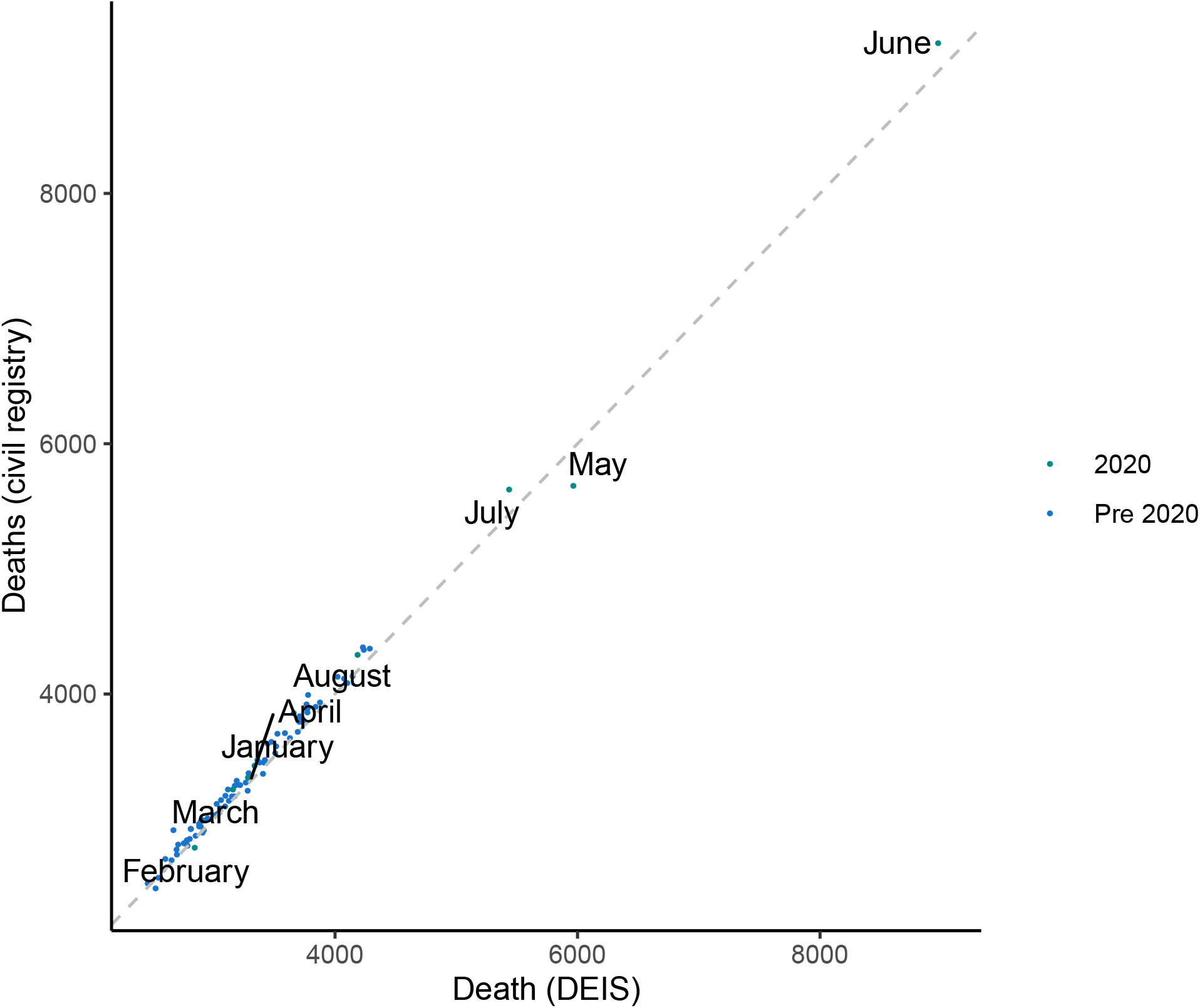
Number of registered deaths per month in Region Metropolitana from January 2010 to July 2020, according to DEIS and the civil registry.

**Figure S16:**
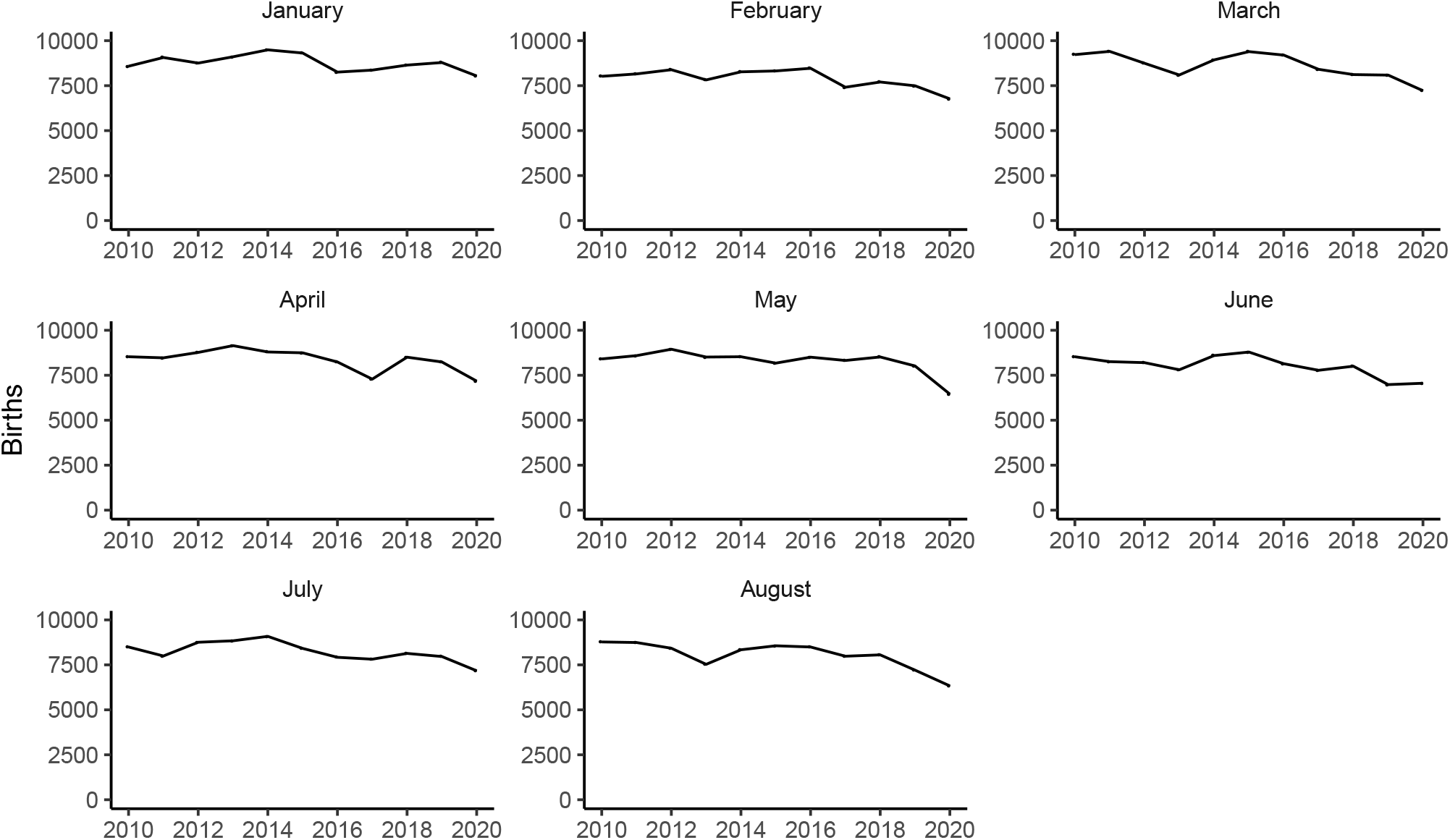
**Number of registered births per month in Region Metropolitana**, from January 2010 to July 2020.

**Figure S17:**
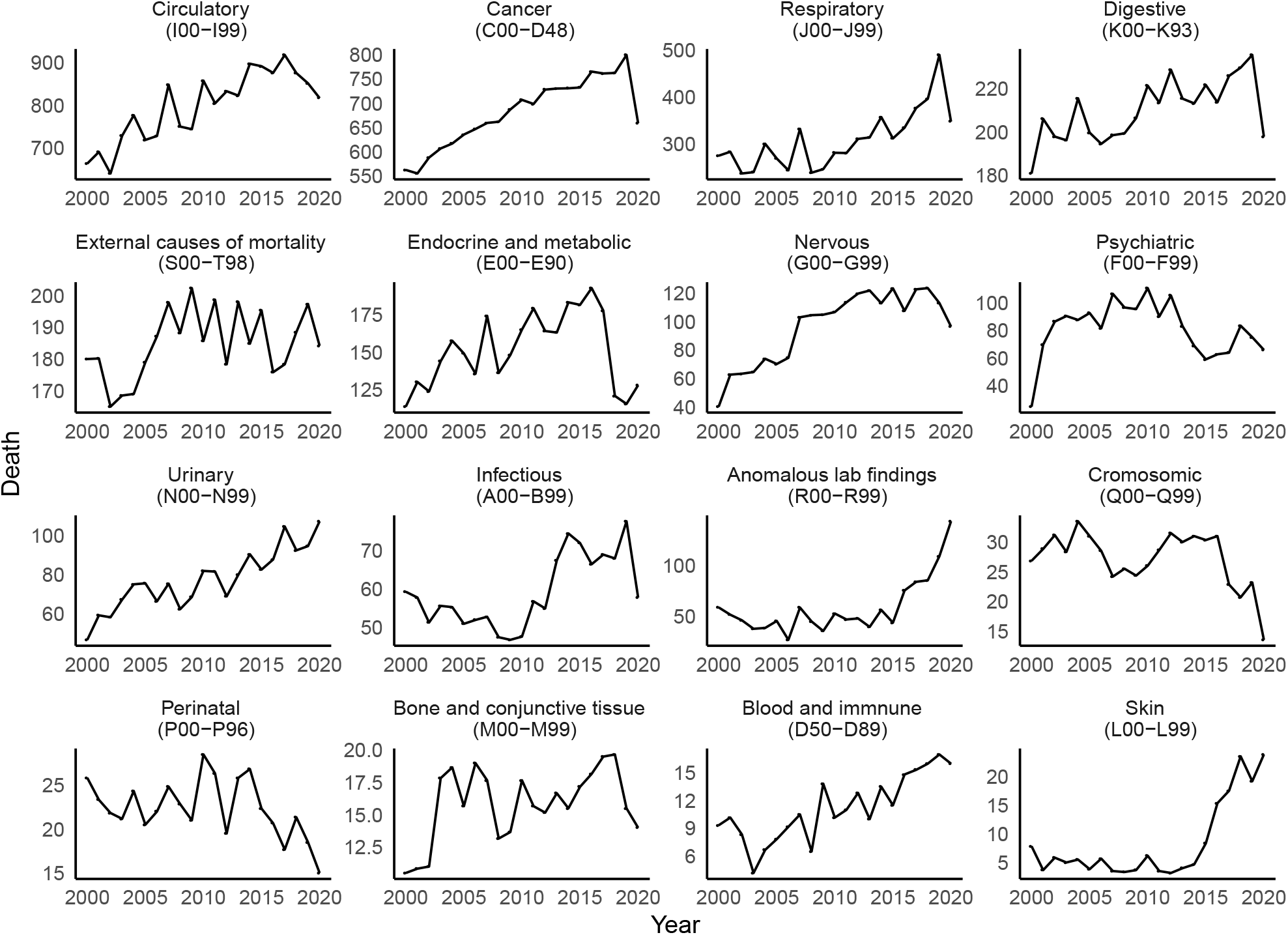
**Total deaths by each cause in *Greater Santiago***, with their ICD-10 codes, between March and August of each year.

**Figure S18:**
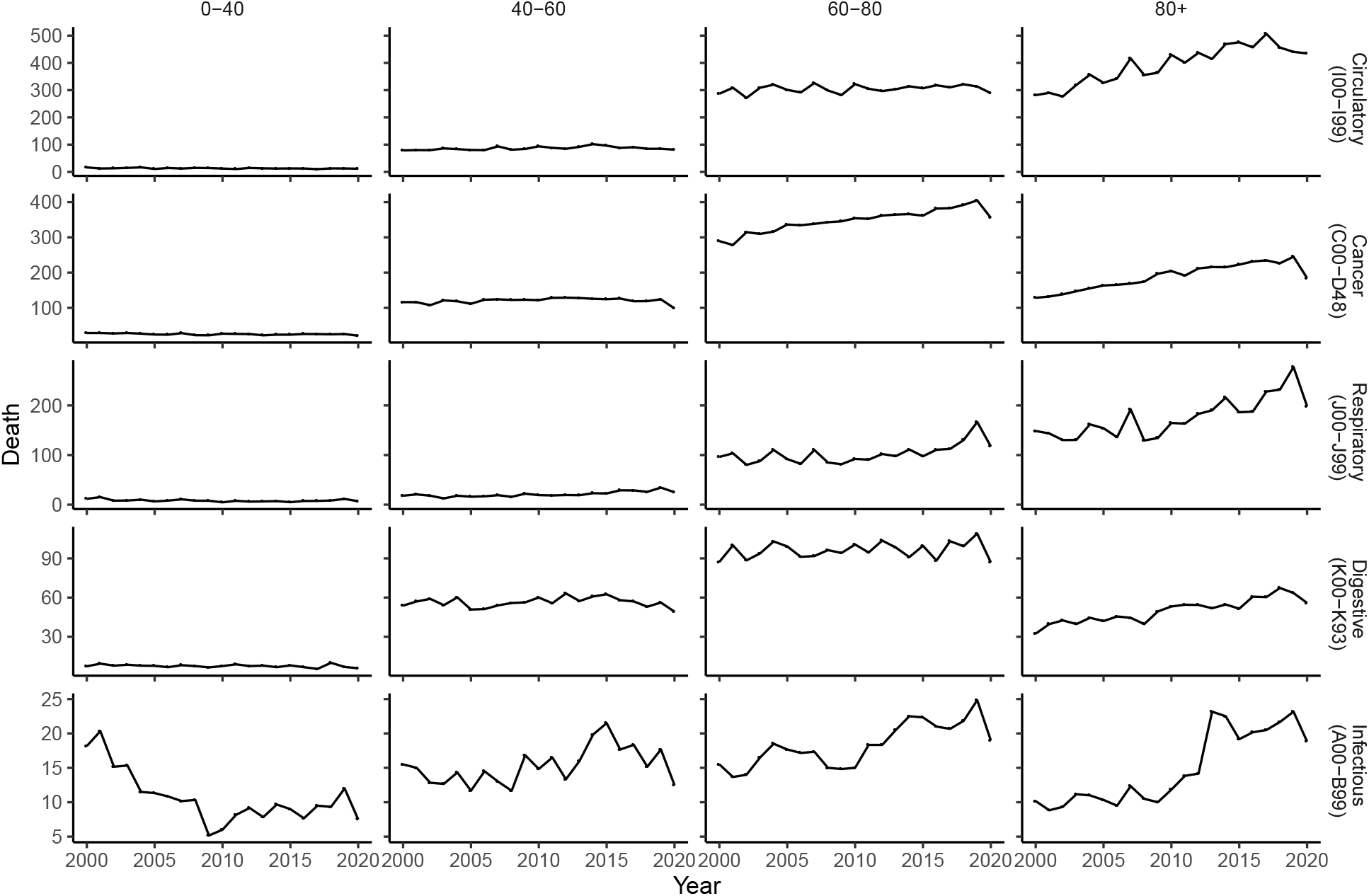
**Total deaths by major causes and age groups in *Greater Santiago***,between March and August of each year.

**Figure S19:**
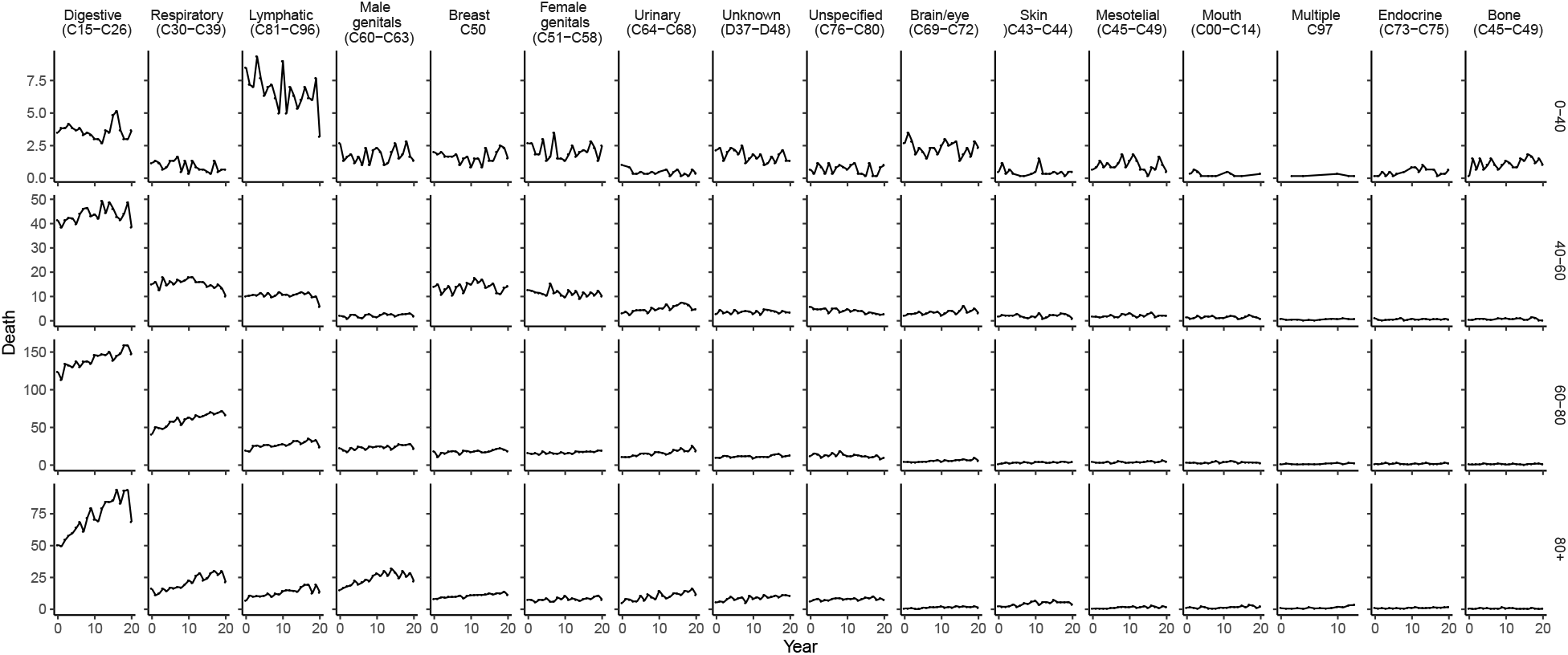
**Total deaths by cancer, by cancer type and age group in *Greater Santiago***,between March and August of each year.

**Figure S20:**
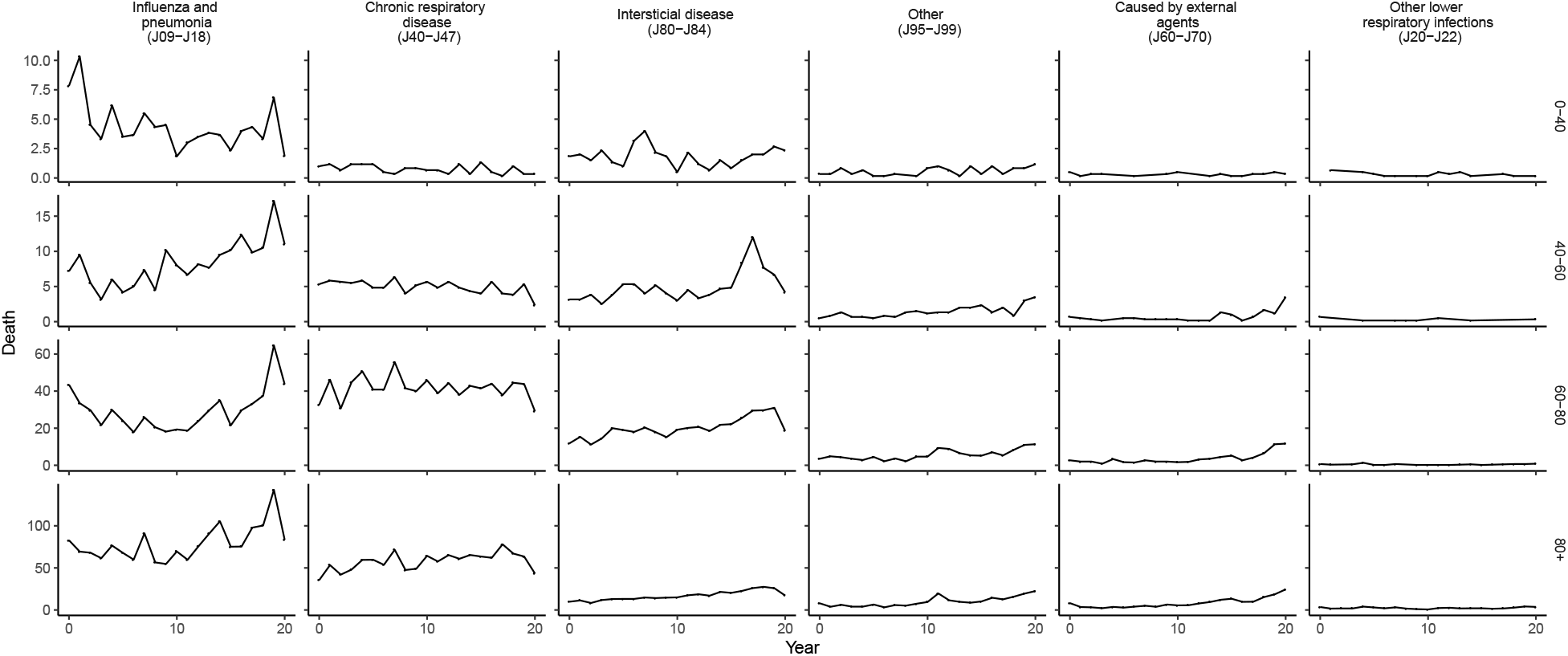
**Total deaths by respiratory causes, by sub-cause and age group in *Greater Santiago***,between March and August of each year.

**Figure S21:**
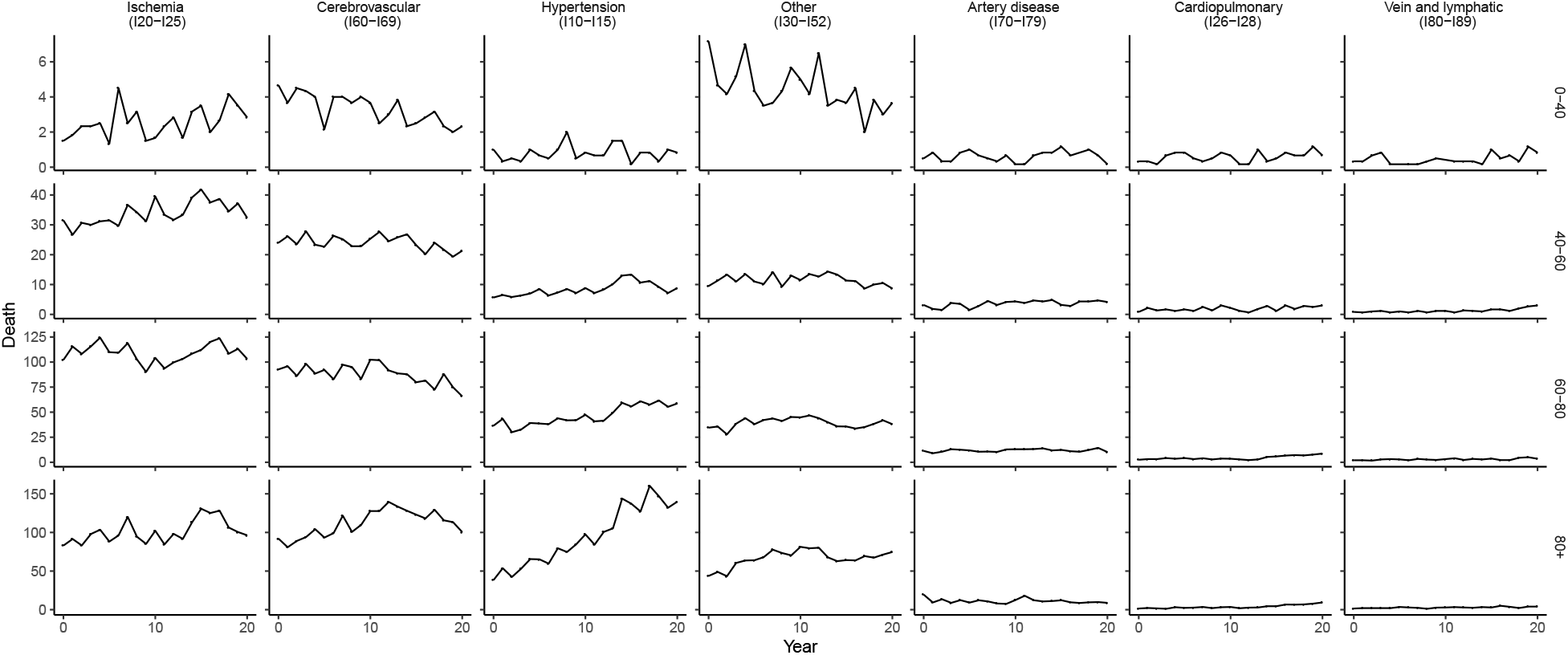
**Total deaths by circulatory cause, by sub-cause and age group in *Greater Santiago***, between March and August of each year.

**Figure S22:**
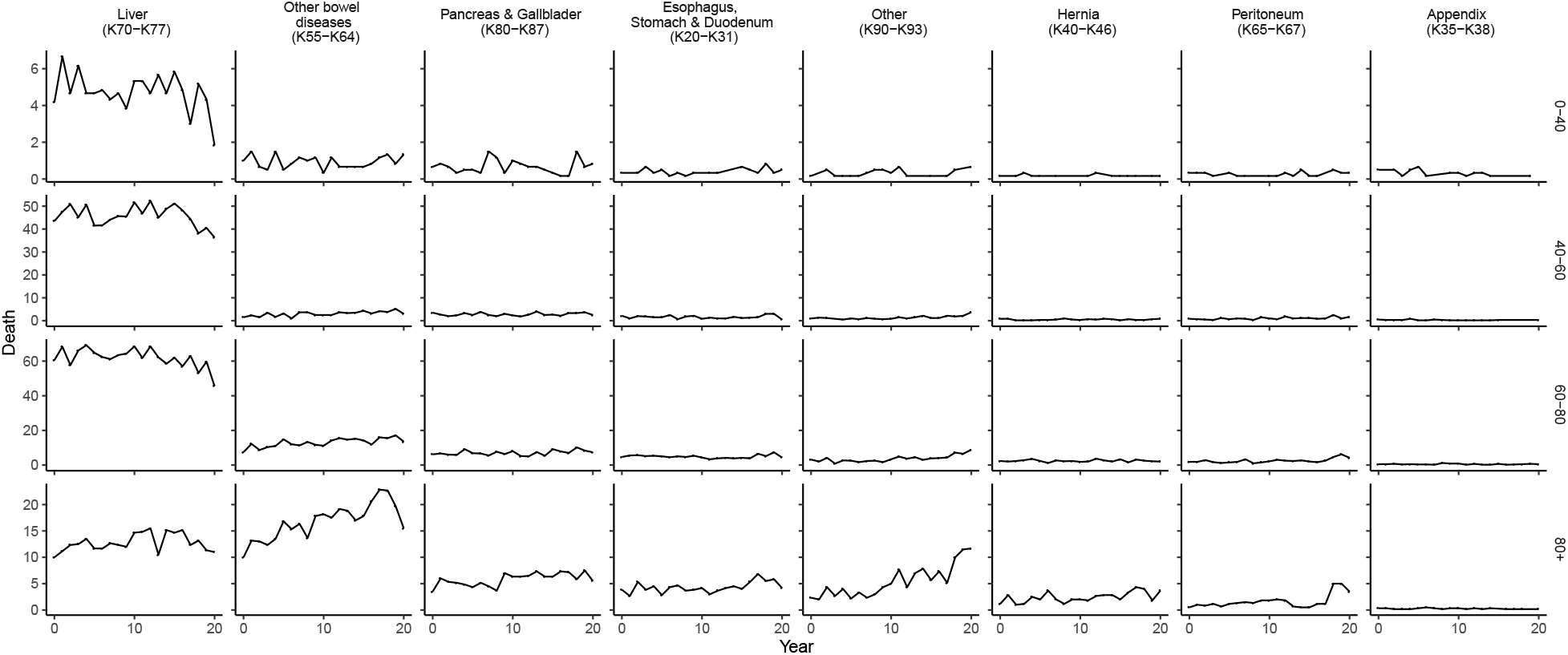
**Total deaths by digestive cause, by sub-cause and age group in *Greater Santiago***, between March and August of each year.

**Figure S23:**
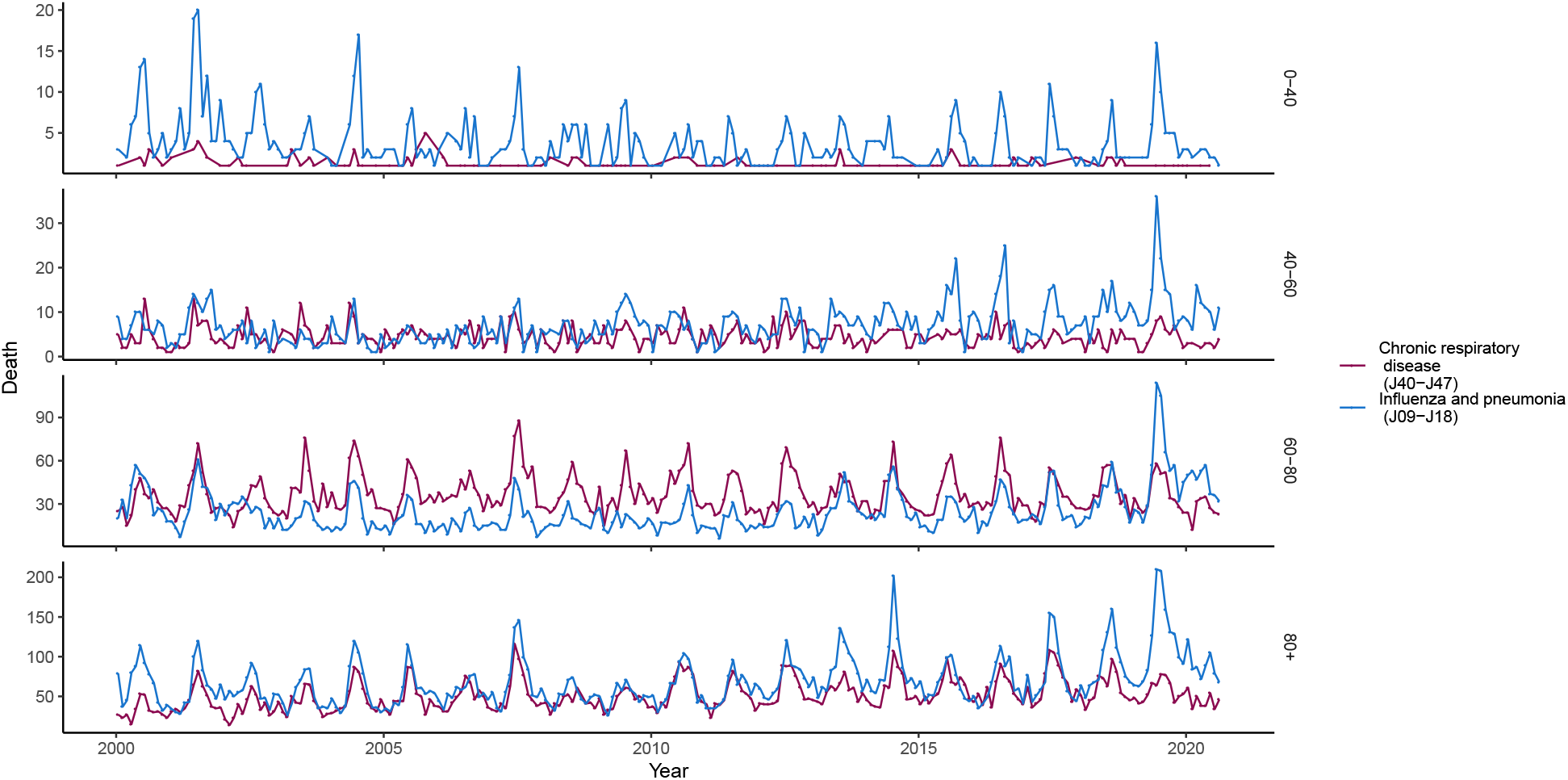
Historical weekly deaths in *Greater Santiago* by some respiratory causes reveal a seasonal trend for some age groups.

**Figure S24:**
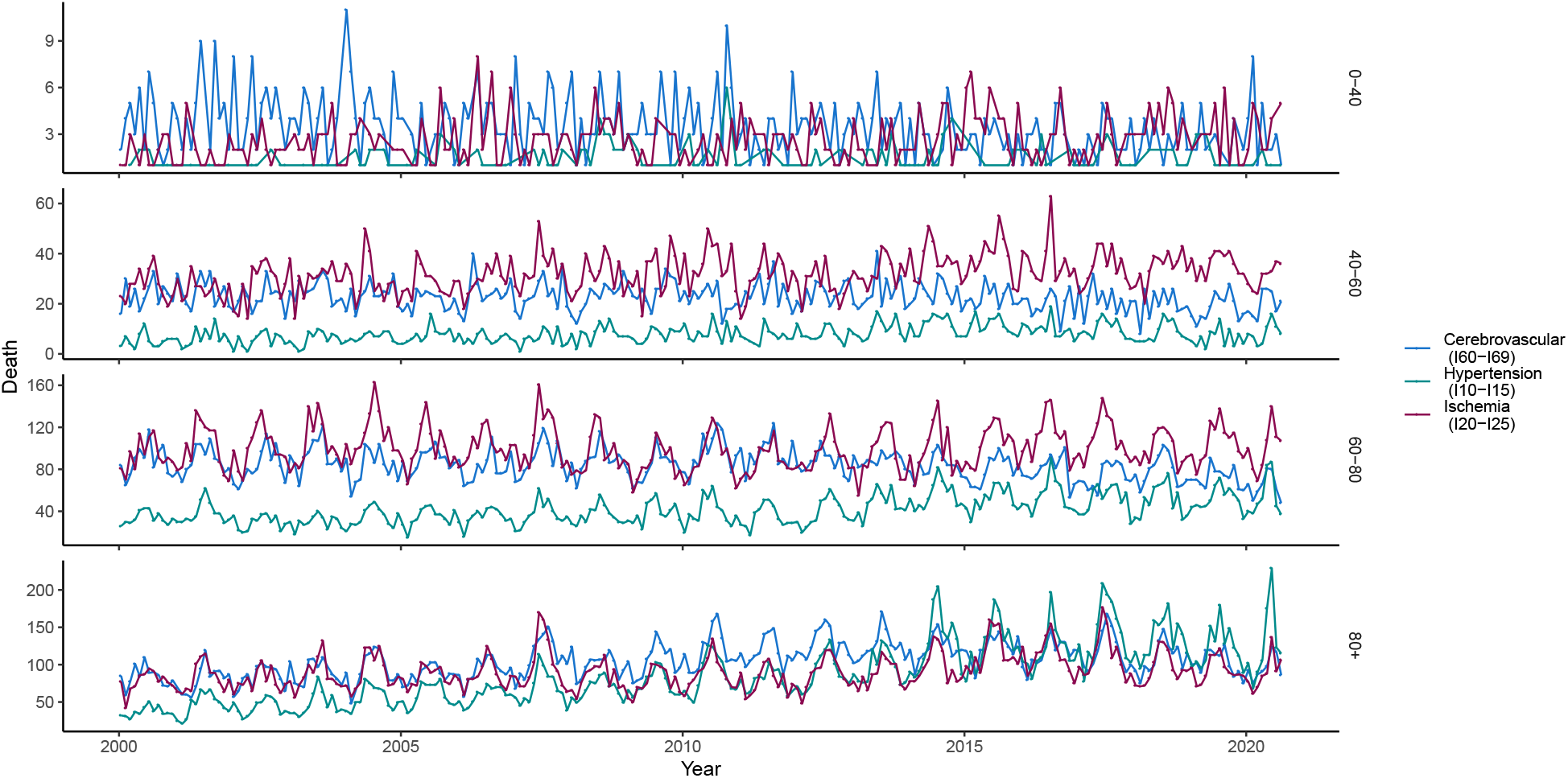
Historical weekly deaths in *Greater Santiago* by some circulatory causes reveal a seasonal trend for some age groups.

**Figure S25:**
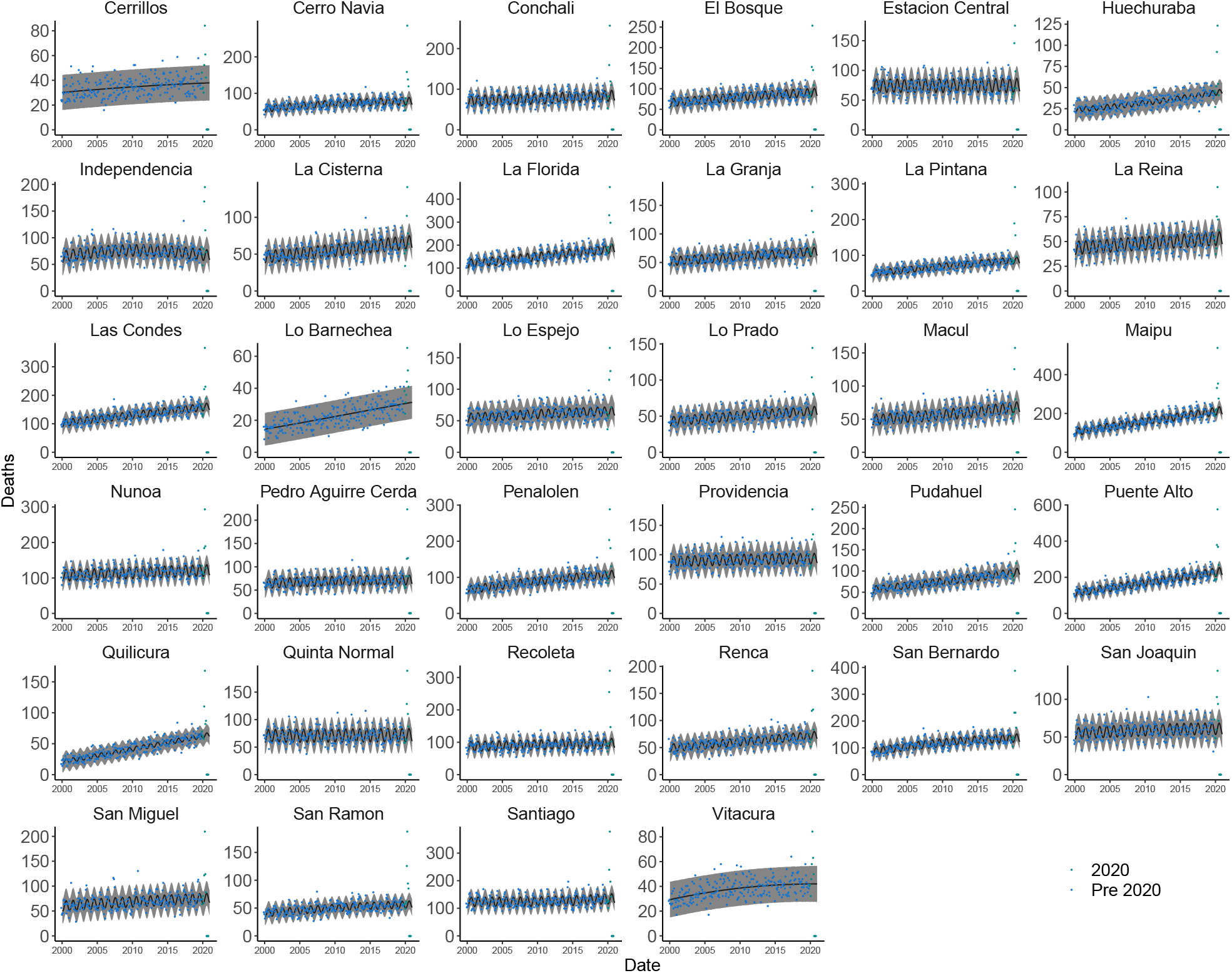
Number of monthly deaths in 2002-2020 in urban municipalities, all ages. GP predictions and confidence intervals are shown as black line surrounded by gray areas, respectively

**Figure S26:**
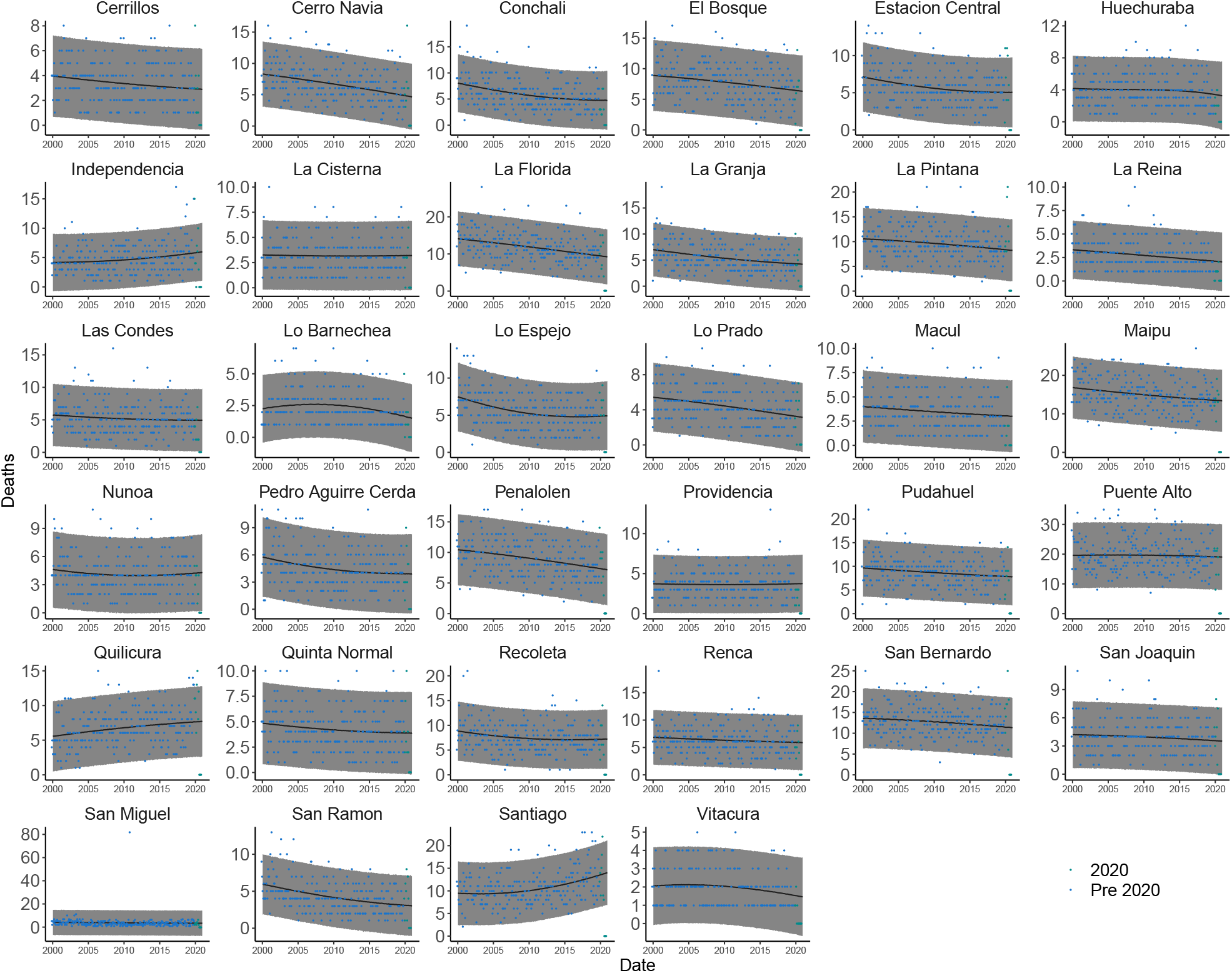
Number of monthly deaths in 2002-2020 in urban municipalities, 0-40 age group. GP predictions and confidence intervals are shown as black line surrounded by gray areas, respectively

**Figure S27:**
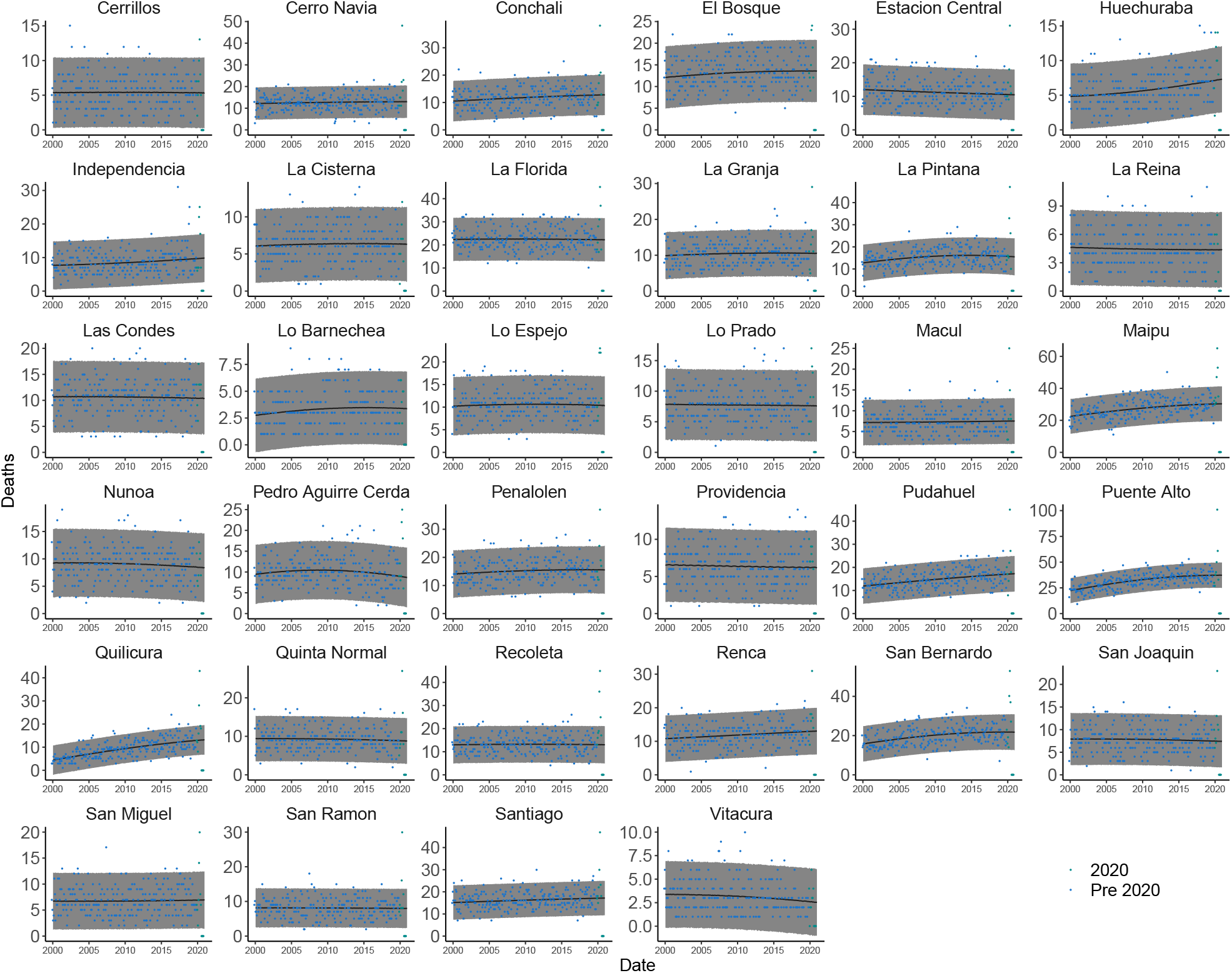
Number of monthly deaths in 2002-2020 in urban municipalities, 40-60 age group. GP predictions and confidence intervals are shown as black line surrounded by gray areas, respectively

**Figure S28:**
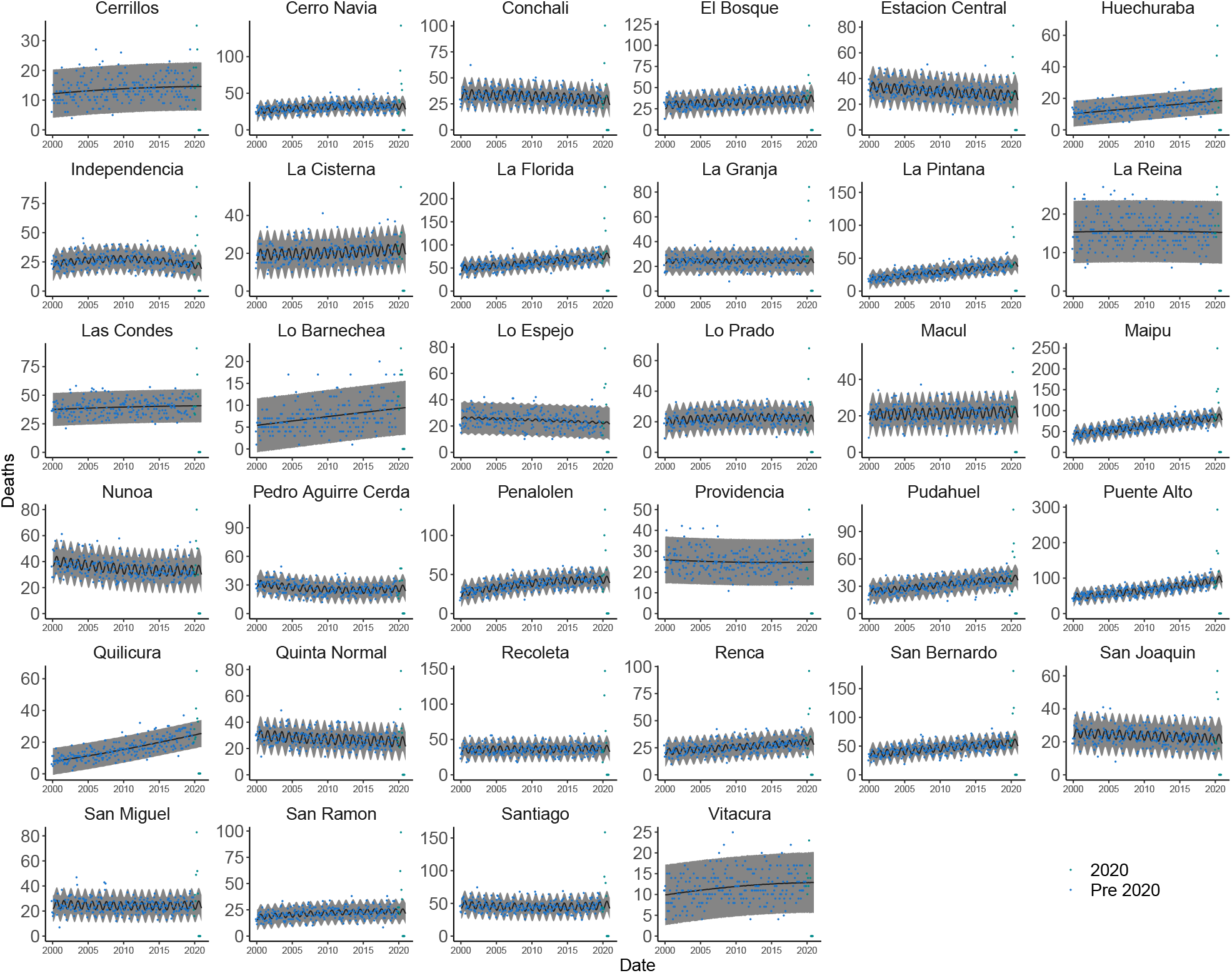
Number of monthly deaths in 2002-2020 in urban municipalities, 60-80 age group. GP predictions and confidence intervals are shown as black line surrounded by gray areas, respectively

**Figure S29:**
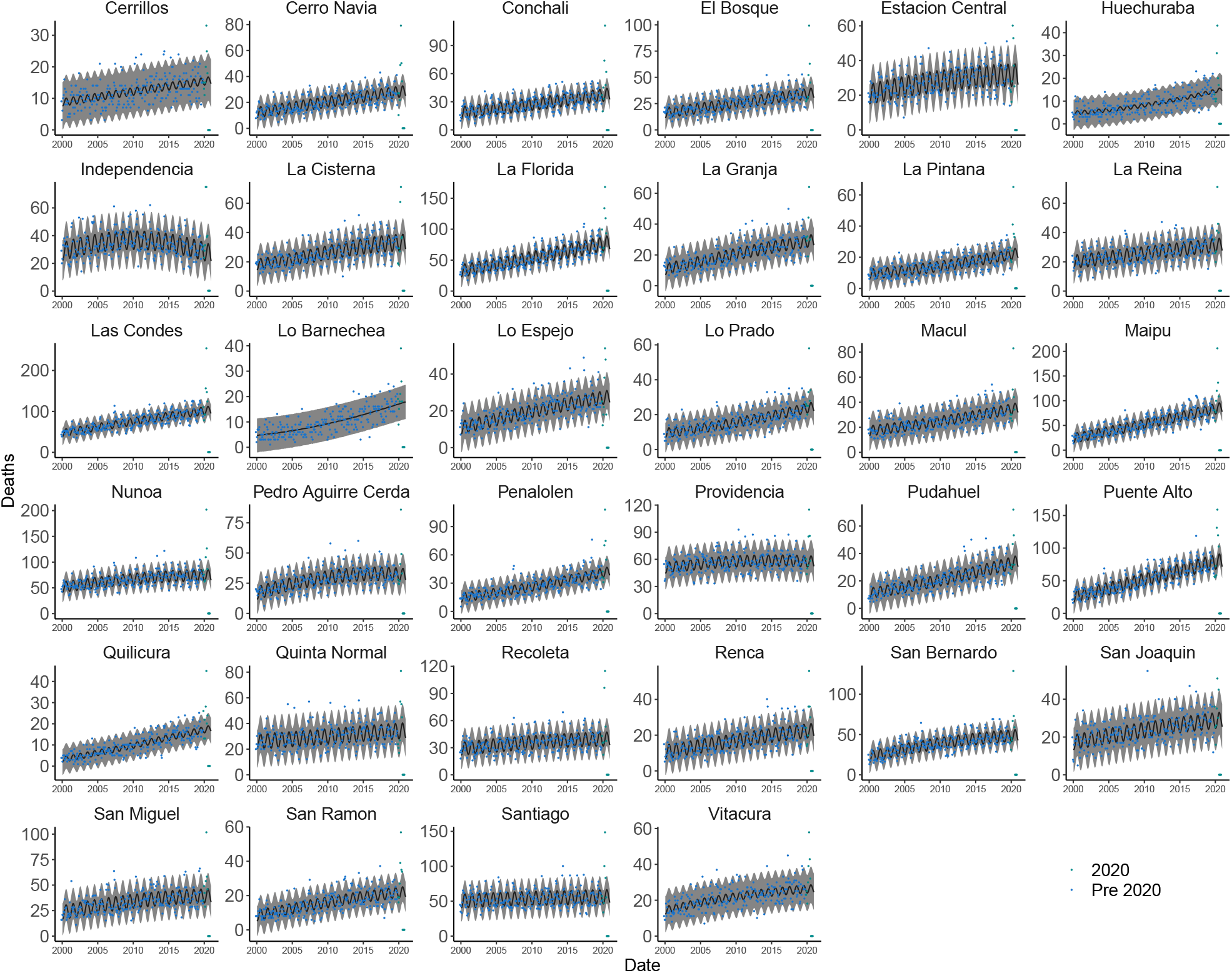
Number of monthly deaths in 2002-2020 in urban municipalities, 80+ age group. GP predictions and confidence intervals are shown as black line surrounded by gray areas, respectively

**Figure S30:**
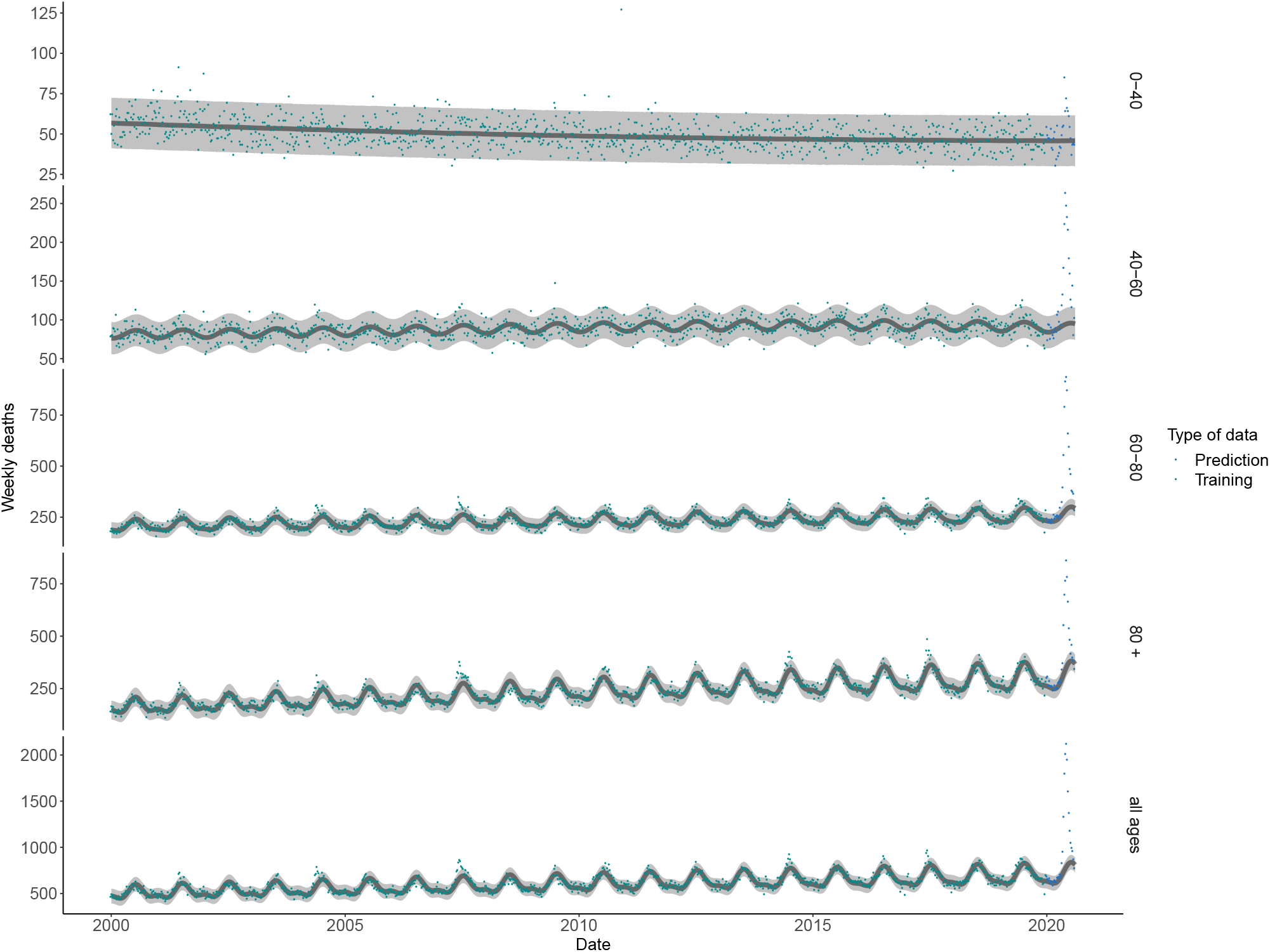
Historical weekly mortality in the *Greater Santiago*, by age group.

**Figure S31:**
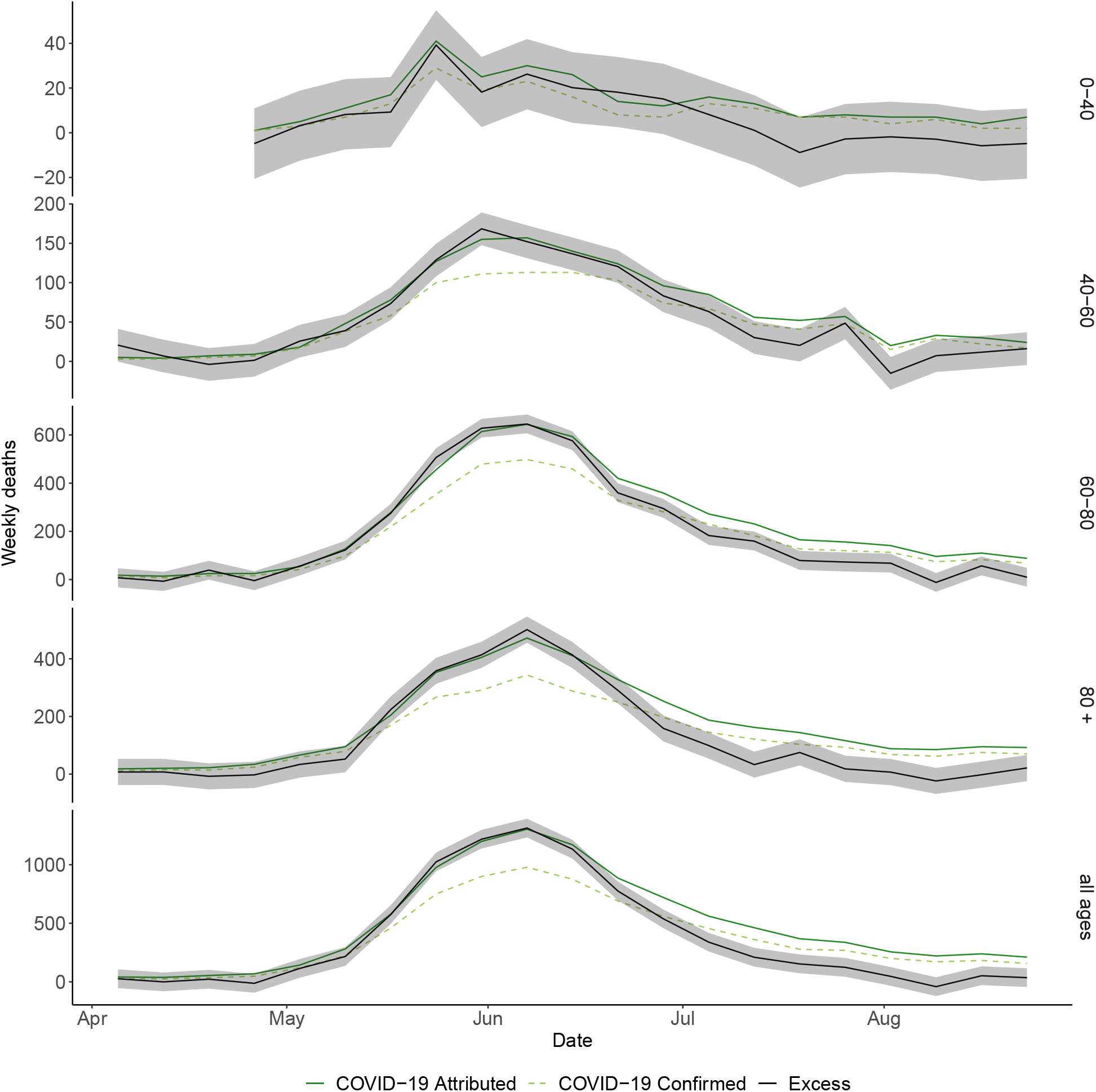
Weekly excess deaths in Region Metropolitana. until August 1st, 2020. This supplements Fig.2C in the main text, showing the time-varying excess for each age group.

**Figure S32:**
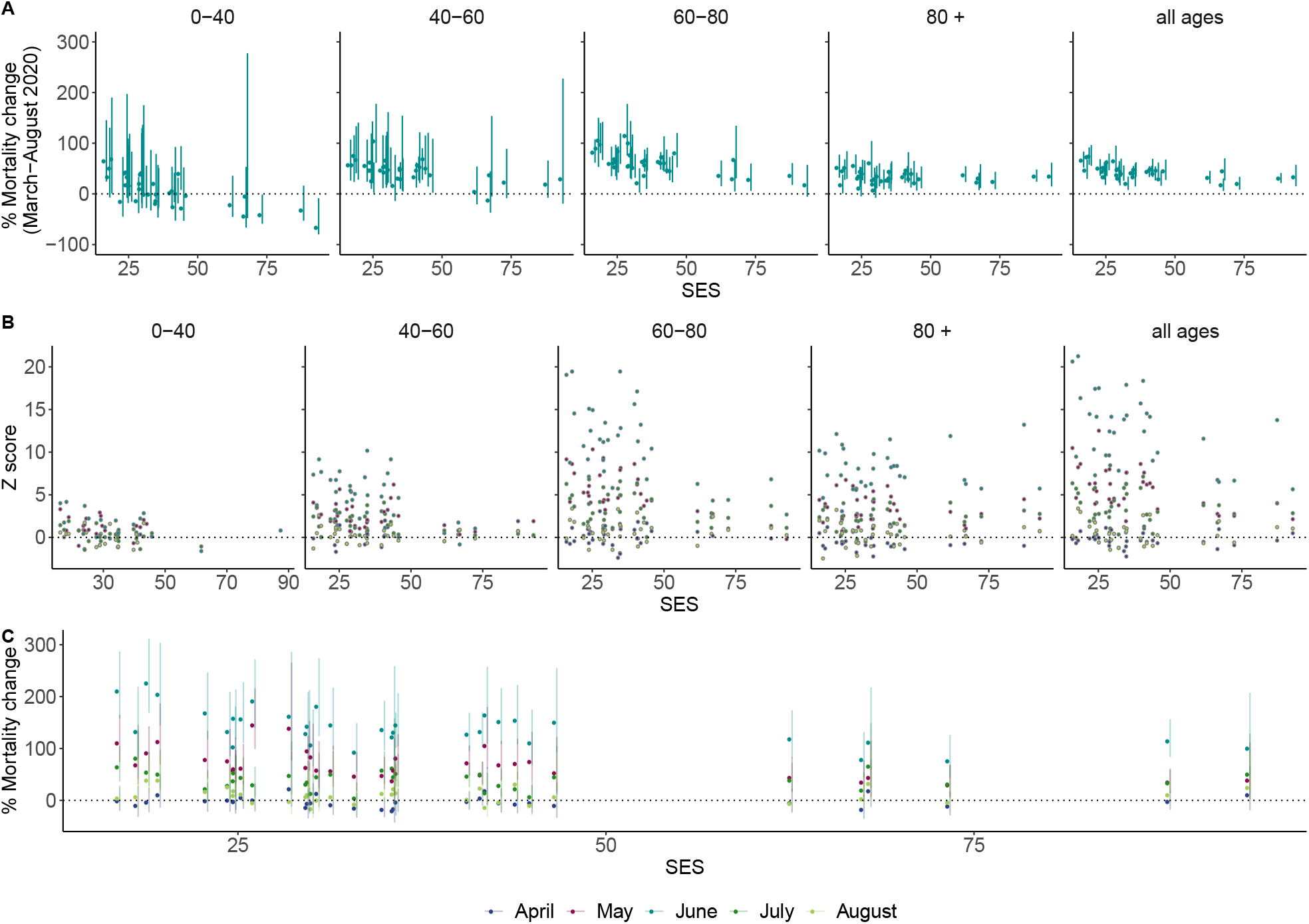
Relation between Z-scores and *social priority index* for different age groups. This figure supplements Fig 2E in the main text: here, **B** is exactly Fig 2E but we show Z-scores of each month, instead of the entire period. On **A** and **C** we show an alternative to the z-score: the % of change with respect to what was expected for this year. The uncertainty that otherwise is captured in the *z* − *score* is now reflected in the length of the confidence intervals (see for example [34]) for this quantity. More specifically, **C** is analog with **B** with this alternative measure, while **A** shows the overall change, collapsing all months.

**Figure S33:**
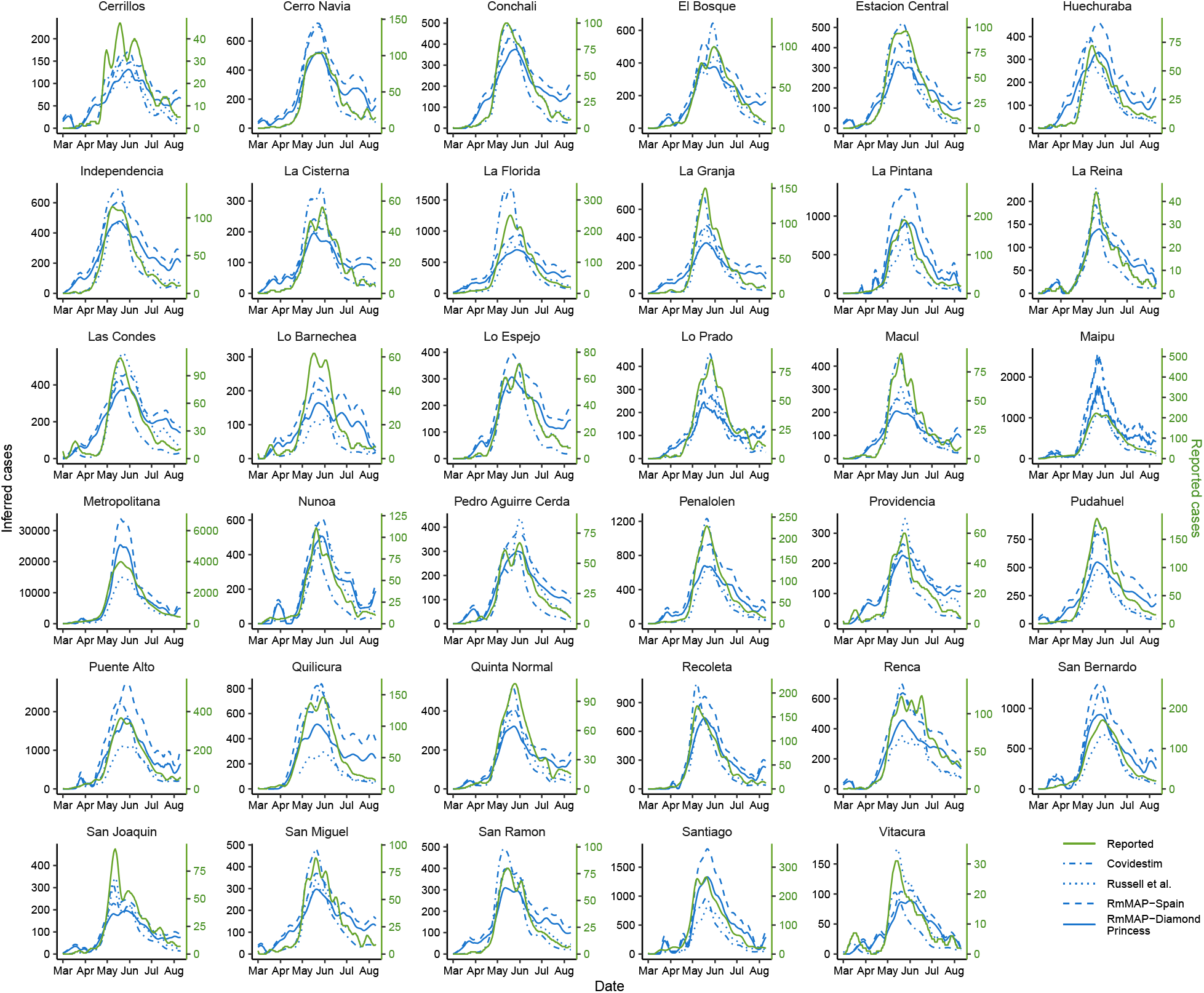
Reconstruction of true epidemic curves at municipalities of *Greater Santiago*. This figure supplements the reconstructions in the main text (Fig. 3A) by displaying results for all municipalities, for all methods and a longer period. We compared our regularized Richardson-Lucy method using two age-stratified IFR (from Spain and Diamond Princess) with the off-the-shelve Covidestim [23] and Russell’s [24] method.

**Figure S34:**
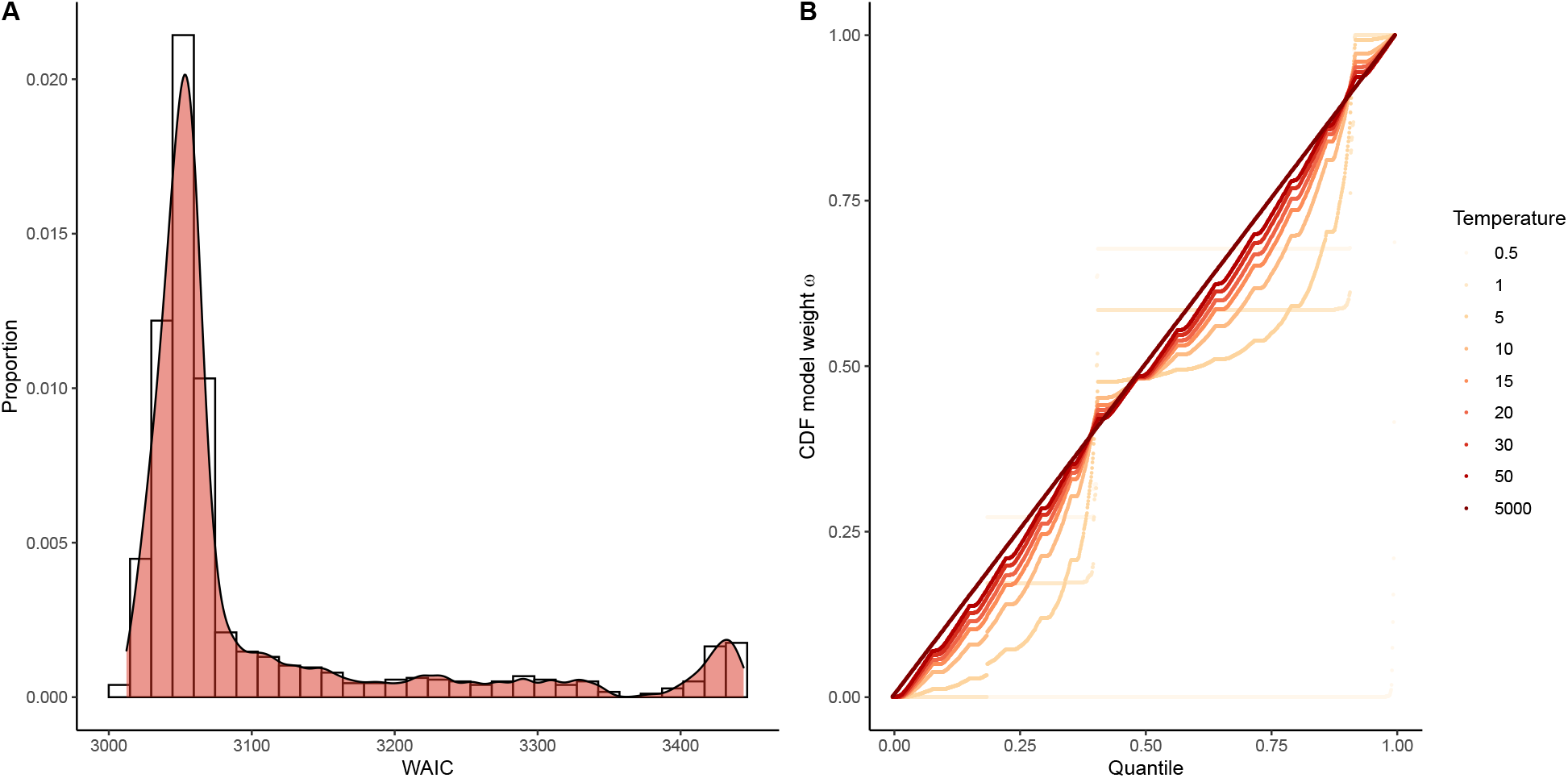
Mixture of models. **B** histogram of WAICS (that we use as a proxy for model weights) show heterogeneity. **B** CDF of model weights for different temperature (*τ*) values. Our model averaging criterion is based on a tempering of the WAICS. Here, we show that if *τ* is too small only one or two models are effectively weighted, and as we increase the temperature more models are chosen until eventually all models are weighted equally.

**Figure S35:**
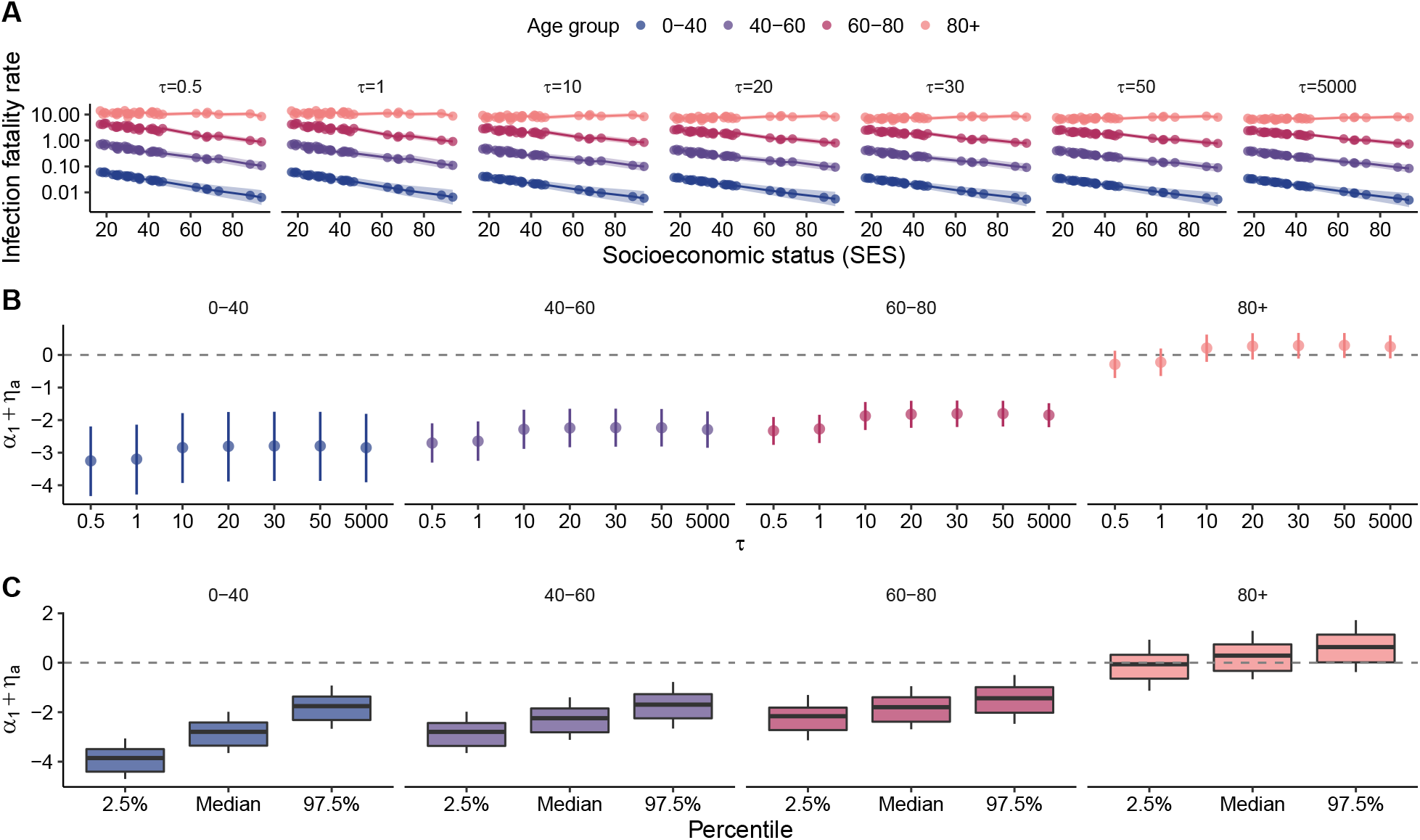
Robustness analysis for IFR linear model. (equation (17), with or without the spatial random effect *υ*_*m*_). **A** IFR as a function of SES, for different model averages (*τ*); Fig. 5B in the main text corresponds to *τ* = 1. **B** the sign of *α*_1_ + *η*_*a*_ indicates whether IFRs are decreasing as a function of SES. We show that for all model averages (*τ*) this coefficient is significantly smaller than zero in all age groups with the exception of the oldest, where the relation is unclear. **C** Similar to **B**, but now we show boxplots of relevant percentiles of this coefficient where each model contributes with a sample.

**Figure S36:**
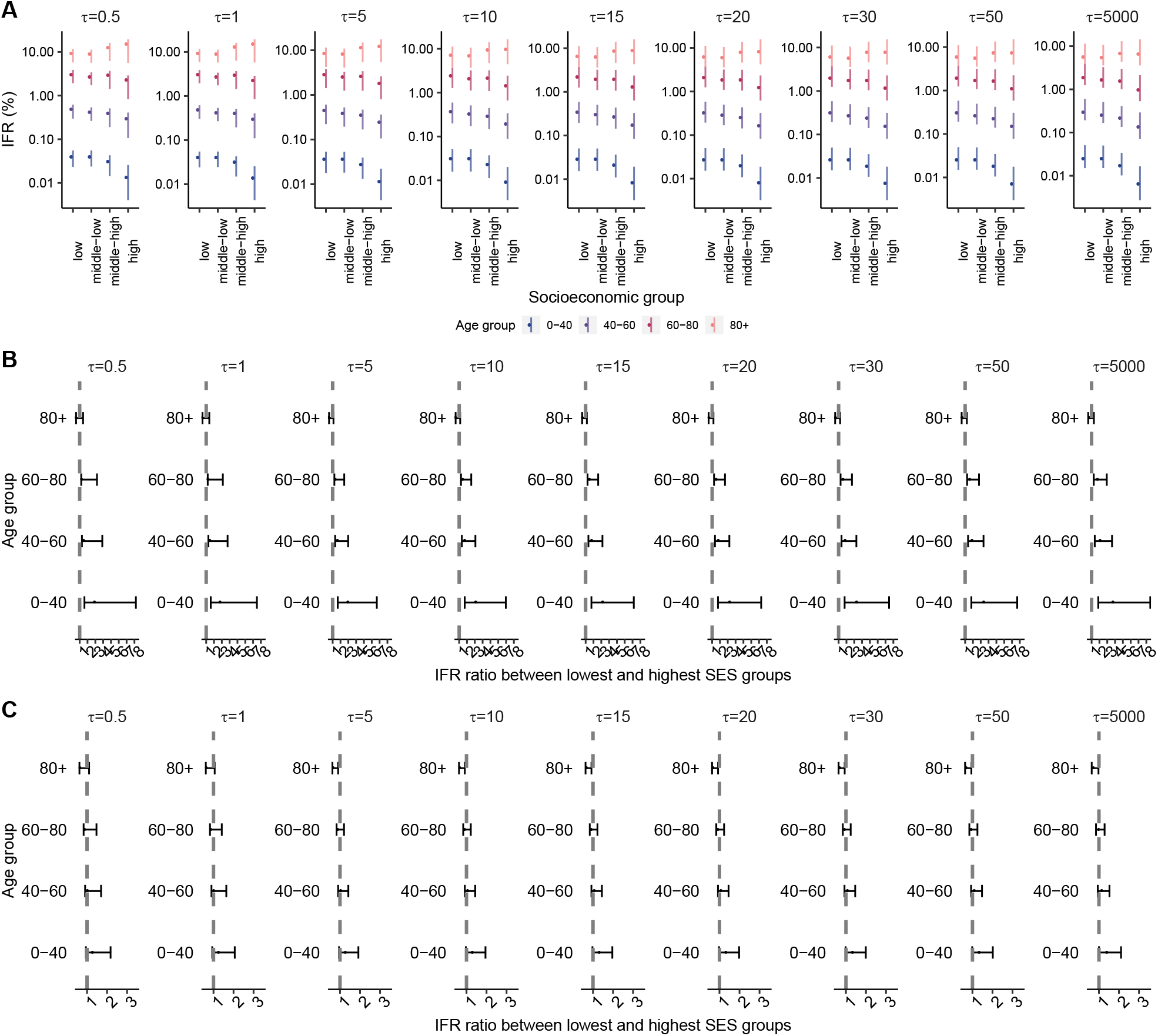
Robustness analysis for IFR categorical model (equation (26)). **A** IFR (95% credible interval) as a function of SES strata, for different model averages (*τ*).**B** IFR ratio between lowest and highest SES strata as a function of *τ* (Fig. 5C in the main text corresponds to *τ* = 1). **C** For completeness, we also show IFR ratio between middle-low and middle-high SES strata. The effects here appear attenuated and more often are not significant.

**Figure S37:**
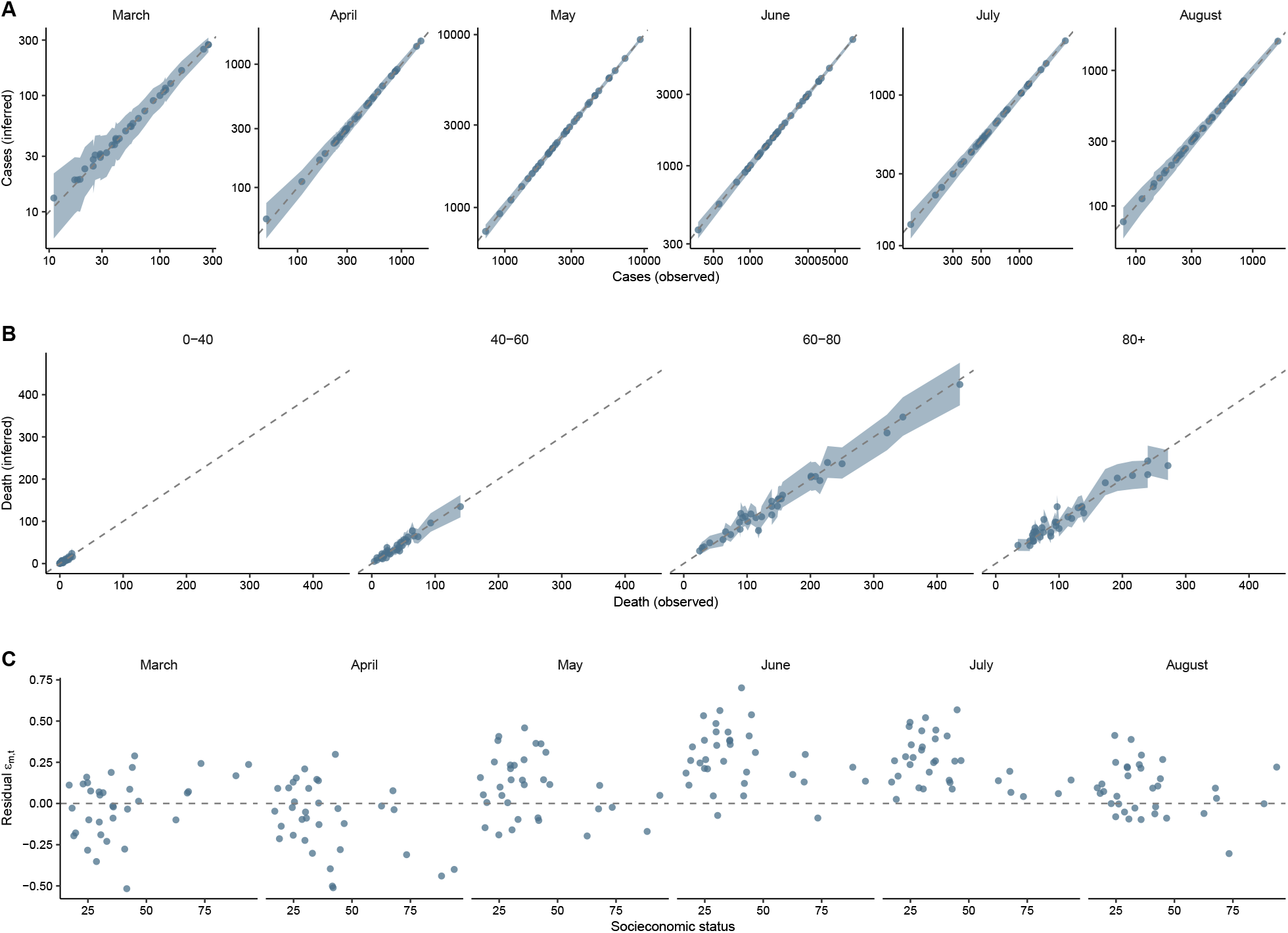
Posterior predictive checks. **A** Predicted cases (with 95% credible intervals) of our model, along with recorded cases for each months. **B** Predicted deaths for each age group. Here predictions are less aligned with true values, although the somewhat wider credible intervals still cover the parameter. **C** Residuals (posterior median) *E*_*m,t*_ across all months (*t*) and municipalities *m* (represented though their SES).

**Figure S38:**
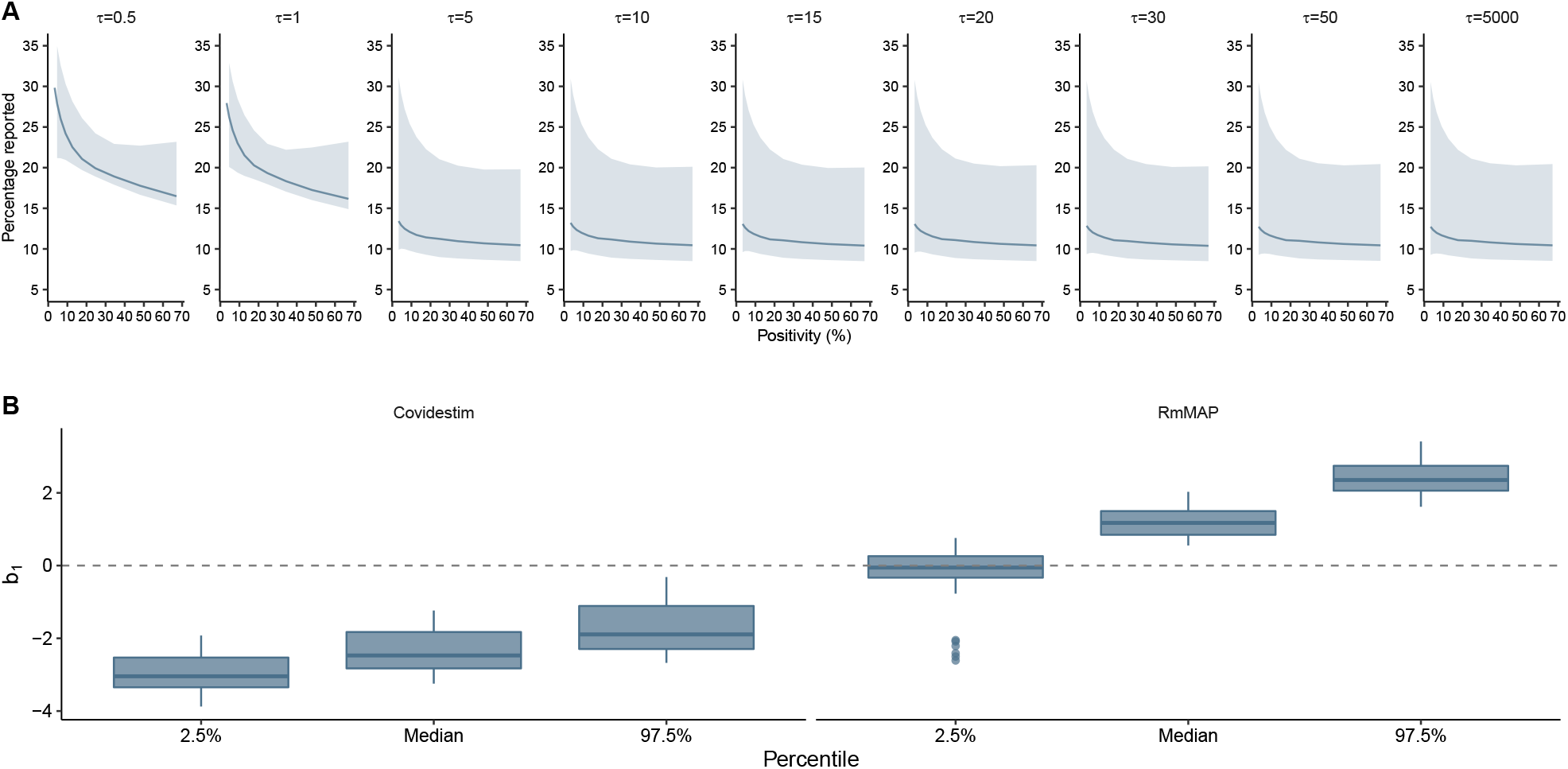
Estimation of ascertainment rates. **A** Inferred proportion of cases reported (with 95% credible bands) as a function of positivity rates for our mixture of models. **B** Boxplot of relevant percentiles for the coefficient *b*_1_ ,indicating the strength of the relation between log-positivity and logit ascertainment. For models whose prior on *γ* is based on Covidestim estimates the inferred relation is always negative, and the opposite (less intuitive) pattern emerges when using *RmMAP*.

**Figure S39:**
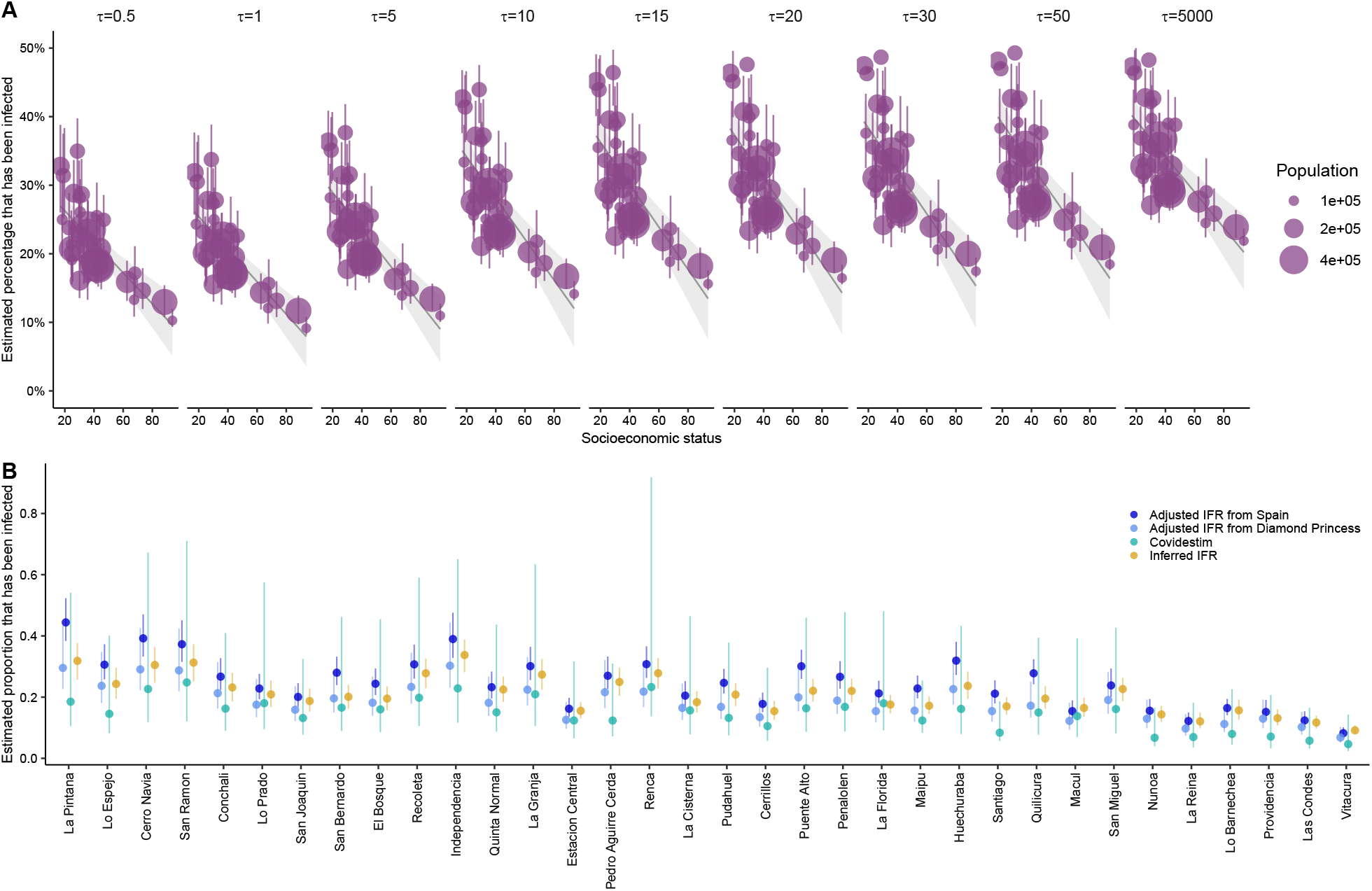
Estimation of proportion infected. **A** Inferred proportion of people who were infected for all different municipalities (sorted by SES), and for many mixtures of models with different temperature *τ* . **B** Comparison of proportion infected by all available methods; the ones based on a fixed IFR (*RmMAP*-type), Covidestim, and our method in S1.4.

**Figure S40:**
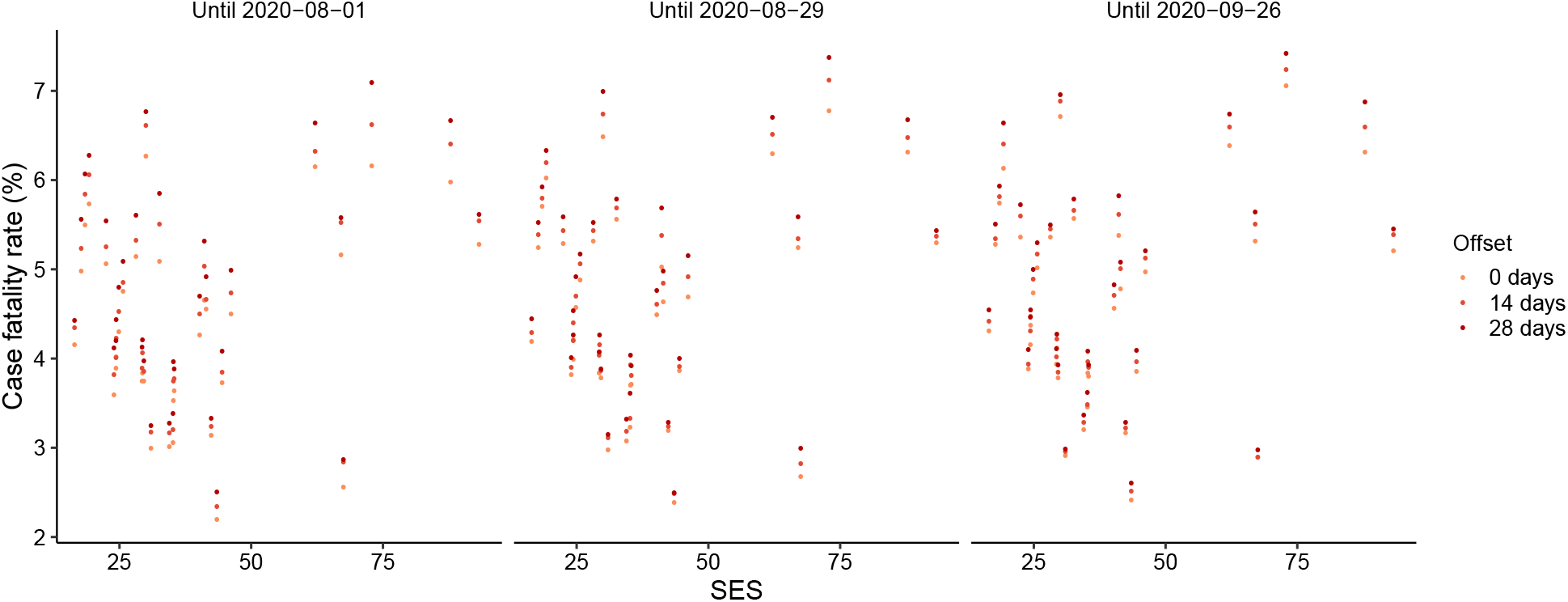
Stability of CFR estimates for slighly different designs. We show lump CFR estimates at the municipality level (marginalizing over age information) using slight variations of the naive *cfr*_*t*_ = *deaths*_*t*_*/cases*_*t*_ formulas, where *t* represents the oldest date until which *cases*_*t*_ and *deaths*_*t*_ are recorded. To acknowledge the possibity of bias in estimated CFR because of right truncation, we computed the ratios with an offset of Δ after *t*, so that this new CFR estimate may better grasp deaths that would occur in the future.

**Figure S41:**
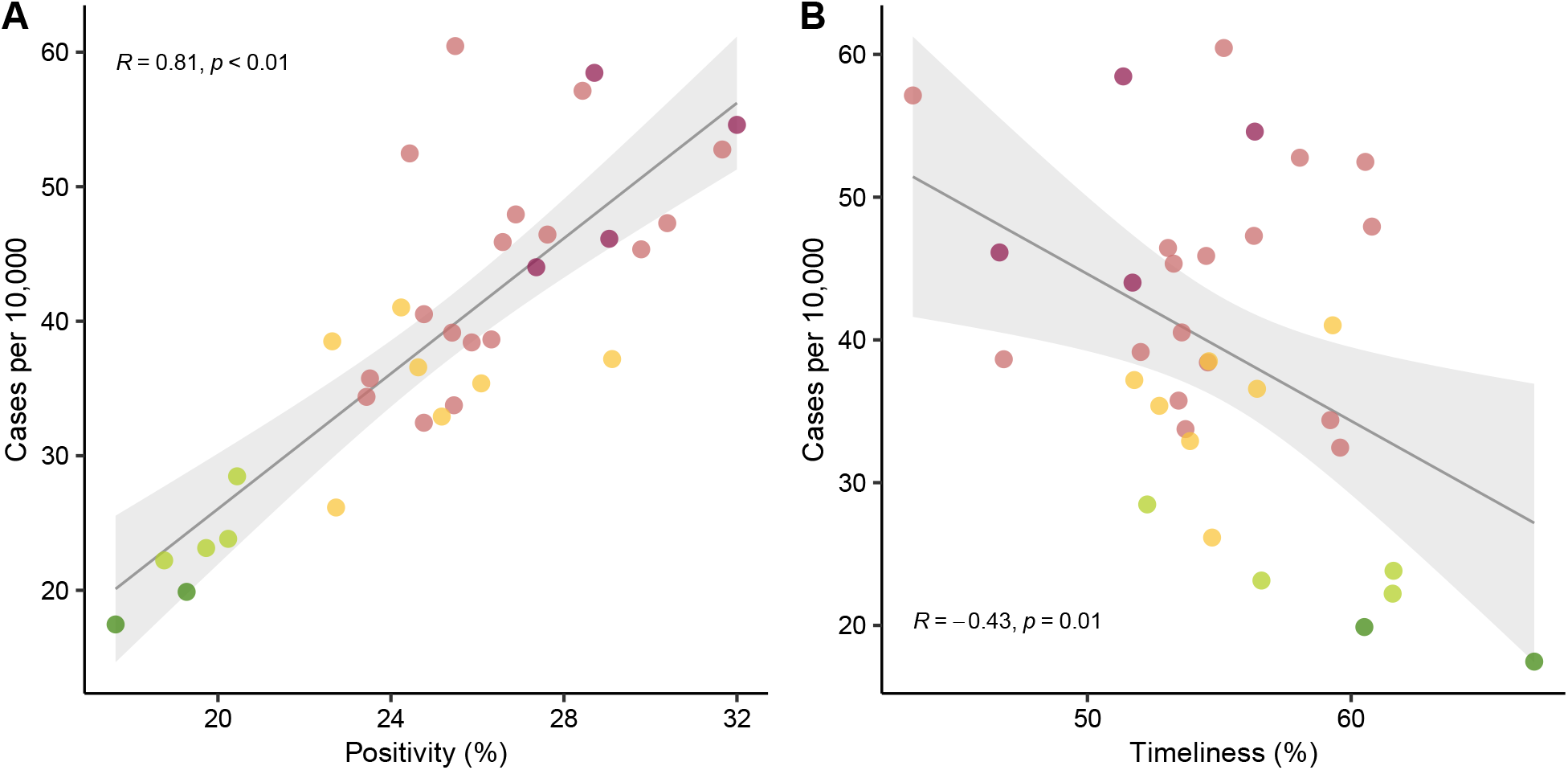
Positivity and timeliness as a function of cases. This figure extend Figs. 4C and 4G in the main text, by showing *averages* over weeks of cases, timelinesses and positivities. **A** Average of weekly cases as a function of average positivity **B** Average of weekly cases as a function of average timeliness.

**Table S1:** Empirical lower and upper coverage probabilities (percentage of times data fell below or above the 2.5 and 97.5 percentiles from samples of model, respectively, on data from 2000 to 2019) for each urban municipality and age group, and also for the entire Region Metropolitana (last row).

|  |  |  |  |  |  | 0-40 |  | 40-60 |  | 60-80 |  | 80+ |  | All ages |  |
| --- | --- | --- | --- | --- | --- | --- | --- | --- | --- | --- | --- | --- | --- | --- | --- |
| 2-3 | 4-5 | 6-7 | 8-9 | 10-11 |  | lower | upper | lower | upper | lower | upper | lower | upper | lower | upper |
|  |  |  |  |  | Municipality |  |  |  |  |  |  |  |  |  |  |
|  |  |  |  |  | Cerrillos | 3.06 | 96.9 | 2.94 | 97.1 | 4.58 | 96.2 | 5. | 95.8 | 5.42 | 95.4 |
|  |  |  |  |  | Cerro Navia | 4.58 | 95.4 | 5.42 | 97.5 | 3.33 | 96.7 | 4.17 | 97.5 | 2.92 | 98.8 |
|  |  |  |  |  | Conchali | 4.60 | 95.8 | 5.83 | 96.7 | 4.58 | 97.5 | 5. | 95.8 | 4.17 | 97.1 |
|  |  |  |  |  | El Bosque | 3.75 | 96.7 | 2.08 | 98.8 | 2.08 | 98.3 | 3.75 | 98.3 | 4.17 | 97.1 |
|  |  |  |  |  | Estacion Central | 3.77 | 96.7 | 5.83 | 94.6 | 3.75 | 97.5 | 4.58 | 97.5 | 5. | 96.7 |
|  |  |  |  |  | Huechuraba | 2.98 | 97.0 | 4.60 | 95.8 | 3.75 | 97.9 | 4.20 | 97.1 | 2.92 | 97.9 |
|  |  |  |  |  | Independencia | 3.39 | 96.6 | 2.5 | 97.5 | 3.75 | 97.1 | 4.17 | 97.1 | 5. | 96.7 |
|  |  |  |  |  | La Cisterna | 3.72 | 96.3 | 3.35 | 98.3 | 4.17 | 97.1 | 3.33 | 97.5 | 2.5 | 98.3 |
|  |  |  |  |  | La Florida | 4.17 | 97.1 | 7.08 | 94.6 | 3.75 | 97.5 | 4.17 | 97.5 | 4.17 | 97.1 |
|  |  |  |  |  | La Granja | 4.22 | 96.6 | 3.75 | 97.1 | 3.75 | 98.3 | 2.92 | 97.5 | 4.17 | 97.1 |
|  |  |  |  |  | La Pintana | 5. | 97.1 | 2.92 | 97.9 | 4.58 | 96.2 | 3.75 | 97.5 | 3.75 | 97.5 |
|  |  |  |  |  | La Reina | 3.21 | 96.8 | 2.51 | 97.5 | 5.83 | 95.8 | 3.75 | 97.5 | 3.75 | 97.1 |
|  |  |  |  |  | Las Condes | 4.17 | 95.8 | 6.25 | 96.7 | 4.58 | 96.7 | 3.75 | 97.9 | 3.75 | 97.5 |
|  |  |  |  |  | Lo Barnechea | 5.26 | 94.7 | 4.82 | 95.2 | 4.18 | 96.2 | 5. | 95.8 | 6.25 | 95.8 |
|  |  |  |  |  | Lo Espejo | 3.38 | 97.0 | 6.25 | 95 | 4.17 | 96.7 | 4.58 | 96.2 | 5.42 | 96.2 |
|  |  |  |  |  | Lo Prado | 3.00 | 97.4 | 5. | 96.2 | 2.08 | 98.3 | 5. | 96.7 | 4.58 | 96.7 |
|  |  |  |  |  | Macul | 3.88 | 96.1 | 3.33 | 96.7 | 3.75 | 98.3 | 4.58 | 95.8 | 5. | 95.8 |
|  |  |  |  |  | Maipu | 5.42 | 96.2 | 4.58 | 97.1 | 4.17 | 97.5 | 5. | 97.1 | 4.17 | 97.5 |
|  |  |  |  |  | Nunoa | 4.24 | 95.8 | 5. | 96.7 | 5.42 | 95.8 | 3.75 | 96.7 | 3.33 | 97.5 |
|  |  |  |  |  | Pedro Aguirre Cerda | 4.62 | 96.2 | 4.58 | 97.1 | 4.58 | 97.5 | 4.17 | 96.2 | 2.5 | 99.2 |
|  |  |  |  |  | Penalolen | 5. | 96.7 | 5.83 | 95.8 | 4.58 | 97.1 | 3.75 | 96.7 | 3.75 | 97.1 |
|  |  |  |  |  | Providencia | 3.07 | 96.9 | 4.58 | 97.1 | 4.17 | 96.7 | 4.58 | 96.7 | 3.75 | 97.1 |
|  |  |  |  |  | Pudahuel | 5.42 | 96.7 | 3.75 | 97.1 | 5. | 97.5 | 3.33 | 97.5 | 3.33 | 97.1 |
|  |  |  |  |  | Puente Alto | 5. | 95.8 | 4.17 | 97.1 | 5. | 96.7 | 3.33 | 97.9 | 3.75 | 97.1 |
|  |  |  |  |  | Quilicura | 5.83 | 95.4 | 5.83 | 95.8 | 4.17 | 96.7 | 5.46 | 97.5 | 3.75 | 98.3 |
|  |  |  |  |  | Quinta Normal | 3.00 | 97.0 | 5. | 97.1 | 3.33 | 97.9 | 3.33 | 96.7 | 4.58 | 96.7 |
|  |  |  |  |  | Recoleta | 4.17 | 97.5 | 5. | 96.2 | 4.17 | 97.1 | 4.58 | 96.2 | 3.33 | 97.5 |
|  |  |  |  |  | Renca | 2.5 | 97.9 | 3.75 | 97.5 | 4.17 | 97.1 | 4.58 | 96.7 | 5. | 96.2 |
|  |  |  |  |  | San Bernardo | 3.75 | 96.7 | 3.33 | 97.5 | 4.58 | 98.8 | 4.58 | 97.1 | 4.17 | 96.7 |
|  |  |  |  |  | San Joaquin | 3.03 | 97.0 | 4.18 | 97.1 | 5. | 96.2 | 3.33 | 97.5 | 4.58 | 97.1 |
|  |  |  |  |  | San Miguel | 0.427 | 99.6 | 2.08 | 97.9 | 2.08 | 98.3 | 4.58 | 96.2 | 4.17 | 96.7 |
|  |  |  |  |  | San Ramon | 4.70 | 95.3 | 4.58 | 96.7 | 4.58 | 97.5 | 3.33 | 97.5 | 3.33 | 97.9 |
|  |  |  |  |  | Santiago | 4.58 | 95.8 | 5.83 | 95.8 | 4.17 | 97.5 | 5.42 | 95.8 | 5.83 | 97.1 |
|  |  |  |  |  | Vitacura | 2.66 | 97.3 | 4.98 | 95.0 | 3.75 | 97.5 | 5. | 97.1 | 3.75 | 97.1 |
|  |  |  |  |  | Region Metropolitana | 3.35 | 97.8 | 4.41 | 97.6 | 4.98 | 96.0 | 4.89 | 96.7 | 4.50 | 96.8 |

**Table S2:** Summary of the main variations of the model; we considered changes over the functional form of *IFR* and for the prior of *γ*, the temporal profile of infections per municipality.

| Feature of model | Original formulation | Variation 1 | Variation 2 |
| --- | --- | --- | --- |
| $\text{logit}(IFR_{m,a}) =$ | $\alpha_0 + (\alpha_1 + \eta_a) \times SES_m + \delta_a$ | $\alpha_0 + (\alpha_1 + \eta_a) \times SES_m + \delta_a + v_m$ | $\alpha_0 + \psi_{q(m),a}$ |
| Prior on $\gamma$ | from Covidestim | from RmMAP | n.a. |

**Table S3:** Total deaths in *Greater Santiago* a occurred between 2020-01-01 and 2020-08-30 according for four subsequent weekly death reports by DEIS

| Date reported | 2020-09-24 | 2020-10-01 | 2020-10-08 | 2020-12-10 |
| --- | --- | --- | --- | --- |
| Total deaths | 37,386 | 37,387 | 37,393 | 37,410 |

